# Barriers and enablers to blood culture sampling: a systematic review and theoretical domains framework survey in Indonesia, Thailand and Vietnam

**DOI:** 10.1101/2022.11.19.22282535

**Authors:** Pornpan Suntornsut, Koe Stella Asadinia, Ralalicia Limato, Alice Tamara, Linda W.A. Rotty, Rendra Bramanti, Dwi U. Nusantara, Erni J. Nelwan, Suwimon Khusuwan, Watthanapong Suphamongkholchaikul, Parinya Chamnan, Watcharapong Piyaphanee, Vu Thi Lan Huong, Nguyen Hai Yen, Khanh Nguyen Hong, Pham Ngoc Thach, Le Minh Quang, Vu Hai Vinh, Chau Minh Duc, Vo Thi Hoang Dung Em, Elinor Harriss, H Rogier van Doorn, Raph L. Hamers, Fabiana Lorencatto, Lou Atkins, Direk Limmathurotsakul

## Abstract

**Background:** Blood culture (BC) sampling is recommended for all suspected sepsis patients prior to antibiotic administration. Here, we aimed to identify barriers and enablers to BC sampling in three Southeast Asian countries.

**Methods:** We conducted a systematic review of studies evaluating barriers/enablers to BC sampling from 1900 to 2020 globally (PROSPERO, CRD42020206557). Using the findings of the systematic review, we developed and conducted a Theoretical Domains Framework (TDF)-based survey with a case scenario question among doctors and final-year medical students in Indonesia, Thailand and Vietnam.

**Findings:** In the systematic review, we identified 6,175 unique records from the databases, of which 25 met the eligibility criteria. Studies were conducted in 37 high-income countries (HICs) and 41 low-and middle-income countries (LMICs). Of 14 TDF domains, three and seven were not assessed in HICs and LMICs by the studies included in the systematic review, respectively. 1,070 medical doctors and 238 final-year medical students completed the survey. The proportion of respondents who would definitely take BC in the case scenario was 89.8% for Thai, 50.5% for Vietnamese and 31.3% for Indonesians (p<0.001). Eight TDF domains were considered key in influencing BC sampling, including ‘no awareness of guideline [TDF-knowledge]’, ‘low priority of BC [TDF-goals]’, ‘no intention to follow guidelines [TDF-intention]’, ‘level of doctors who can order or initiate an order for BC [TDF-social professional role and identity]’, ‘no norms of BC sampling [TDF-social influence]’, ‘perceived cost-effectiveness of BC [TDF-environmental context and resources]’, ‘regulation on cost reimbursement [TDF-behavioural regulation]’ and ‘consequences that discourage BC sampling [TDF-reinforcement].’ However, there was substantial heterogeneity between the countries across most domains.

**Conclusions:** Evidence on barriers and enablers to BC sampling is limited globally. We identified individual, socio-cultural and environmental barriers/enablers to BC sampling across different countries, which represent potential targets for interventions. Context-specific multifaceted interventions at both hospital and policy levels are required to improve diagnostic stewardship practices.

**Funding:** Wellcome Trust, UK (220557/Z/20/Z).

## Introduction

Blood culture (BC) is a crucial diagnostic, which can guide antibiotic treatment decisions of severe bacterial infections, and may improve patient outcomes.^1,2^ The cumulative results of BC are also crucial to inform antimicrobial resistance (AMR) surveillance, at the hospital, country and global levels.^3^ International guidelines on sepsis management have been stressing the importance of obtaining BC before or, when not possible, within 24 hours after administration of antibiotics.^1,4^

Nonetheless, BC sampling rates are generally underutilized, both in high-income countries (HICs) and low and middle-income countries (LMICs), with wide variations in reported rates between hospitals and global regions. Reported hospital BC sampling rates ranged from 196 to 308 per 1,000 patient-days in the United States,^5,6^ from 6.7 to 86.5 per 1,000 patient-days in the European Union,^7^ from 0 to 82 per 1,000 patient-days in the Central Asian and European Surveillance of AMR network (CAESAR),^8^ and 31, 82 and 10 per 1,000 patient-days in selected hospitals in Indonesia,^9^ Thailand^10^ and Vietnam^11^, respectively.

A range of barriers and enablers have been identified that influence BC sampling, based on different study designs, theories and frameworks. Lack of clear guidelines, training, microbiological infrastructure, and positive attitudes regarding BC among medical practitioners, are commonly reported barriers.^8,12-15^ However, to date the evidence has not been systematically assessed or integrated.

Changing the behavior of medical practitioners is complex, and systematic approach have been shown useful to understand factors influencing adherence to guidelines or recommendations so as to inform the design of future interventions.^16-18^ The Theoretical Domains Framework (TDF) has been developed by synthesizing a wide range of theories, and enabling researchers to investigate a broader range of individual, socio-cultural and environmental behavioral influences than they would with a single theory alone.^16-18^ The TDF has been widely used to explore barriers and enablers to healthcare professional behaviors, including diagnostic testing, antimicrobial stewardship, and infection prevention control.^19-22^

Here, we aimed to identify barriers and enablers to BC sampling in three middle-income countries in Southeast Asia (SEA) using a theory-based approach informed by the TDF. Specifically, we aimed to integrate available evidence by conducting (a) a global systematic literature review, and (b) a TDF survey among doctors and final-year medical students in Indonesia, Thailand and Vietnam

## Methods

### Systematic review

We conducted a systematic review by searching five bibliographic databases (PubMed, Ovid Embase, Scopus, Web of Science, the Cochrane Database of Systematic Reviews and Databases of Systematic Reviews) for studies evaluating and reporting primary data on barriers/enablers to BC sampling, published in English between 1 January 1900 to 31 June 2020 (PROSPERO, CRD42020206557). We used the search terms “blood culture”, “bloodstream infection” or “diagnostic stewardship”, and “barriers”, “enablers”, “facilitators”, “knowledge”, “attitude” or “practice” (Appendix S1 and S2). We additionally searched the reference list of all relevant articles to identify additional articles. We included qualitative studies, mixed-method studies, quantitative descriptive studies, quantitative randomized controlled trials, and quantitative non-randomized interventional studies.^23^

Two investigators (PS and DL) independently screened records for eligible studies based on title and abstract. Full texts of articles deemed potentially eligible were retrieved and independently screened by two investigators (PS and DL) for final inclusion. Data were extracted and assessed using a standardised data abstraction form. Data included study characteristics, sample size, criteria used to define Theory Domain Frameworks (TDF) domains, and reported barriers/enablers. Study type was categorized and quality assessment of the evidence was performed using the Mixed Methods Appraisal Tool (MMAT).^24^ In the event of different conclusions, a third reviewer (LA) adjudicated to determine a conclusion.

### TDF survey

We developed a TDF survey questionnaire, comprising a case scenario and all 14 TDF domains of barriers/enablers to BC sampling, through an iterative process of several rounds of review, based on the findings of our systematic literature review, a previous study (included in the review) which used case scenarios in which BC ordering is strongly recommended by international guidelines,^13^ and previous TDF surveys on other health topics.^25-28^ Each question used a five-point Likert scale representing the level of perceived barriers/enablers to BC sampling under all TDF domains (Appendix S3).

The initial questionnaire was translated into Thai, Vietnamese and Indonesian language and piloted among 3-6 final-year medical students and 10-19 medical doctors in each country (a total of 54 respondents) to test the clarity of questions and choice answers in each language and to ensure no potential key barriers/enablers were omitted. We asked respondents to complete the survey and provide feedback using 1:1 interviews via phone or using online meeting software. The questionnaire was revised and finalized based on the pilot study results. One free-text question was added (i.e. Question 6-5, “Additional comments about emotional factors…” [Appendix S4]), a total of 27 choice answers were added, and languages and wordings were revised. The final questionnaire included 54 questions about barriers/enablers to BC sampling and respondents’ demographic characteristics (Appendix S4).

Based on a sample size calculation for descriptive studies (appendix S1), our enrolment target was 1500 respondents (100 final-year medical students and 400 medical doctors in each of the three countries). We used a convenient sampling approach, distributing invitations in letters, emails, pamphlets and online social media platforms, through existing collaborations in hospitals and universities in the three survey countries. The online survey was conducted using the Qualtrics survey platform.

For the pilot, verbal informed consent was obtained from study respondents prior to participation. For the TDF survey, electronic informed consent was obtained from study respondents prior to participation. The study was approved by the Oxford University Tropical Research Ethics Committee (OXTREC545-21) and local ethical committees at Iskak Tulungagung Hospital (070/7303/407.206/2021), Prof. Dr. R.D. Kandou Hospital (156/EC/KEPK-KANDOU/IX/2021), Pasar Minggu Hospital (EOCRU/RCH.216/10.2021/1145) in Indonesia, The National Hospital for Tropical Diseases (14HDDD/NDTU) in Vietnam, Faculty of Tropical Medicine, Mahidol University (TMEC21-069), Sunpasitthiprasong Hospital (065/64S) and Chiangrai Prachanukroh Hospital (CR 0032.102/EC023) in Thailand.

### Analysis

For each question, we defined that respondents who answered “definitely”/”likely”, “all the time”/”often” or “strongly agree”/”agree” perceived the importance or agreement with that barrier/enabler. The proportion of respondents who answered likewise, after excluding respondents who answered ‘I do not know’ or ‘I do not want to answer’, was present. Groups were compared by Chi-squared or Fisher exact tests as appropriate. Logistic regression models with random effects for countries, for hospital type nested in the same country, and for professional roles nested in the same hospital type were used to evaluate the association between respondents’ answers about each barrier/enabler and to the case scenario. Statistical analyses were performed using Stata 15.1 (StataCorp, US).

We integrated the results from the systematic review and TDF survey to identify and rank important TDF domains for Indonesia, Thailand and Vietnam. Each domain identified was scored on an established set of four ‘importance criteria’ (modified from a previous TDF study^29^): (1) ‘frequency’ (number of respondents and studies that identified each domain); (2) ‘elaboration’ (number of themes) within each domain; (3) ‘expressed importance’ (either a statement from the authors’ interpretation or direct quotes from study respondents expressing importance); and (4) ‘association between reported barriers/enablers and BC practice’ (the answer to the case scenario was used as a proxy for BC practice).

Lastly, we mapped identified TDF domains to the COM-B (‘capability’, ‘opportunity’, ‘motivation’ and ‘behaviour’) model.^16-18^ COM-B forms the hub of the Behaviour Change Wheel (BCW), a framework which signposts to potentially relevant intervention strategies. This allowed us to list all intervention types and policy options that were likely to be effective in addressing identified barriers and enablers.

### Role of the funding source

The funders of the study had no role in the study design, data collection, data analysis, data interpretation, or writing of the report.

## Results

### Study characteristics

#### Systematic review

We identified 6,175 unique records from the databases (Figure 1, Appendix S5). After title and abstract screening, 884 articles were selected for full-text review. 859 articles were excluded because they did not meet the eligibility criteria and 25 original studies were included (Appendix S6). Those studies were conducted in or included participants from 37 HICs and 41 LMICs (Figure 2, Appendix S7), and comprised 15 (60%) quantitative studies, six (24%) quantitative non-randomized interventional studies, two (8%) qualitative studies, one mixed-method study, and one quantitative randomized controlled trial (4% each). Of 25 studies, eight (32%) were judged to be at low risk of bias, 14 (56%) at medium risk of bias and three (12%) at high risk of bias (Appendix S8).

**Figure 1:**
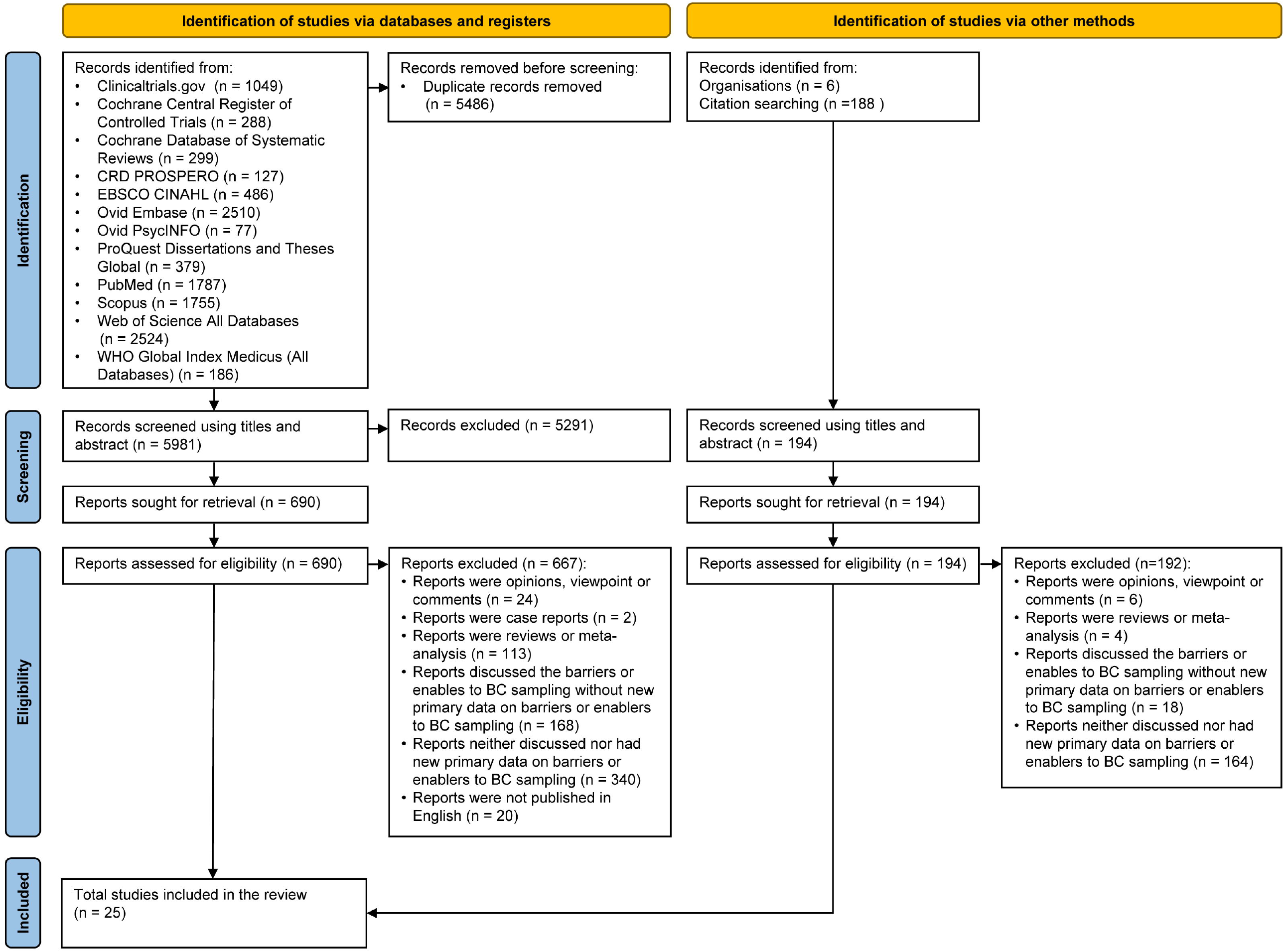
Flow diagram of the study selection process for the systematic review

**Figure 2:**
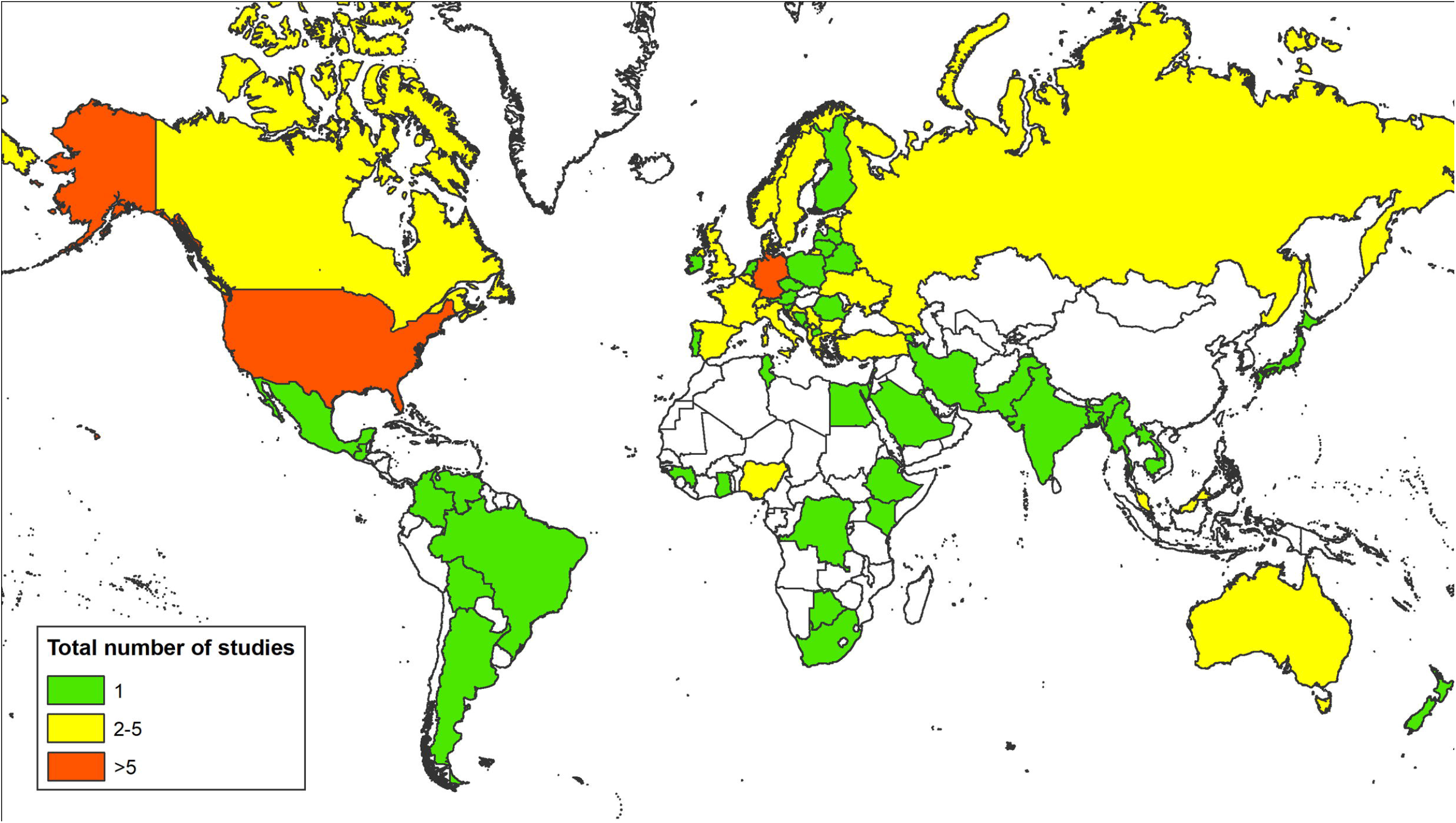
Global map visualizing the number of studies included in the systematic review. Note. the map shows the number of studies conducted in or included participants from each country.

The systematic review identified barriers/enablers of BC sampling across 11 of 14 TDF domains. The most common domain reported was ‘knowledge’, followed by ‘behavioural regulation’, ‘environmental context and resources’, ‘memory, attention and decision processes’, ‘beliefs about consequences’ and ‘social influence’ (Table 1). Of 14 TDF domains, three and seven were not assessed in HICs and LMICs by the studies included in the systematic review, respectively (Appendix S9).

**Table 1:** Barriers and enablers of the 14 domains of the Theoretical Domains Framework (TDF) identified through the systematic review of the global evidence and the TDF survey in Indonesia, Thailand and Vietnam

| Theoretical Domains Framework (TDF) domains* | Systematic review of the global evidence |  |  |  | TDF survey in Indonesia, Thailand and Vietnam |  |
| --- | --- | --- | --- | --- | --- | --- |
|  | Frequency (N=study) | Barriers (N=study) | Enablers (N=study) | Mixed** (N=study) | Barriers | Enablers |
| Knowledge | 17 | 5 | 6 | 6 | Yes, very important | Yes, very important |
| Goals | 3 | 2 | 1 | 0 | Yes, very important | Yes, very important |
| Intentions | 2 | 1 | 0 | 1 | Yes, very important | Yes, very important |
| Social professional role and identity | 2 | 1 | 1 | 0 | Yes, very important | Yes, very important |
| Social influences | 6 | 1 | 5 | 0 | Yes, very important | Yes, very important |
| Environmental context and resources | 11 | 3 | 2 | 6 | Yes, very important | Yes, very important |
| Behavioural regulation | 15 | 1 | 6 | 6 | Yes, very important | Yes, very important |
| Reinforcement | Not evaluated | Not evaluated | Not evaluated | Not evaluated | Yes, very important | Yes, very important |
| Beliefs about consequences | 6 | 4 | 0 | 2 | Yes | Yes |
| Memory, attention and decision processes | 10 | 1 | 7 | 2 | Yes | Yes |
| Optimism | Not evaluated | Not evaluated | Not evaluated | Not evaluated | Yes | Yes |
| Skills | 1 | 0 | 0 | 1 | Yes | Yes |
| Beliefs about capabilities | 1 | 0 | 1 | 0 | Yes | Yes |
| Emotion | Not evaluated | Not evaluated | Not evaluated | Not evaluated | Yes | No |

\* TDF Domains are in rank order from most important to least important in Indonesia, Thailand and Vietnam. \*\*Mixed = Mixed barriers and enablers

#### TDF survey

From 1 December 2021 to 30 April 2022, 1,070 medical doctors and 238 final-year medical students in Indonesia, Thailand and Vietnam completed the online TDF survey covering all 14 TDF domains. Half of respondents were female (n=680, 52%) and most worked in governmental hospitals (n=980, 75.4%) (Table 2). The most common department was internal medicine (n=450, 34.4%), followed by emergency (n=175, 13.4%) and pediatrics (n=153, 11.7%). Respondents were from 24 of 34 provinces in Indonesia, 39 of 77 provinces in Thailand, and 25 of 63 provinces in Vietnam.

**Table 2:** Demographics and answer to the case scenario - the TDF survey

| Variables | Indonesia<br>(n=503) | Thailand<br>(n=304) | Vietnam<br>(n=501) | P<br>values |
| --- | --- | --- | --- | --- |
| **Female gender** | 263 (52.3%) | 195 (64.1%) | 222 (44.3%) | <0.001 |
| **Hospital types** |  |  |  |  |
| Government hospital | 340 (67.6%) | 209 (68.8%) | 431 (86.0%) | <0.001 |
| Private hospital | 113 (22.5%) | 15 (4.9%) | 17 (3.4%) |  |
| University hospital | 26 (5.2%) | 76 (25.0%) | 29 (5.8%) |  |
| Other<sup>1</sup> | 19 (3.8%) | 2 (0.7%) | 22 (4.4%) |  |
| I do not want to answer | 5 (1.0%) | 2 (0.7%) | 2 (0.4%) |  |
| **Hospital bed size** |  |  |  |  |
| <200 | 99 (19.7%) | 35 (11.5%) | 24 (4.8%) | <0.001 |
| 201-400 | 107 (21.3%) | 46 (15.1%) | 29 (5.8%) |  |
| 401-600 | 72 (14.3%) | 39 (12.8%) | 62 (12.4%) |  |
| 601-1,000 | 66 (13.1%) | 45 (14.8%) | 144 (28.7%) |  |
| 1,001-2,000 | 39 (7.8%) | 82 (27.0%) | 125 (25.0%) |  |
| >2,000 | 27 (5.4%) | 30 (9.9%) | 74 (14.8%) |  |
| I do not know | 89 (17.7%) | 27 (8.9%) | 35 (7.0%) |  |
| I do not want to answer | 4 (0.8%) | 0 (0%) | 8 (1.6%) |  |
| **Current job<sup>2</sup>** |  |  |  |  |
| Medical doctor – executive level | 13 (2.6%) | 5 (1.6%) | 17 (3.4%) | <0.001 |
| Medical doctor – consultant level | 74 (14.7%) | 75 (24.7%) | 198 (39.5%) |  |
| Medical doctor – physician level | 124 (24.7%) | 38 (12.5%) | 112 (22.4%) |  |
| Medical doctor – resident level | 168 (33.4%) | 63 (20.7%) | 101 (20.2%) |  |
| Medical doctor – intern level | 33 (6.6%) | 35 (11.5%) | 14 (2.8%) |  |
| Final-year medical student | 91 (18.1%) | 88 (28.9%) | 59 (11.8%) |  |
| **Department** |  |  |  |  |
| Internal medicine | 149 (29.6%) | 155 (51.0%) | 146 (29.1%) | <0.001 |
| Pediatrics | 65 (12.9%) | 43 (14.1%) | 45 (9.0%) | 0.05 |
| Infection disease division/department | 12 (2.4%) | 5 (1.6%) | 56 (11.2%) | <0.001 |
| Surgery | 21 (4.2%) | 45 (14.8%) | 81 (16.2%) | <0.001 |
| Orthopaedics | 6 (1.2%) | 18 (5.9%) | 14 (2.8%) | 0.001 |
| Obstetrics / Gynaecology | 20 (4.0%) | 29 (9.5%) | 7 (1.4%) | <0.001 |
| Emergency department | 112 (22.3%) | 34 (11.2%) | 29 (5.8%) | <0.001 |
| Intensive care unit | 45 (8.9%) | 13 (4.3%) | 51 (10.2%) | 0.01 |
| **Case-study: Would you take a BC sample in a case presenting with community-acquired sepsis?<sup>3,4</sup>** |  |  |  |  |
| Definitely (>95-100% of the time) | 157 (31.2%) | 273 (89.8%) | 252 (50.3%) | <0.001 |
| Likely (75-95% of the time) | 138 (27.4%) | 23 (7.6%) | 149 (29.7%) |  |
| Maybe (25-74% of the time) | 116 (23.1%) | 5 (1.6%) | 70 (14.0%) |  |
| Unlikely (5-24% of the time) | 44 (8.7%) | 2 (0.7%) | 19 (3.8%) |  |
| Rarely (ranging from never to <5% of the time) | 46 (9.1%) | 1 (0.3%) | 9 (1.8%) |  |
| I do not know | 1 (0.2%) | 0 (0%) | 1 (0.2%) |  |
| I do not want to answer | 1 (0.2%) | 0 (0%) | 1 (0.2%) |  |

See Appendix S4 for the questionnaire and Appendix S11 for the complete results of the TDF survey. <sup>1</sup> Included clinics (n=3) and text answers that could not be used to determine the hospital type such as internship and medical students. <sup>2</sup> In the survey, for a medical doctor, ‘executive level’ was defined as having an administrative position without clinical work, ‘consultant’ was defined as having a clinical specialty degree, ‘resident’ as currently under postgraduate clinical training, ‘physician’ as having no clinical specialty/subspecialty degree and not under postgraduate clinical training, and ‘intern’ as a recent medical school graduate in the first year of post-graduate on-the-job training. <sup>3</sup> In the survey, for the questions asking “Would you ...” or “How often ...”, the Likert scale options were defined as ‘definitely’ or ‘all the time’ (reflecting >95-100% of the time or case, respectively), ‘likely’
or 'often' (75-95%), 'uncertain' or 'moderately' (25-74%), 'unlikely' or 'occasionally' (5-24%), and 'rarely' (<5%). Each question also included 'I do not know' and 'I do not want to answer' options.<sup>4</sup> Case scenario. "A 72-year-old woman who was brought to the emergency department of your hospital by her daughter when she noticed the patient was more confused than her baseline and was found to have a high fever and fast breathing. She had an auscultatory finding compatible with pneumonia. It is decided that this patient will be admitted to your hospital." If you have an authority to take a blood culture, would you take blood culture sample(s) in this case on admission?

Based on the sepsis case scenario (Table 2), half of respondents (52.3%, 682/1,304) answered that they would definitely take BC. However, the responses were significantly different between the three countries (p<0.001). Most Thai respondents (89.8%, 273/304) answered that they would definitely take BC, while only half of Vietnamese respondents (50.5%, 252/499) and about a third of Indonesian respondents (31.3%, 157/501) did.

### Thematic synthesis for domains identified as having high importance

We combined the results obtained from the systematic review and the TDF survey (Table 1 and Appendix S10), and present content themes, in rank order, within each of the eight domains that were considered very important (i.e. key) in the three countries in SEA in the section below, with example references and quotes (Table 3).

**Table 3:**
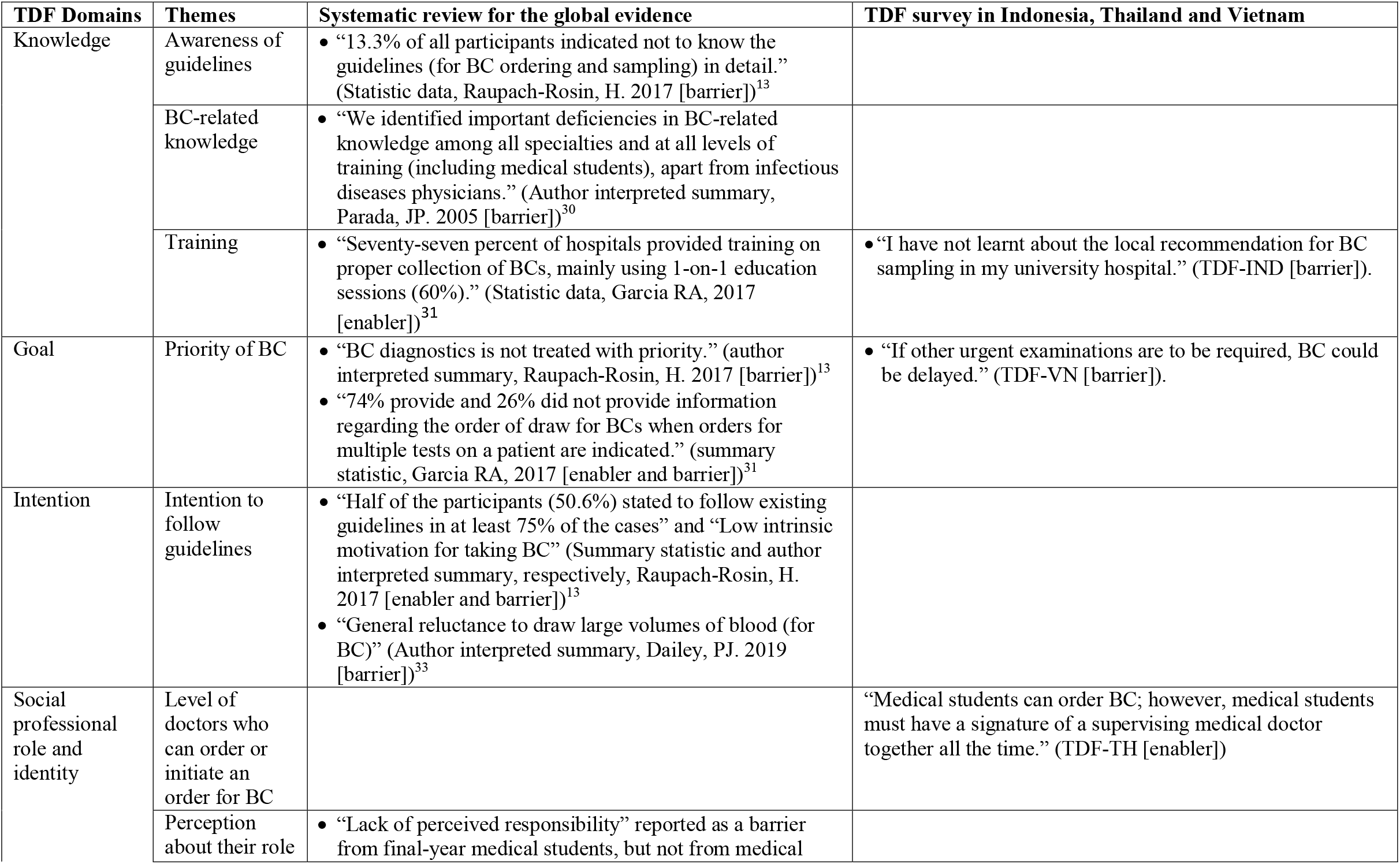
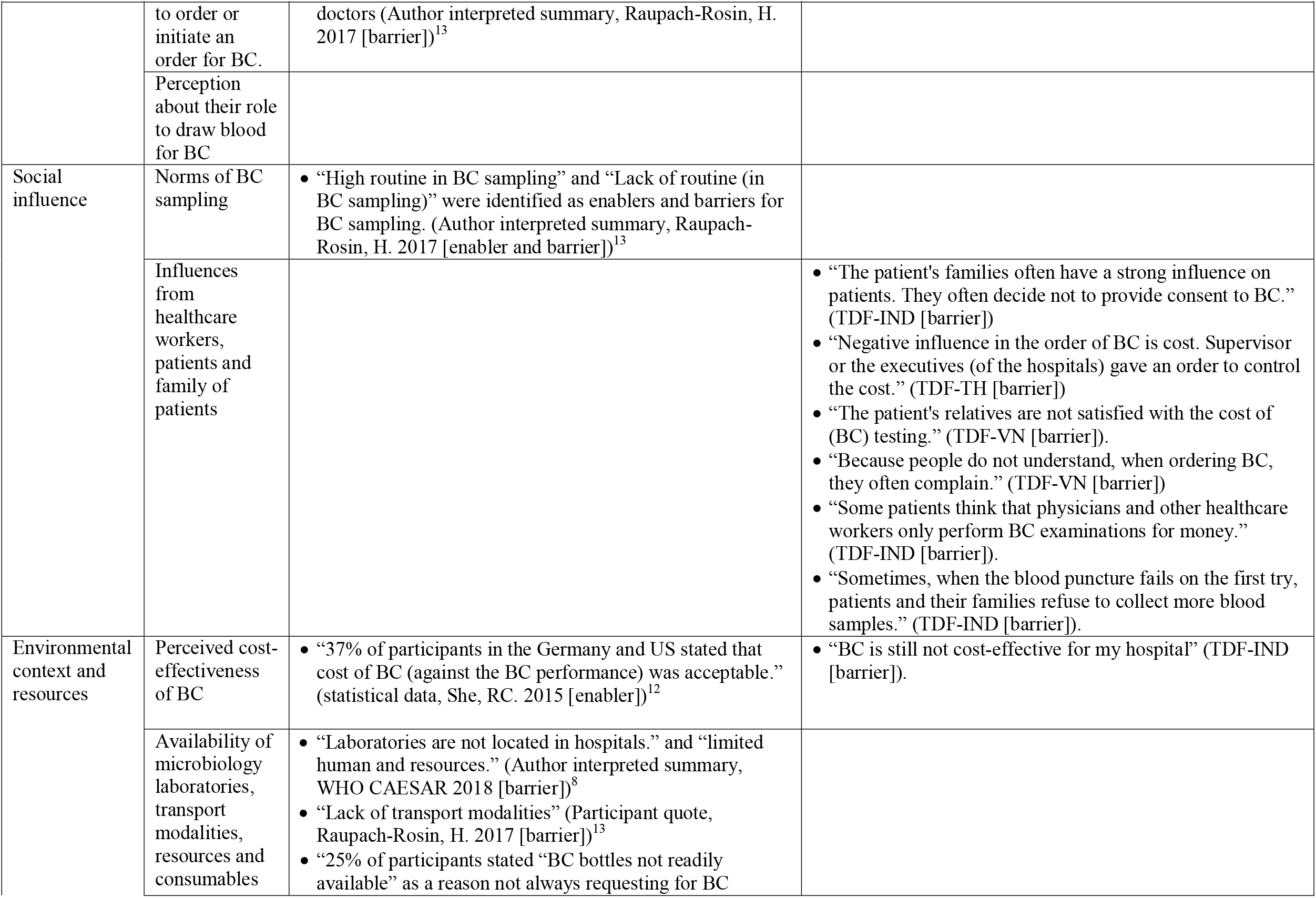
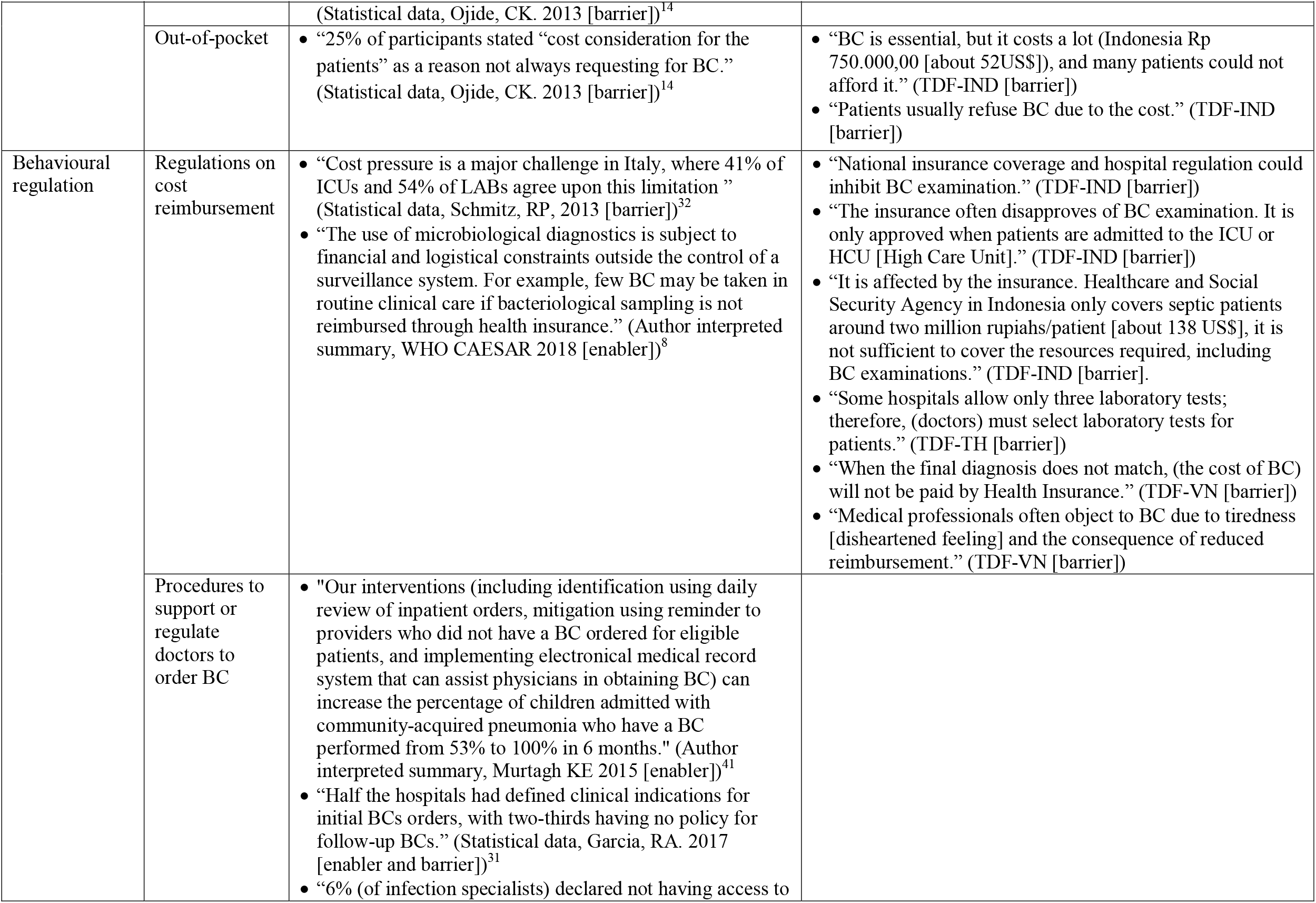
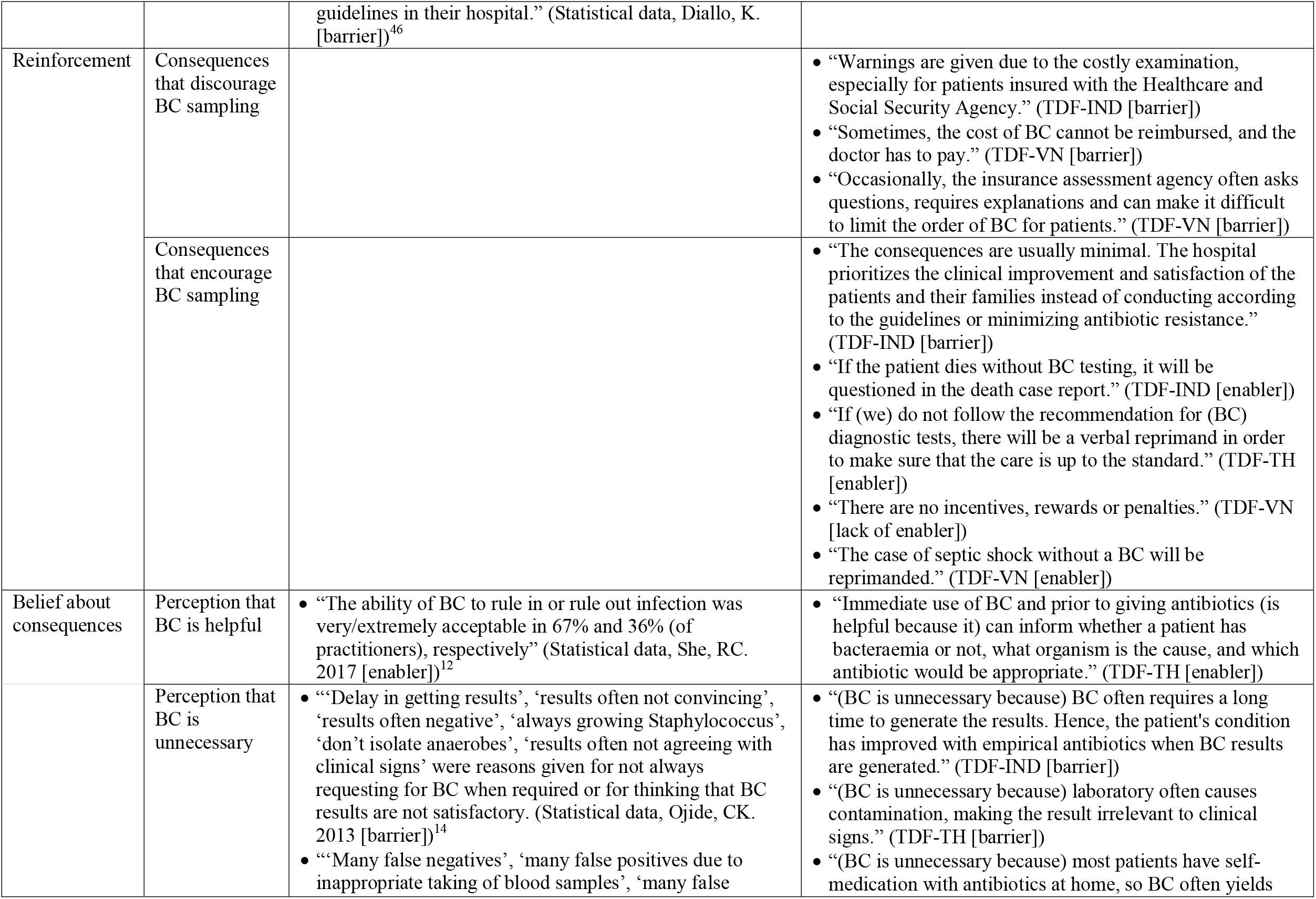
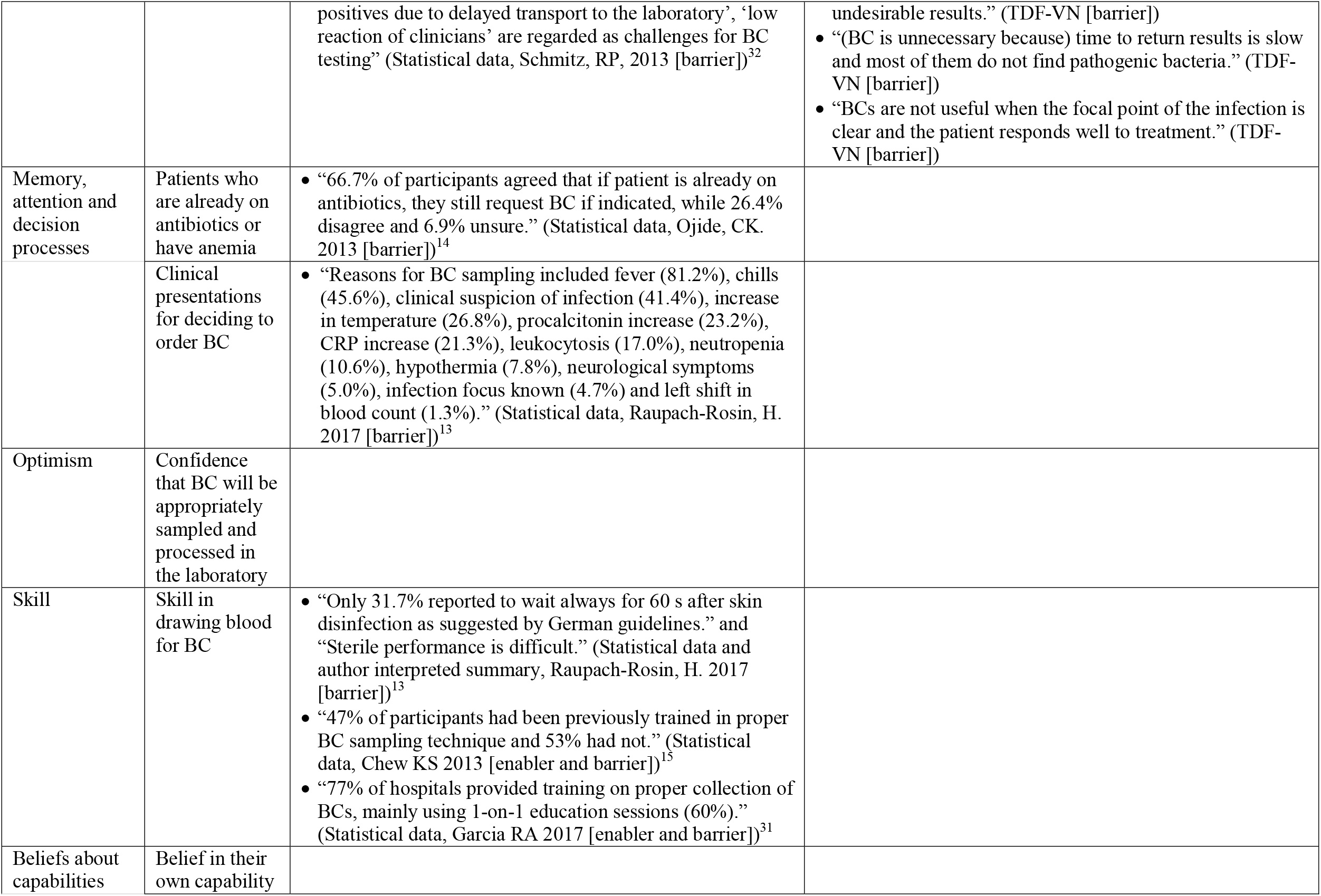
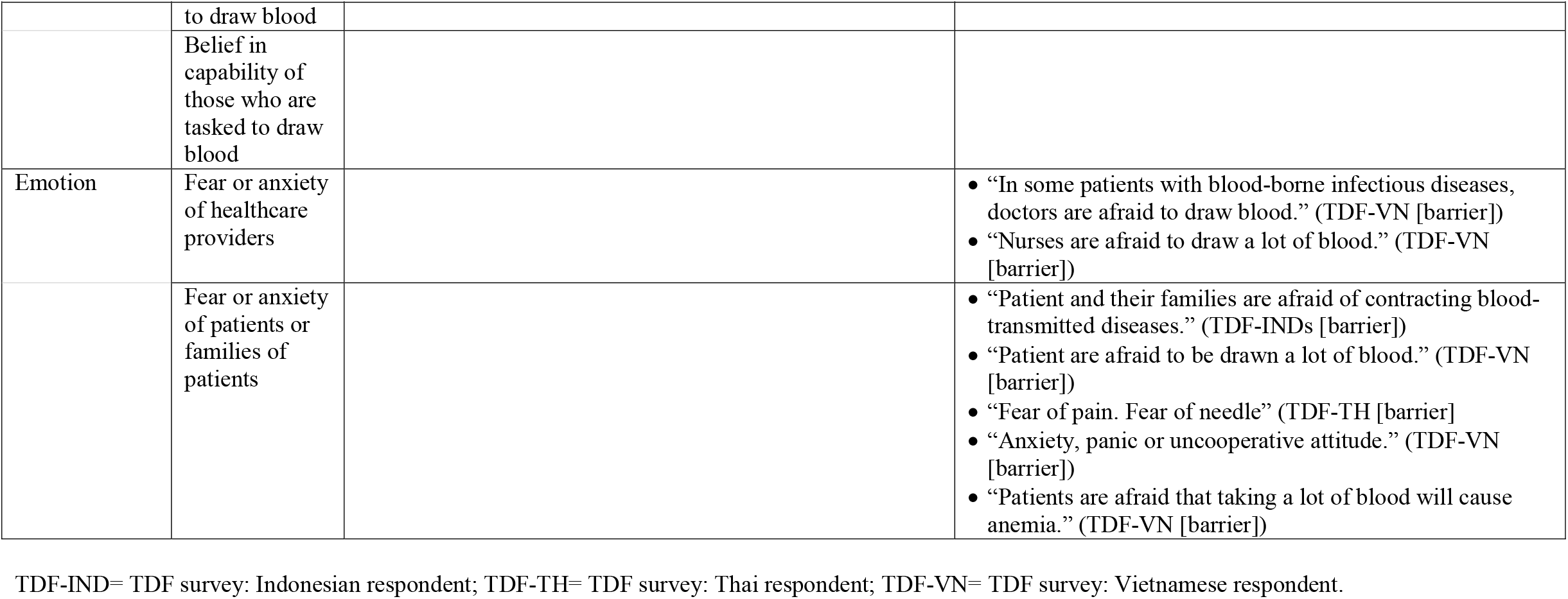
Sample quotes derived from the systematic review of the global evidence and the TDF survey in Indonesia, Thailand and Vietnam

#### Knowledge

##### Theme. Awareness of guidelines

In the review, “lack of knowledge of existing guidelines” was reported as a key barrier to BC sampling.^13^ In our survey, 59.0% (756/1,282) of respondents answered that they know of guidelines for BC sampling being used in their hospitals, while 29.3% (376/1,282) answered that they do not know if their hospitals use any guidelines, and 11.7% (150/1,282) answered that their hospitals do not use any guidelines (Appendix S11). Among those who answered that they know of local guidelines, 81.0% (612/756) stated that their local guidelines recommend BC sampling in all patients presenting with sepsis. About half (47.8%, 596/1,248) answered that they know of international guidelines, and most of those (95.0%, 566/596) stated that the international guidelines recommend BC sampling in all patients presenting with sepsis. Respondents who answered that they know of local guidelines (odds ratio [OR] 2.55, 95% confidence interval [CI] 1.93-3.38, p<0.001) or international guidelines (OR 1.97, 95%CI 1.50-2.57, p<0.001) were more likely to answer with “definitely take BC” in the case scenario (Appendix S12).

##### Theme: BC-related knowledge

In the review, previous studies reported deficiencies in BC-related knowledge, i.e. the number of BC bottles to be collected, amount of blood volume needed be collected, temperature of BC incubation, necessity of aseptic techniques and impact of antibiotics prior to BC collection.^12,15,30^ Our survey did not assess BC-related knowledge as this study did not focus on quality of BC practice.

##### Theme: Training

In the review ‘lack of training’ on BC was commonly reported as a barrier.^15,31,32^ In our survey, 37.8% (153/407) of Indonesian respondents, 24.9% (64/257) of Thai respondents and 12.4% (52/421) of Vietnamese respondents responded that there were no training, lectures, classes or meetings that provide knowledge about local/national/international guidelines for BC sampling in their hospitals (p<0.001). Respondents who answered that there are training, lectures, classes or meetings that provide knowledge about guidelines for BC sampling were more likely to answer with “definitely take BC” in the case scenario (OR 1.68; 95%CI 1.18-2.38, p=0.004).

#### Goals

##### Theme: Priority of BC

In the review, “low priority attributed to BC” was reported as a key barrier to BC sampling. However, “lack of time” or “time constraints” is commonly used to represent “low priority to BC” in the working environment with similar work density in hospitals.^13,32^ In many settings, ordering or initiating an order for BC can take only few seconds by writing “blood culture” in the doctor order form. In our survey, we used a question asking about the priority of BC compared to that of empirical antibiotics. 91.3% (274/300) of Thai respondents answered that they obtain BC prior to receiving empirical antibiotics in patients presenting with sepsis all the time or often, while 80.0% (380/475) of Vietnamese respondents and 54.2% (251/463) of Indonesian respondents answered likewise (p<0.001). Respondents who gave priority to BC were more likely to answer with “definitely take BC” in the case scenario (OR 4.25, 95%CI 3.04-5.94, p<0.001).

#### Intention

##### Theme: Intention to follow guidelines

In the review, “low intrinsic motivation”^13^ and “general reluctance to draw large volumes of blood”^33^ were reported as barriers to BC sampling. In our survey, among those who answered that they know of local guidelines, 92.9% (157/169) of Thai respondents answered that they plan to follow local guidelines all the time or often, while 82.0% (283/345) of Vietnamese respondents and 74.1% (172/232) of Indonesian respondents answered likewise. Those respondents were more likely to answer with “definitely take BC” in the case scenario (OR 2.92, 95% CI 1.88-4.53, p<0.001).

#### Social professional role and identity

##### Theme: Level of doctors who can order or initiate an order for BC

In the review, a survey evaluated the role of nurses to decide on BC testing in HICs,^32^ but none of the studies included evaluated the role of different levels of doctors on BC sampling. In our survey, >75% of Thai respondents answered that all levels of medical doctors (consultants, physicians, residents and interns) can order or initiate an order for BC in their hospitals. Most Indonesian and Vietnamese respondents (87.9%, 870/990) answered that consultants can, but fewer answered that physicians (61.8%, 612/990), residents (59.1%, 585/990) and interns (20.3%, 201/990) can (p<0.001). A quarter of Thai respondents (28.7%, 87/303) answered that final-year medical students can order or initiate an order for BC under supervision of attending medical doctors, while Indonesian respondents (2.2%, 11/500) and Vietnamese respondents (0.6%, 3/490) rarely answered likewise (p<0.001). None reported that nurses can order or initiate an order for BC.

##### Theme: Perception about their role to order or initiate an order for BC

In the review, ‘lack of perceived responsibility’ was reported as a barrier from final-year medical students, but not from medical doctors.^13^ In our survey, most medical doctors (86.5%, 905/1,046) answered that it is very appropriate or appropriate for them to order BC or initiate an order for BC, while only about half of final-year medical students (49.8%; 115/231) answered likewise (p<0.001). Those respondents were more likely to answer with “definitely take BC” in the case scenario (OR 3.36, 95%CI 2.50-4.51, p<0.001).

##### Theme: perception about their role to draw blood for BC

In our survey, most respondents (72.8%, 949/1,303) answered that registered nurses are tasked to draw blood from patients for BC, followed by microbiology laboratory team (36.0%, 469/1,303), specialized blood draw team (27.4%, 357/1,303), residents (25.4%, 331/1,303), physicians (23.5%, 306/1,303), consultants (23.2%, 302/1,303), interns (17.8%, 229/1,303) and final-year medical students (11.6%, 151/1,303). Of respondents who answered that they are tasked to draw blood from patients for BC, 69.1% (248/359) responded that it is very appropriate or appropriate for their role to draw blood for BC. Those respondents were more likely to answer with “definitely take BC” in the case scenario (OR 1.94, 95%CI 1.04-3.64, p=0.04).

#### Social influence

##### Theme: Norms of BC sampling

In the review, ‘lack of norms’ and ‘routine sampling’ were reported as important barriers^13^ and enablers^34,35^ to BC sampling. In our survey, 78.5% (233/297) of Thai respondents answered that they order BC because they are following local norms all the time or often, while 51.5% (238/462) of Vietnamese respondents and 43.8% (180/411) of Indonesian respondents answered likewise (p<0.001). Those respondents were more likely to answer with “definitely take BC” in the case scenario (OR 2.20, 95%CI 1.67-2.90, p<0.001).

##### Theme: Influences from healthcare workers, patients and family of patients

In the review, none of the studies included assessed the influences from other healthcare workers, patients or family of patients for BC sampling. In our survey, most respondents (79.4%) answered that there are very positive or positive influences on BC sampling from consultants, followed by residents (64.5%), doctors (64.6%), heads of department (65.9%), executive levels (50.6%), nurses (47.6%), interns (45.2%), patients (43.0%) and family of patients (31.9%). Some respondents said that there are negative or very negative influence in BC sampling from family of patients (6.8%), nurses (5.2%), patients (4.3%) and executives of the hospital (3.6%). Numerous quotes on this theme as a barrier were noted (Table 3).

#### Environmental context and resources

##### Theme: Perceived cost-effectiveness of BC

In the survey, ‘cost of BC’ or ‘economic pressure’ was reported as a barrier to BC sampling in both HICs^12,13,32,36^ and LMICs.^14^ In our survey, most (78.0%, 950/1,218) respondents considered that BC is very likely or likely to be cost-effective. Those respondents were more likely to answer with “definitely take BC” in the case scenario (OR 1.63, 95%CI 1.17-2.26, p<0.001).

##### Theme: Availability of microbiology laboratories, transport modalities, resources and consumables

In the review, unavailability of laboratories,^8^ logistical barriers,^8^ lack of BC bottles^14,37^ and limited human and resources^8^ were reported as barriers to BC sampling. In our survey, some respondents could not order BC because microbiology laboratories are not available or not functioning (13.4%, 157/1,174) or consumables (such as BC bottles, needles, syringes, blood collection set, etc.) are not available (12.7%, 150/1,181) all the time or often. Those respondents were not associated with BC practice in the case scenario (p>0.20 both)

##### Theme: Out-of-pocket

In the review, “cost consideration for the patients” was reported as a barrier to BC sampling.^14^ In the TDF survey, 23.3% (78/335) of Indonesian respondents answered that patients have to pay for BC using their own money (i.e. out of pocket) all the time or often, while 12.2% (28/230) of Thai participant and 8.3% (34/408) of Vietnamese participant answered likewise (p<0.001). Those respondents were not associated with BC practice in the case scenario (p=0.29)

#### Behavioural regulation

##### Theme: Regulation of cost reimbursement

In the review, “cost reimbursement” was reported as barriers to BC sampling.^8^ In our survey, 15.0% (196/1,308) and 11.6% (152/1,308) of respondents stated that ‘whether patients have a health scheme or insurance that covers the cost of BC’ and that ‘whether patients are likely to have a final diagnosis that includes the cost of BC in the package of fee for service’ are their additional reasons for deciding to order BC, respectively. Those respondents were not associated with BC practice in the case scenario (p>0.20, both). However, numerous quotes on this theme were noted (Table 3).

##### Theme: Procedures to support or regulate doctors to order BC

Interventional studies used development and dissemination of local guidelines, posters, notifications (i.e. feedback) and computer systems to support doctors to order BC per guidelines.^38-43^ In our survey, the most common procedures to support or regulate doctors to order BC in their hospitals was presence of local hospital guidelines (33.2%, 352/1,060), followed by case reviews (e.g. grand rounds or morning ward rounds, and BC is often mentioned) (30.8%, 326/1,060), standard order forms to remind ordering BC (29.9%, 317/1,060), stewardship programmes and reviewing BC is included in the programmes (19.5%, 207/1,060), posters (15.4%, 163/1,060) and computer systems to remind ordering BC (10.7%, 113/1,060). Respondents who answered that there are local hospital guidelines were more likely to answer with “definitely take BC” in the case scenario (OR 1.45; 95%CI 1.06-1.99, p=0.006).

#### Reinforcement

##### Theme: Consequences that discourage BC sampling

In the review, none of the studies included assessed reinforcement. In our survey, if they order a BC when it is recommended, some respondents (23.1%, 284/1,231) answered that there are either negative social consequences (e.g. verbal reprimand or any pressure from supervisors/executives of the hospital as the hospital (may) have to pay for the (extra) cost of BC) or negative material consequences (e.g. a negative score, that doctors are at risk of having to spend extra time and effort to reimburse the cost of BC from any health scheme or insurance, or that doctors are at risk of having to pay for the [extra] cost of BC themselves). Those who answered that there are negative consequences were less likely to answer with “definitely take BC” in the case scenario (OR 0.48; 95%CI 0.34-0.67, p<0.001).

##### Theme: Consequences that encourage BC sampling

In the survey, 23.7% (294/1,243) of respondents answered that there are either positive social (e.g. praise) or positive material (e.g. a positive score) consequences if they order a BC when it is recommended. Those respondents were less likely to answer with “definitely take BC” in the case scenario (OR 0.53; 95%CI 0.37-0.74, p<0.001). Nonetheless, we found that respondents who answered that there are positive consequences that encourage BC sampling when recommended were also answered that there are negative consequences that discourage BC sampling when recommended with moderate agreement beyond that expected by chance (Kappa value 0.46, p<0.001).

We also evaluated whether they are negative consequences if practitioners do not order a BC when it is recommended. 37.7% (464/1,230) of respondents answered that there are either negative social (e.g. verbal reprimand) or negative material (e.g. a negative score) consequences if they do not order a BC when it is recommended. Those respondents were not associated with answering with “definitely order BC” in the case scenario (p=0.42).

Additional results and the content themes in the domains that were identified as important in influencing in BC sampling are described in Appendix S1.

### Intervention types and policy options to improve BC sampling practice

We used the links between TDF and BCW, and listed all suggested intervention types and policy options related to very important TDF domains in Indonesia, Thailand and Vietnam (Table 4). A range of potential strategies were identified. Some strategies target individual reinforcement, environmental structure and social influence (e.g. providing an example for physicians to aspire to or imitate the BC sampling practice [modelling] and increasing means and reducing barriers to increase capability and opportunity to order BC for all levels of doctors [enablement]). Some strategies operate at the policy or service provision level (e.g. changing regulation of cost reimbursement [fiscal], development or implementation of local guidelines [guideline] and establishing rules or principles of BC practice [regulation]).

**Table 4.** Suggested intervention types and policy options to improve BC sampling practice based on very important TDF domains in Indonesia, Thailand and Vietnam

|  | COM-B components<sup>1</sup> |  |  |  |  |
| --- | --- | --- | --- | --- | --- |
|  | Psychological capability<br>(TDF: Know, Beh Reg) | Reflective motivation<br>(TDF: Goals, Int and Id) | Automatic motivation<br>(TDF: Reinf) | Physical opportunity<br>(TDF: Env) | Social opportunity<br>(TDF: Soc) |
| **Intervention types<sup>2</sup>** |  |  |  |  |  |
| Education | √ | √ |  |  |  |
| Persuasion |  | √ | √ |  |  |
| Incentivisation |  | √ | √ |  |  |
| Coercion |  | √ | √ |  |  |
| Training | √ |  |  |  |  |
| Restriction |  |  |  | √ | √ |
| Environmental restructuring |  |  | √ | √ | √ |
| Modelling |  |  | √ |  |  |
| Enablement | √ |  | √ | √ | √ |
| **Policy options<sup>2</sup>** |  |  |  |  |  |
| Communication/marketing |  | √ | √ |  |  |
| Guidelines | √ | √ | √ | √ | √ |
| Fiscal | √ | √ | √ | √ | √ |
| Regulation | √ | √ | √ | √ | √ |
| Legislation | √ | √ | √ | √ | √ |
| Environmental/social planning | √ |  | √ | √ | √ |
| Service provision | √ | √ | √ | √ | √ |

<sup>1</sup> COM-B component stands for Capability (Physical capability or Psychological capability), Opportunity (Physical opportunity or Social opportunity), Motivation (Automatic motivation or Reflective motivation)–Behaviour, represents source of the behaviours and is the core of the Behaviour Change Wheel (BCW). TDF = Theory domain framework. Know = Knowledge, Beh Reg = Behavioural regulation, Goals = Goals, Int = Intentions, Id = Social/professional role and identity, Reinf = Reinforcement, Env = Environmental context and resources, Soc = Social influences. <sup>2</sup> Suggested intervention types and policy options were identified using the links between TDF, the components of the COM-B and the BCW.<sup>16-18</sup>

## Discussion

Our review shows that the existing global evidence on barriers and enablers to BC sampling is limited, particularly for LMICs, and does not allow for drawing firm conclusions. Our survey in Indonesia, Thailand and Vietnam shows that ‘no awareness of guideline [TDF-knowledge]’, ‘low priority of BC [TDF-goals]’, ‘no intention to follow guidelines [TDF-intention]’, ‘level of doctors who can order or initiate an order for BC [TDF-social professional role and identity]’, ‘no norms of BC sampling [TDF-social influence]’, ‘perceived cost-effectiveness of BC [TDF-environmental context and resources]’, ‘regulation on cost reimbursement [TDF-behavioural regulation]’ and ‘consequences that discourage BC sampling [TDF-reinforcement]’ appeared to be key in influencing BC sampling. In Thailand,^10^ where BC utilization rate is relatively high compared to Indonesia^9^ and Vietnam,^11^ the frequencies of each barrier being reported by respondents is lower for most domains.

‘Low priority to BC [TDF-goals]’, ‘no intention to follow guidelines [TDF-intention]’, ‘lack of perception about their role to order or initiate an order for BC [TDF-social professional role and identity]’ and ‘lack of norms of BC sampling [TDF-social influence]’ are likely key barriers to BC sampling in both HICs^13,36,44^ and other LMICs^33^ where resources for BC sampling are available to some extent.

To our knowledge, ‘level of doctors who can order or initiate an order for BC [TDF-social professional role and identity]’ and ‘influence from healthcare workers, patients and families of patients [TDF-social influence]’ have never been systematically evaluated and reported as a barrier to BC sampling. Those are important barriers as, in many hospitals in both HICs and LMICs, final-year medical students and interns are responsible for most BC ordering and acquisition^30^ and influences from other parties can discourage BC sampling.

Remarkably, the cost of BC seems to have influence on executive level doctors, patients, families of patients, or those who set regulations on cost reimbursement of BC. This is shown by many quotes related to the cost of BC in the theme ‘influences from healthcare workers, patients and family of patients [TDF-social influence]’, ‘regulation on cost reimbursement [TDF-behavioural regulation]’ and ‘consequences that discourage BC sampling [TDF-reinforcement]’, respectively (Table 3). All stakeholders in each study country will need to consider all suggested intervention functions and policy options (e.g. providing clear posters emphasizing local guidelines of BC sampling over wide areas in hospitals to reduce negative influences from other parties on BC practice [environmental restructuring], changing regulation of cost reimbursement and finding financial support for BC sampling per local guidelines [fiscal], repeatedly announcing to all levels of healthcare workers that negative consequences that discourage BC sampling per local guidelines will not be tolerated [enablement] for cost-related barriers), and develop intervention content in details based on local context for the changes of BC practice to be successful.^16-18,45^

This study also has several limitations. First, the systematic review was limited in scope and quality due to limitation and heterogeneity of studies included. The TDF survey included a convenient sample of hospitals and practitioners, which might have led to selection bias. This limited our ability to draw firm conclusions on the contemporary situation on barriers and enablers to BC sampling globally and in the SEA region. Therefore, the data should be interpreted cautiously. Second, the exclusion of non-peer-reviewed and non-English literature meant that we might not have included local publications and documents from local government and/or non-government institutions. However, their exclusion likely improved the quality of the evidence as those literature may not always follow gold-standard or recommended guidelines for evaluation. Third, the survey could not reach the target sample size in Thailand despite substantial efforts. Nonetheless, the study had enough power to estimate the prevalence of barriers and enablers in Thailand, and compare that with those observed in Indonesia and Vietnam.

In conclusion, among three LMICs in SEA barriers to BC sampling were varied and heterogeneous. This comprehensive analysis using TDF gives information across the entire spectrum of behavioral influences of BC sampling. These results can help local healthcare providers and policy makers to develop and implement interventions aiming to improve diagnostic stewardship practices.

## Data Availability

All produced in the present study are available upon reasonable request to the corresponding author.

## Declaration of interests

The authors declare no competing financial interests.

## Data sharing

The study protocol for the systematic review, with all relevant tools, is publicly available in PROSPERO, CRD42020206557. All produced in the present study are available upon reasonable request to the corresponding author.

## Acknowledgments

The authors thank the respondents and staff at all the study hospitals and Mahidol-Oxford Tropical Medicine Research Unit. The authors thank Surveillance and Epidemiology of Drug-resistant Infections Consortium (SEDRIC) for the support and comments on the study. We thank Le Nguyen Minh Hoa, Vu Minh Duy Pharm, Dam Thi Hong Hanh and Hoang Bao Long for support in data collection. This research was funded by the Wellcome Trust (220557/Z/20/Z). For the purpose of Open Access, the author has applied a CC BY public copyright licence to any Author Accepted Manuscript version arising from this submission.

## Author contributions

F.L., L.A. and D.L. designed and supervised the study. P.S., K.S.A., R.L., V.T.L.H., H.R.v.D. and R.L.H. participated in project design and facilitated data collection. A.T., L.W.A.R., R.B., E.J.N., D.U.N., S.K., W.S., P.C., W.P., N.H.Y., P.N.T., L.M.Q., V.H.V., C.M.D., V.T.H.D.E. and E.H. facilitated data collection. P.S. analyzed the data and wrote the first draft of the manuscript. All authors contributed to the writing or revision of the manuscript. P.S. and D.L. verified the data.

## Notes

### Competing Interest Statement

The authors have declared no competing interest.

### Clinical Protocols

https://www.crd.york.ac.uk/prospero/display_record.php?ID=CRD42020206557

### Author Declarations

The study was approved by the Oxford University Tropical Research Ethics Committee (OXTREC545-21) and local ethical committees at Iskak Tulungagung Hospital (070/7303/407.206/2021), Prof. Dr. R.D. Kandou Hospital (156/EC/KEPK-KANDOU/IX/2021), Pasar Minggu Hospital (EOCRU/RCH.216/10.2021/1145) in Indonesia, The National Hospital for Tropical Diseases (14HDDD/NDTU) in Vietnam, Faculty of Tropical Medicine, Mahidol University (TMEC21-069), Sunpasitthiprasong Hospital (065/64S) and Chiangrai Prachanukroh Hospital (CR 0032.102/EC023) in Thailand.

