## Appendix for "Barriers and enablers to blood culture sampling: a systematic review and theoretical domains framework survey in Indonesia, Thailand and Vietnam"

**Table of contents**

|  | **Page** |
| --- | --- |
| Appendix S1: Supplementary Text | 2 |
| Supplementary Methods | 2 |
| Supplementary Results | 4 |
| Supplementary Discussions | 8 |
| Appendix S2. Search strategies for the systematic review | 10 |
| Appendix S3. Theoretical Domains Framework: Definitions and examples | 14 |
| Appendix S4. TDF survey questionnaire | 17 |
| Appendix S5. PRISMA checklist | 34 |
| Appendix S6. Characteristics of studies included in the systematic review | 37 |
| Appendix S7. Study methods and locations of studies included in the systematic review | 43 |
| Appendix S8. Quality assessment of studies included in the systematic review | 44 |
| Appendix S9. Barriers and enablers of the 14 TDF domains identified in HICs and LMICs derived from the systematic review | 45 |
| Appendix S10. Criteria and rank of TDF domains for Indonesia, Thailand and Vietnam | 46 |
| Appendix S11. Results of the TDF survey in Indonesia, Thailand and Vietnam | 47 |
| Appendix S12. Associations between barriers or enablers and the responses that they would definitely take BC in the case scenario in the TDF survey | 64 |
| Appendix S13. Links between TDF, COM-B components (Capability, Opportunity, motivation and behaviour components), and suggested intervention types and policies | 71 |
| Appendix S14. References | 73 |

**Appendix S1: Supplementary Text**

**Supplementary Methods**

**A systematic review**

“Lack of time” was classified as a barrier in the domain “Goal” as the statement represents the low priority attributed to BC diagnostics since time is relative.^1^ Reported barriers or enablers stratified by professional roles were categorized to the domain of the barriers or enablers rather than the domain “professional role and identity”. For example, “all specialties and at all levels of training (including medical students), apart from infectious diseases physicians, lack important deficiencies in blood culture (BC)-related knowledge.^2^” (author interpreted summary, Parada JP et al)^2^ was categorized to the domain “knowledge”.

We categorised the findings according to the 14 TDF domains based on information presented in the publications. For example, ‘norms of BC sampling’^1^ could be categorized to domain ‘social influence’ or ‘behaviour regulation’ based on detailed information.^3-5^ We categorized the reported ‘norms of BC sampling’^1^ to domain ‘social influence’ based on limited information presented in the publications.

**TDF survey**

We conducted a TDF survey to comprehensively assess the reported and potential barriers and enablers in three countries in Southeast Asia (SEA), i.e. Indonesia, Thailand and Vietnam. The three countries were selected based on the study team’s long history of working and collaborating with hospitals and ministry of health in the three countries.

In 2020, Indonesia (GDP per capita: 3,869.6 US$) and Vietnam (GDP per capita: 2,785.7 US$) were a lower-middle-income country and Thailand (GDP per capita:7,186.9 US$) was an upper-middle-income country in SEA.^6^

The healthcare systems in SEA are highly diverse.^7^ Indonesia has a decentralised public healthcare system, in which provincial or district-level governments have the authority over most public hospitals, and a substantial private health sector. To achieve the goal of universal healthcare coverage (UHC), in 2014 the Government introduced national health insurance (Jaminan Kesehatan Nasional), which had reached 84% of the population by 2021. Thailand achieved the status of UHC in 2002 in terms of insurance entitlement, when the gross national income per capita was 1,900 US$.^8^ It is shown that UHC in Thailand can improve quality of care without undermining the efficiency and equity of the policy.^9^ Vietnam has implemented social health insurance (SHI) since 1992, and SHI had a role as a financial mechanism towards achieving UHC,^10^ which had reached 82% of the population in 2018. The benefit package of universal SHI in Vietnam is considered generous, particularly regarding the drugs subsidized.^10^ However, out-of-pocket payments are still high.^10,11^ In 2019, percentages of out-of-pocket expenditure among all health expenditure were 35%, 9% and 43% in Indonesia, Thailand and Vietnam, respectively.^12^

We used a simple formula for calculating sample sizes for descriptive studies.^13^ Assuming prevalence of a barrier or enabler to be 50% among medical doctors, with a margin of error 5%, the sample size of medical doctors was estimated to be at least 385 per country. Assuming prevalence of a barrier or enabler to be 50% among final-year medical students, with a margin of error 10%, the sample size of final-year medical students was estimated to be at least 97 per country. Therefore, we aimed to enroll 400 medical doctors and 100 final-year medical students in each country (a total of 1,500 respondents).

During the pilot survey, we included ‘monetary reward’ and ‘monetary fine’ as examples of positive and negative consequences to BC sampling, respectively. We received strong feedbacks that those are not present for BC sampling in Indonesia, Thailand and Vietnam. Therefore, the word ‘monetary reward’ and ‘monetary fine’ were removed.

**Analysis**

We decided not to conduct multivariable logistic regressions because we considered that each TDF domain could influence BC sampling practice via a causal relationship. The degree of agreement between two questions was estimated using the Kappa index. This describes the level of association, both positive and negative, beyond that caused by chance, as follows: 0.00–0.20, slight; 0.21–0.40, fair; 0.41–0.60, moderate; 0.61–0.80, substantial; 0.81–1.00, high.

**Additional analysis**

We explored whether the answers of respondents who completed the survey were different from the answers of respondents who did not complete the survey. We compared the answers to the case scenario between those who completed the questionnaire and those who answered the case scenario (Question 1-3 in the questionnaire) but did not complete the questionnaire. Logistic regression model with random effects for countries was used for the analysis.

**Supplementary Results**

Additional results and the content themes in the domains that were identified as important in influencing in BC sampling are described in further detail in the sections below.

***Belief about consequences***

*Theme: Perceived that BC is helpful.* In the review, BC’s ability to rule in an infection and to exclude infections were reported as enablers to BC sampling.^14^ In our survey, most respondents strongly agreed or agreed that BC is helpful in adjusting antibiotics (94.0%, 1,224/1,302), clinical decisions (93.6%, 1,220/1,303), detecting AMR bacterial infections (92.1%, 1,199/1,302) and ruling in an infection (90.2%, 1,172/1,299), reducing overuse of antibiotics (87.4%, 1,140/1,304), reducing patient mortality (79.2%, 1,027/1,297). 72.3% (938/1,298) of respondents strongly agreed or agreed that BC is helpful in reducing length of hospital stay. 60.5% (786/1,300) of respondents strongly agreed or agreed that BC is helpful in ruling out an infection. Most respondents strongly agreed or agreed that accumulative results of BC are helpful in understanding epidemiology of AMR bacterial infections (94.5%, 1,228/1,299). Respondents who answered that BC is helpful in clinical decisions (OR 2.96, 95%CI 1.71-5.12, p<0.001), to reduce patient mortality (OR 1.61; 95%CI 1.18-2.20, p=0.003), to rule in an infection (OR 1.58, 95%CI 1.04-2.39, p=0.03), or to reduce length of hospital stay (OR 1.53, 95%CI, 1.14-2.04, p=0.004) and those who answered that accumulative results of BC are helping in understanding epidemiology of AMR bacterial infections (OR 2.89, 95%CI 1.60-5.19, p<0.001) were more likely to answer with “definitely take BC” in the case scenario. Respondents who answered that BC is helpful to rule out an infection, to detecting AMR bacterial infection, in adjusting antibiotics, or reduce overuse of antibiotics were not associated with BC practice in the case scenario (p>0.10, all).

*Theme: Perceived that BC is unnecessary.* In the review, ‘BC results often negative’, ‘many false negatives and false positives’, ‘always growing Staphylococcus’, ‘results often not convincing’, ‘lack of trust in BC results’, ‘lack of clinical utility’, ‘delay in getting results’, ‘don’t isolate anaerobes’, ‘results often not agreeing with clinical signs’ and ‘not a requirement for treating every case’ were reported as barriers of BC sampling.^1,15-18^ In our survey, some respondents strongly agreed or agreed that BC is unnecessary because it is not too late to collect BC later, particularly if patients do not improve after receiving empirical antibiotic treatment (32.7%, 423/1,293), the therapeutic consequence of BC sampling is questionable (18.6%, 238/1,277), antibiotic therapy can be determined based on clinical presentations (17.5%, 228/1,301), results are often delayed (17.0%, 220/1,298) quality of laboratory is questionable (15.3%, 194/1,269), the scientific basis of the guideline on BC is questionable (15.0%, 191/1,277), results are often negative or no growth (11.4%, 148/1,295), and results are often contaminated (11.1%, 143/1,288). Respondents who answered that BC is unnecessary because antibiotic therapy can be determined based on clinical presentation (OR 0.51, 95%CI 0.36-0.73, p<0.001), the scientific basis of the guideline on BC is questionable (OR 0.66, 95%CI 0.45-0.98, p=0.04), results are often delayed (OR 0.48, 95%CI 0.33-0.69, p<0.001), results are often not interpretable (OR 0.54, 95%CI 0.34-0.87, p=0.01), results are often negative or no growth (OR 0.58, 95%CI 0.39-0.88, p=0.01), cultures are often contaminated (OR 0.64, 95%CI 0.42-0.98, p=0.04), BC is not benefiting the patients (OR 0.37; 95%CI 0.24-0.57, p<0.001), a contaminated result often leads to wrong therapeutic approach (OR 0.53; 95%CI 0.30-0.95, p=0.03), it is not too late to collect BC later, particularly if patients do not improve after receiving empirical antibiotic treatment (OR 0.37; 95%CI 0.27-0.52, p<0.001), quality of laboratory is questionable (OR 0.48; 95%CI 0.33-0.70, p<0.001) or levels of local antibiotic resistance are low (OR 0.64; 95%CI 0.41-0.98, p=0.04) were less likely to answer with “definitely take BC” in the case scenario. Respondents who answered that BC is unnecessary because the therapeutic consequence of BC is questionable, or results often do not agree with clinical signs were not associated with BC practice in the case scenario (p>0.20, both).

***Memory, attention and decision processes***

*Theme: Patients who are already on antibiotics or have anemia.* In the review, ‘ if patient is already on antibiotics’ was reported as a barrier to BC sampling when indicated.^15^ In our survey, 10.2% (131/1,287) of respondents stated that they will definite or likely not order BC when patients are already on antibiotics even if BC is recommended. Those respondents were not associated with BC practice in the case scenario (p=0.13). 22.3% (280/1,258) of respondents stated that they will definite or likely not order BC when patients have anemia even if BC is recommended. Those respondents were not associated with BC practice in the case scenario (p=0.55).

*Theme: Clinical presentations for deciding to order BC.* In the review, reported reasons for BC sampling included fever, chills, clinical suspicion of infection, increase in temperature, procalcitonin increase, CRP increase, leukocytosis, neutropenia, hypothermia, neurological symptoms, infection focus known and left shift in blood count.^1^ A survey study in the Germany and US found that respondents perceived febrile neutropenic patients, patients in critical/intensive care and those with indwelling devices are likely to have BSI and BC sampling.^14^ In our survey, among respondents who responded that they know of local guidelines, some stated that patients with no clinical improvement after receiving empirical antibiotics (36.2%, 274/756), presenting with fever of unknown origin (30.6%, 231/756), suspected of hospital-acquired infection (30.8%, 233/756), presenting with chronic fever (28.6%, 216/756) or suspected of infection caused by antimicrobial-resistant organisms (28.6%, 216/756) are their additional reasons to order BC.

In the review, reported reasons for sending anaerobic BC were clinical concerns with necrotizing enterocolitis, intraperitoneal infection, bowel rupture, recent surgery, a history of wound infection, clinical suspicion and immunosuppressed patients, and a reported reason to not obtain anaerobic BC was a concern regarding excessive blood volume.^19,20^ Reported reasons for follow-up BC sampling were persistent fever, follow-up of positive BC, new fever, leukocytosis and haemodynamic changes.^21,22^ Our survey did not evaluate barriers/enablers to anaerobic BC or follow-up BC sampling.

***Optimism***

*Theme: Optimism about the laboratory.* In the review, no studies included assessed optimism as a barrier/enabler to BC sampling. In our survey, most (80.5%, 1,034/1,285) respondents answered that they are strongly optimistic or optimistic that a BC will be sampled and processed in the laboratory appropriately if they order a BC. Respondents who were strongly optimistic or optimistic about the laboratory were more likely to answer with “definitely take BC” in the case scenario (OR 1.78, 95%CI 1.29-2.46, p<0.001)

***Skills***

*Theme: Skills in drawing blood for BC.* In the review, “sterile performance is difficulty” was reported as a barrier.^1^ In our study, among respondents whom were tasked to draw blood from patients for BC in their hospitals, 44.1% (143/324) answered that their skill of drawing blood from patients for BC is very good or good, 44.8% (145/324) fair, and 11.1% (36/324) poor or very poor. Respondents who answer that they have very good or good skill in drawing blood for BC was more likely to answer with “definitely take BC” in the case scenario (OR 1.74; 95%CI 1.02-2.07, p=0.04).

***Belief about capabilities***

*Theme: Belief in their own capability to draw blood*. In the review, ‘belief in capability to interpret microbiology results’ was reported as an enabler;^23^ however, no studies assessed belief in capability to draw blood for BC. In our survey, 73.9% (244/358) of respondents answered that they are strongly confident or confident that they can draw BC successfully. Respondents who answered that they are strongly confident or confident that they can draw BC successfully were not associated with BC practice in the case scenario (p=0.36). 74.8% (246/329) answered that they are strongly confident or confident that they can draw BC appropriately using aseptic technique. Respondents who answered that they are strongly confident or confident that they can draw BC appropriately using aseptic technique were not associated with BC practice in the case scenario (p=0.11).

*Theme: Belief in capability of those who are tasked to draw blood.* In our survey, 88.5% (1,151/1,300) of respondents answered that they are strongly confident or confident that those who are tasked to draw BC can draw BC successfully. Those respondents were not associated with BC practice in the case scenario (p=0.13). 76.7% (996/1,298) of respondents answered that they are strongly confident or confident that those who are tasked to draw BC can draw BC appropriately using aseptic technique. Those respondents were not associated with BC practice in the case scenario (p=0.23).

***Emotion***

*Theme: Fear or anxiety of healthcare providers* and *Fear or anxiety of patients or family of patients.* In the review, no studies included assessed emotion as a barrier/enabler to BC sampling. In our survey, some (7.1%, 93/1,308) respondents stated that there are emotional factors associated with ordering BC. Those include fear or anxiety related to pain, needles, blood-borne diseases, and high volume of blood (Table 3). Those respondents who answered that there are emotional factors associated with ordering BC were not associated with BC practice in the case scenario (p=0.82).

**Additional analysis**

We explored whether there was any evidence showing a difference between respondents who completed and did not complete the survey. Of 2,095 respondents who agreed to participate the online survey, 1,308 (62.4%) completed the questionnaire, 256 (12.2%) answered the question about the case-study (Question 1-3) but did not completed the questionnaire, and 531 (25.3%) did not answer the question about the case-study. The proportion of patients who answered that they would definitely take BC for the case scenario was not different between those who completed the questionnaire (52.1%; 682/1,308) and those who answered the question about the case scenario but did not complete the questionnaire (51.2%; 131/256) (p=0.08).

**Supplementary Discussions**

A fear of ‘blood stealing’ or ‘blood selling’ was reported as a barrier to blood specimen collection in sub-Saharan Africa.^24^ The study was not included in the systematic review because the study was not specific to BC sampling. In our pilot survey and the TDF survey, we did not observe the fear of ‘blood stealing’ or ‘blood selling’ in Indonesia, Thailand and Vietnam. It is likely that emotional barriers to BC sampling are different depending on local regions.

**Appendix S2: Search strategies for the systematic review**

Scoping searches were undertaken on the following websites and databases (list in alphabetical order) on 02/10/2020 and EBSCO CINAHL search strategy (run 01/12/2020) by Eli Harriss, a librarian at the Bodleian Health Care Libraries, University of Oxford.

**Cochrane Database of Systematic Reviews Issue 10 of 12, October 2020**

**Cochrane Central Register of Controlled Trials Issue 10 of 12, October 2020**

1. #1	MeSH descriptor: [Blood Culture] explode all trees	11

2. #2	"blood cultur*"	154

3. #3	"diagnostic stewardship*"	0

4. #4	((diagno* or detect*) and ("bloodstream infec*" or bacteraemia* or bacteremia*))	837

5. #5	#1 or #2 or #3 or #4	939

6. #6	knowledge or attitude* or practice* or perception* or KAP or belief* or motivation* or barrier* or enable* or facilitate* or obstacle* or behavio* or social* or order* or skill* or competen* or abilit* or memory* or decision* or regulation* or professional* or role* or capabilit* or environment* or resource* or survey* or routine* or guideline* or limit* or identity or optimis* or consequence* or reinforcement or intention* or goal* or memory or attention or context* or resource* or emotion*	669746

7. #7	#5 and #6	618

**EBSCO CINAHL search strategy (run 01/12/2020)**

1. S1 (MH "Blood Culture")

2. S2 TI "blood cultur*"

3. S3 TI "diagnostic stewardship*"

4. S4 TI ((diagno* or detect*) and ("bloodstream infec*" or bacteraemia* or bacteremia*))

5. S5 S1 OR S2 OR S3 OR S4

6. S6 TI ( knowledge or attitude* or practice* or perception* or KAP or belief* or motivation* or barrier* or enable* or facilitate* or obstacle* or behavio* or social* or order* or skill* or competen* or abilit* or memory* or decision* or regulation* or professional* or role* or capabilit* or environment* or resource* or survey* or routine* or guideline* or limit* or identity or optimis* or consequence* or reinforcement or intention* or goal* or memory or attention or context* or resource* or emotion* ) OR AB ( knowledge or attitude* or practice* or perception* or KAP or belief* or motivation* or barrier* or enable* or facilitate* or obstacle* or behavio* or social* or order* or skill* or competen* or abilit* or memory* or decision* or regulation* or professional* or role* or capabilit* or environment* or resource* or survey* or routine* or guideline* or limit* or identity or optimis* or consequence* or reinforcement or intention* or goal* or memory or attention or context* or resource* or emotion* )

7. S7 S5 AND S6

8. S8 S5 AND S6 Expanders - Apply equivalent subjects Narrow by Language: - english

**Database: Embase 1974 to present**

Search Strategy:

1. *blood culture/ (4420)

2. "blood cultur*".ti. (5372)

3. "diagnostic stewardship*".ti. (69)

4. ((diagno* or detect*) and ("bloodstream infec*" or bacteraemia* or bacteremia*)).ti. (1024)

5. 1 or 2 or 3 or 4 (7172)

6. (knowledge or attitude* or practice* or perception* or KAP or belief* or motivation* or barrier* or enable* or facilitate* or obstacle* or behavio* or social* or order* or skill* or competen* or abilit* or memory* or decision* or regulation* or professional* or role* or capabilit* or environment* or resource* or survey* or routine* or guideline* or limit* or identity or optimis* or consequence* or reinforcement or intention* or goal* or memory or attention or context* or resource* or emotion*).ti,ab. (14220815)

7. 5 and 6 (2692)

8. 7 (2692)

9. limit 8 to english language (2510)

**ProQuest Dissertations & Theses Global**

noft(("blood cultur*" OR "diagnostic stewardship*" OR ((diagno* OR detect*) AND ("bloodstream infec*" OR bacteraemia* OR bacteremia*)))) AND noft(knowledge or attitude* or practice* or perception* or KAP or belief* or motivation* or barrier* or enable* or facilitate* or obstacle* or behavio* or social* or order* or skill* or competen* or abilit* or memory* or decision* or regulation* or professional* or role* or capabilit* or environment* or resource* or survey* or routine* or guideline* or limit* or identity or optimis* or consequence* or reinforcement or intention* or goal* or memory or attention or context* or resource* or emotion*)

Applied filters: English

**Database: PsycINFO 1806 to present**

Search Strategy:

1. "blood cultur*".tw. (114)

2. "diagnostic stewardship*".tw. (0)

3. ((diagno* or detect*) and ("bloodstream infec*" or bacteraemia* or bacteremia*)).tw. (38)

4. 1 or 2 or 3 (146)

5. (knowledge or attitude* or practice* or perception* or KAP or belief* or motivation* or barrier* or enable* or facilitate* or obstacle* or behavio* or social* or order* or skill* or competen* or abilit* or memory* or decision* or regulation* or professional* or role* or capabilit* or environment* or resource* or survey* or routine* or guideline* or limit* or identity or optimis* or consequence* or reinforcement or intention* or goal* or memory or attention or context* or resource* or emotion*).tw. (3724276)

6. 4 and 5 (80)

7. 6 (80)

8. limit 7 to english language (77)

**PubMed**

(("Blood Culture"[Mesh] or "blood cultur*"[Title] or "diagnostic stewardship*"[Title]) or ((diagno*[Title] or detect*[Title]) and ("bloodstream infec*" [Title] or bacteraemia*[Title] or bacteremia*[Title]))) AND (knowledge[Title/Abstract] OR attitude*[Title/Abstract] OR practice*[Title/Abstract] OR perception*[Title/Abstract] OR KAP[Title/Abstract] OR belief*[Title/Abstract] OR motivation*[Title/Abstract] OR barrier*[Title/Abstract] OR enable*[Title/Abstract] OR facilitate*[Title/Abstract] OR obstacle*[Title/Abstract] OR behavio*[Title/Abstract] OR social*[Title/Abstract] OR order*[Title/Abstract] OR skill*[Title/Abstract] OR competen*[Title/Abstract] OR abilit*[Title/Abstract] OR memory*[Title/Abstract] OR decision*[Title/Abstract] OR regulation*[Title/Abstract] OR professional*[Title/Abstract] OR role*[Title/Abstract] OR capabilit*[Title/Abstract] OR environment*[Title/Abstract] OR resource*[Title/Abstract] OR survey*[Title/Abstract] OR routine*[Title/Abstract] OR guideline*[Title/Abstract] OR limit*[Title/Abstract] OR identity[Title/Abstract] OR optimis*[Title/Abstract] OR consequence*[Title/Abstract] OR reinforcement[Title/Abstract] OR intention*[Title/Abstract] OR goal*[Title/Abstract] OR memory[Title/Abstract] OR attention[Title/Abstract] OR context*[Title/Abstract] OR resource*[Title/Abstract] OR emotion*[Title/Abstract]) Filters applied: English.

**Scopus**

( ( TITLE ( "blood cultur*" ) OR TITLE ( "diagnostic stewardship*" ) OR TITLE ( ( ( diagno* OR detect* ) AND ( "bloodstream infec*" OR bacteraemia* OR bacteremia* ) ) ) ) ) AND ( ( TITLE ( knowledge OR attitude* OR practice* OR perception* OR kap OR belief* OR motivation* OR barrier* OR enable* OR facilitate* OR obstacle* OR behavio* OR social* OR order* OR skill* OR competen* OR abilit* OR memory* OR decision* OR regulation* OR professional ) OR TITLE ( role* OR capabilit* OR environment* OR resource* OR survey* OR routine* OR guideline* OR limit* OR identity OR optimis* OR consequence* OR reinforcement OR intention* OR goal* OR memory OR attention OR context* OR resource* OR emotion* ) OR ABS ( knowledge OR attitude* OR practice* OR perception* OR kap OR belief* OR motivation* OR barrier* OR enable* OR facilitate* OR obstacle* OR behavio* OR social* OR order* OR skill* OR competen* OR abilit* OR memory* OR decision* OR regulation* OR professional ) OR ABS ( role* OR capabilit* OR environment* OR resource* OR survey* OR routine* OR guideline* OR limit* OR identity OR optimis* OR consequence* OR reinforcement OR intention* OR goal* OR memory OR attention OR context* OR resource* OR emotion* ) ) ) AND ( LIMIT-TO ( LANGUAGE , "English" ) )

**Web of Science All Databases**

TITLE: ("blood cultur*" or "diagnostic stewardship*" or ( ( diagno* OR detect* ) AND ( "bloodstream infec*" OR bacteraemia* OR bacteremia* ) )) AND TOPIC: (knowledge or attitude* or practice* or perception* or KAP or belief* or motivation* or barrier* or enable* or facilitate* or obstacle* or behavio* or social* or order* or skill* or competen* or abilit* or memory* or decision* or regulation* or professional* or role* or capabilit* or environment* or resource* or survey* or routine* or guideline* or limit* or identity or optimis* or consequence* or reinforcement or intention* or goal* or memory or attention or context* or resource* or emotion*)

Refined by: LANGUAGES: ( ENGLISH )

Timespan: All years. Databases: WOS, BCI, CCC, DRCI, DIIDW, KJD, MEDLINE, RSCI, SCIELO, ZOOREC.

**WHO Global Index Medicus**

Title, abstract, subject: ("blood cultur*" or "diagnostic stewardship*" or ((diagno* OR detect*) AND ("bloodstream infec*" OR bacteraemia* OR bacteremia*))) AND (knowledge or attitude* or practice* or perception* or KAP or belief* or motivation* or barrier* or enable* or facilitate* or obstacle* or behavio* or social* or order* or skill* or competen* or abilit* or memory* or decision* or regulation* or professional* or role* or capabilit* or environment* or resource* or survey* or routine* or guideline* or limit* or identity or optimis* or consequence* or reinforcement or intention* or goal* or memory or attention or context* or resource* or emotion*)

Filter: English

**Clinicaltrials.gov**

**Other terms: blood culture**

**CRD PROSPERO**

[**https://www.crd.york.ac.uk/prospero/#searchadvanced**](https://www.crd.york.ac.uk/prospero/#searchadvanced)

**NB WHO International Clinical Trials Registries – not operational at this time**

[**https://www.who.int/ictrp/en/**](https://www.who.int/ictrp/en/)

**Appendix S3. Theoretical Domains Framework: Definitions and examples**

| TDF domain and definition | Examples related to blood culture sampling |
| --- | --- |
| Knowledge: awareness of the existence of something | In the context of this study, knowledge of the condition/scientific rationale could relate to their knowledge of:   - when and whom blood culture (BC) should be sampled - local and international guidelines for BC sampling     Knowledge may be both correct and incorrect |
| Skills: ability or proficiency acquired through practice | In the context of this study, skills/competence include skill of participant to draw blood for BC sample collection.  Skills may be both present and absent |
| Social professional role and identity: a coherent set of behaviours and displayed personal qualities of an individual in a social or work setting | In the context of this study, professional role may relate to the extent that healthcare professionals feel that ordering or initiating an order for BC are part of their professional role or their job description.  Personal identity may relate to how a participant views their role of   - ordering or initiating an order for BC - drawing blood for BC |
| Beliefs about capabilities: acceptance of the truth/reality about or validity of an ability, talent or facility that a person can put to constructive use | In the context of this study, beliefs about capabilities relates to the judgments on medical doctor/final-year medical student’s ability to:   - draw blood successfully - draw blood appropriately   As BC may be ordered by respondents but collected by other professionals, beliefs about capabilities also include their judgments on the ability of persons who are tasked to draw blood  “Successfully” means obtaining blood, and “Appropriately” means that general guidelines for BC specimen collection such as aseptic technique are followed. |
| Optimism: confidence that things will happen for the best or that desired goals will be attained | In the context of this study, optimism related to their judgment regarding that a BC will be sampled and processed in the laboratory appropriately if they order a BC.  This includes optimism and pessimism. |
| Beliefs about consequences*:* acceptance of the truth/reality about or validity of outcomes of a behaviour in a given situation | In the context of this study, beliefs about their judgments on:   - the purpose, value, and effectiveness of BC - negative/positive outcomes of BC |
| Reinforcement: increasing the probability of a response by arranging a dependent relationship, or contingency, between the response and a given stimulus | In the context of this study, reinforcements relate to their judgments on:   - receiving an incentive or reward (these can be social [e.g. praise] or material [e.g. a positive score]) for ordering a BC when recommended - receiving any negative consequences (these can be social [e.g. verbal reprimand or that you/doctors are at risk of being scrutinized] or material [e.g. a negative score]) for not ordering BC when recommended   As feedbacks could discourage the behavior, reinforcement also include judgements on:   - receiving any negative consequences for ordering BC when recommended |
| Intentions: conscious decision to perform a behaviour or a resolve to act in a certain way | In the context of this study, intentions relate to the statements on their intention to order BC. |
| Goals: mental representation of outcomes or end states that an individual wants to achieve | In the context of this study, goals relate to the statements on:   - the goals they wish to collect BC prior to giving empirical antibiotics - competing goals (goals that might conflict with BC collection; e.g. giving empirical antibiotics) |
| Memory, attention and decision processes: ability to retain information, focus selectively on aspects of the environment and choose between two or more alternatives | In the context of this study, memory, attention and decision processes relate the statements on how they decide whether to order or not order BC |
| Environmental context and resources: any circumstances of a person’s situation or environment that discourages or encourages the development of skills and abilities, independence, social competence, and adaptive behaviour | In the context of this study, environmental context and resources relates to their perceptions of the:   - Availability of consumables such as bottles, needles, syringes, blood collection set, etc. - Availability of microbiology laboratories - Financial resources, whether patients have to pay out-of-pocket - Cost-effectiveness of BC |
| Social influences: interpersonal processes that can cause an individual to change their thoughts, feeling or behaviours. | In the context of this study, social influences relate to the statements expressing the influence of others on attending BC. Including:   - norms - influences from nurses, other medical doctors, consultants, head of department, executive of the hospitals, patients and family of patients   “Norms” mean usual practice that are typical of or accepted within their hospital. |
| Emotion: a complex reaction pattern, involving experiential, behavioural and physiological elements, by which the individual attempts to deal with a personally significant matter or event | In the context of this study, emotions relate to the statements of expressing their emotional reaction/state relating to order and sample for BC  Any logical reasons or social influence which are stated as “fear of” are categorized as “Memory, attention and decision processes” or “Social influence” as appropriate. |
| Behavioural regulation: anything aimed at managing or changing objectively observed or measured actions | In the context of this study, behavioural regulation relates to the statements about managements or steps taken to   - order BC - adopt local/national/international guidelines for BC sampling |

**Appendix S4. TDF survey questionnaire**

**Online research participant information sheet and electronic consent form**

You are invited to participate in a web-based online survey on **“Barriers and facilitators to ordering blood culture samples in Indonesia, Thailand and Viet Nam”.** This is a research project being conducted under the collaboration between Eijkman Oxford Clinical Research Unit (EOCRU), **Indonesia,** and Mahidol Oxford Tropical Medicine Research Unit (MORU), Faculty of Tropical Medicine, Mahidol University, **Thailand**, Oxford University Clinical Research Unit (OUCRU), **Viet Nam,** Centre for Behaviour Change, University College London, **United Kingdom**.

**PROPOSE:** This study aim to identify barrier and facilitators to the adoption of blood culture sampling recommendations in Indonesia, Thailand and Viet Nam

**PARTICIPATION:** The participants include 1,500 medical doctors and final-year medical students in Indonesia, Thailand and Viet Nam (500 participants per country). The survey is voluntary. You may refuse to take part in the research or exit the survey at any time without penalty. You are free to decline to answer any particular question you do not wish to answer for any reason.

**PROCEDURE:** You may have received an invitation from clinical directors, head of final-year medical student, or head of recently graduated medical doctors to do this online survey. You may also receive two email reminders about the invitation. We also ask final-year medical students and medical doctors in those hospitals to share the invitation to the survey to any final-year medical students and medical doctors in the country using their networks such as Facebook, Line and WhatsApp application.

In this survey, we will ask whether you know of any local and international guidelines on when to perform blood culture sampling, whether you would perform blood culture sampling for the constructed case scenario, and why you do or do not perform blood culture sampling. It should take approximately 30 – 40 minutes to complete.

All study data will be entered on a Qualtrics. The participants will be identified by a unique study specific number and/or code in any database. We will ask for your email account or telephone number in order to provide you an electronic gift. You may refuse to providing your email account or telephone number and to receiving an electronic gift. The name and any other identifying detail will NOT be included in any study data electronic file.

**BENEFITS:** You will receive no direct benefits from participating in this research study. However, your responses may help us learn more about what are barriers and facilitators of doctors to order and collect blood culture samples per local, national or international recommendations in different countries. The questionnaire focuses only on when and why blood culture is sampled. Participants will receive a gift or cash (about $4 USD in value) for completing the questionnaire. Participants could receive the gift electronically if email account or telephone number is provided.

**RISKS:** There is the risk that you may find some of the questions to be sensitive, and that some questions may cause emotional discomfort. Nonetheless, the possible risks or discomforts of the study are minimal. If you feel uncomfortable or distressed at any time during this survey, you should feel free to terminate participation. You are free to decline to answer any particular question you do not wish to answer for any reason. The study team does not expect any risks for participants beyond the minimal risks described above regarding confidentiality surrounding sensitive comments that might arise when participating in the qualitative interviews.

**WITHDRAWAL:** The survey is voluntary. You can withdraw from the study without penalty at any time and you are free to decline to answer any particular question you do not wish to answer for any reason with no obligation to give the reason for withdrawal.

**CONFIDENTIALITY:** Although we will collect your identifying information such as your medical license number (student identification no if you are a medical student), email address and telephone number, your identifying information are needed for compensation and your identifying information will be known only to the researchers performing this study or to specific groups for auditing purposes (if requested). These groups are government institutions or organisations authorised to conduct audits such as the ethics committee. Only summary results will be published and anonymous information will be put in open-access scientific database. No one will be able to identify you or your answers, and no one will know whether you participated in the study.

**ETHICAL:** The study protocol, informed consent form, participant information sheet and any proposed advertising material will be submitted to OxTREC, the ethics Committee of the Faculty of Tropical Medicine, Mahidol University, Thailand and (FTMEC), and local ethics committees for written approval.

**CONTACT:** If you have questions at any time about the study or the procedures, you may contact Dr Ralalicia Limato in Indonesia, Pornpan Suntornsut in Thailand, and Dr Vu Thi Lan Huong in Viet Nam.

**DATA PROTECTION**: The University of Oxford is responsible for ensuring the safe and proper use of any personal information you provide, solely for research purposes.

**DATA SHARING**: Data collected for this study will be de-identified and may be shared with other groups of researchers in accordance with the current MORU Data Sharing Policy. All applications will be carefully reviewed by the MORU Data Access Committee before granting any approvals to access data. All researchers accessing the data need to adhere to a set of terms and conditions that aim to protect the interests of research participants and other relevant stakeholders.

**INTERNET AND DEVICE REQUIREMENT:** This online questionnaire requires good internet connection and relatively up-to-date devices. Mobile devices with small screens may not show the questions clearly. If your devices are relatively out-of-date or with small screens, we recommend you to use a desktop computer at a place with good internet connection. If you have a problem with the online questionnaire, you may ask for the word file (.doc) or the paper questionnaire by contacting Dr Ralalicia Limato in Indonesia, Pornpan Suntornsut in Thailand, and Dr Vu Thi Lan Huong in Viet Nam.

**ELECTRONIC CONSENT:** Please select your choice below. You may print a copy of this consent form for your records. Clicking on the “Agree” button indicates that I agree to participate in the research study. I have read the above information and I am participating voluntarily.

○ Agree

○ Disagree

**EXPLANATION: The questionnaire may contain ○ for radio button (can take only one answer) ☐ for multiple choices (can take more than one answer)) and open text answer as well. Please indicate your level of opinion and mark in the button or box of your answer.**

Q1-1. **At which type of hospital are you currently working?** If you are currently working at more than one hospital, select where you are currently spending most time. (please select the most relevant answer)

○ Government hospital	(including National hospital, Provincial hospital, District hospital)

○ Private hospital

○ University hospital

○ I do not want to answer

○ Other: ……………

Q1-2. **What is your Medical license number or student ID number?** This is to confirm that you are a medical doctor or a final-year medical student in Indonesia, Thailand or Viet Nam. If you are not a medical doctor or a final-year medical student in Indonesia, Thailand or Viet Nam, you should not participate in this questionnaire. Your identifying information will be known only to the researchers. No one will be able to identify you or your answers, and no one will know whether you participated in the study.

………………………………………………………………….

Q1-3. **As an introduction to the topic blood culture sampling, we present a case scenario to you. We would like to know if you consider taking blood culture samples in your everyday clinical practice and your current hospital setting.**

**If you are currently working at more than one hospital, please consider the hospital you are spending most time as your current hospital setting.**

**case scenario.** “A 72-year-old woman who was brought to the emergency department of your hospital by her daughter when she noticed the patient was more confused than her baseline and was found to have a high fever and fast breathing. She had an auscultatory finding compatible with pneumonia. It is decided that this patient will be admitted to your hospital.”

If you have an authority to take a blood culture, would you take blood culture sample(s) in this case on admission?

○ Definitely (>95-100% of the time)

○ Likely (75-95% of the time)

○ Maybe (25-74% of the time)

○ Unlikely (5-24% of the time)

○ Rarely (ranging from never <5% of the time)

○ I do not know

○ I do not want to answer

Q1-4. Do you know of any recommendation(s) or guideline(s) for blood culture sampling being used in your hospital?

○ Yes

○ No, my hospital does not use any recommendations or guidelines for blood culture sampling (go to Q1-8)

○ I do not know if my hospital uses any recommendations or guidelines. (go to Q1-8)

○ I do not want to answer (go to Q1-8)

(Page break)

Q1-5. **Based on your understanding**, do any following statement(s) represent the recommendation(s) or guideline(s) for blood culture sampling being used in your hospital? (you can select more than one answer)

☐ Recommend blood culture sampling in all patients presenting with SIRS (Systemic inflammatory Response Syndrome [SIRS] is defined as having at least two of the following criteria: fever or hypothermia, tachycardia, tachypnea, and leukocytosis or leucopenia)

☐ Recommend blood culture sampling in all patients presenting with sepsis (‘sepsis’ here is defined as an acute change in total Sequential Organ Failure Assessment [SOFA] score ≥2 points consequent to the infection based on the most recent definition of sepsis [Sepsis-3 criteria])

☐ Recommend blood culture sampling in all patients presenting with septic shock

☐ Recommend blood culture sampling in all patients starting parenteral antibiotic treatment

☐ Recommend blood culture sampling in all patients with no clinical improvement after receiving empirical antibiotics

☐ Recommend blood culture sampling in all patients presenting with infection and having underlying diseases

☐ Recommend blood culture sampling in all patients with chronic fever

☐ Recommend blood culture sampling in all patients with fever of unknown origins

☐ Recommend blood culture sampling in all patients suspected of infections caused by atypical organisms

☐ Recommend blood culture sampling in all patients suspected of infections caused by antimicrobial-resistant organisms

☐ Recommend blood culture sampling in all patients suspected of infections caused by multiple-drug-resistant organisms

☐ Recommend blood culture sampling in all patients suspected of hospital-acquired infections

☐ I do not know

☐ I do not want to answer

☐ Other: ……………

**Due to many factors, there are times that doctors can not follow the recommendation(s) or guideline(s).**

Q1-6. In your current hospital setting, how often do you plan to follow the recommendation(s) or guideline(s) for blood culture sampling being used in your hospital?

○ All the time (>95-100% of the cases)

○ Often (75-95% of the cases)

○ Moderately (25-74% of the cases)

○ Occasionally (5-24% of the cases)

○ Rarely (ranging from never to <5% of the cases)

○ I do not know

○ I do not want to answer

Q1-7. Apart from the recommendation(s) or guideline(s) being used at your hospital (as you answered in the previous question), do you have **any additional reasons** for deciding to do blood culture sampling? (you can select more than one answers that are applicable to your current hospital setting)

☐ No. All reasons are stated in the recommendation(s) or guideline(s) being used in my hospital.

☐ Patients presenting with chills

☐ Patients presenting with sepsis

☐ Patients presenting with septic shock

☐ Patients starting parenteral antibiotic treatment

☐ Patients with no clinical improvement after receiving empirical antibiotics

☐ Patients presenting with infection and having underlying diseases

☐ Patients presenting with chronic fever

☐ Patients presenting with fever of unknown origin

☐ Patients suspected of infections caused by atypical organisms

☐ Patients suspected of infections caused by antimicrobial-resistant organisms

☐ Patients suspected of infections caused by multiple-drug-resistant organisms

☐ Patients suspected of hospital-acquired infections

☐ Laboratory results showing leukocytosis

☐ Laboratory results showing neutropenia

☐ Laboratory results showing left shift in blood count (i.e. showing immature white blood cells)

☐ Laboratory results showing CRP increase

☐ Laboratory results showing procalcitonin increase

☐ Patients can afford the cost of blood culture

☐ Patients have a health scheme or insurance that covers the cost of blood culture

☐ Patients are likely to have a final diagnosis that includes the cost of blood culture in the package of fee for service

☐ I do not know

☐ I do not want to answer

☐ Other: ……………

(Skip to Q1-9 after this question)

(Page break)

Q1-8. In your current hospital setting, what are the **reasons** for deciding to do blood culture sampling? (you can select more than one answer that are applicable for your current hospital setting)

☐ Patients presenting with chills

☐ Patients presenting with sepsis

☐ Patients presenting with septic shock

☐ Patients presenting with infection and having underlying diseases

☐ Patients starting parenteral antibiotic treatment

☐ Patients with no clinical improvement after receiving empirical antibiotics

☐ Patients presenting with infection and having underlying diseases

☐ Patients presenting with chronic fever

☐ Patients presenting with fever of unknown origin

☐ Patients suspected of infections caused by atypical organisms

☐ Patients suspected of infections caused by antimicrobial-resistant organisms

☐ Patients suspected of infections caused by multiple-drug-resistant organisms

☐ Patients suspected of hospital-acquired infections

☐ Laboratory results showing leukocytosis

☐ Laboratory results showing neutropenia

☐ Laboratory results showing left shift in blood count

☐ Laboratory results showing CRP increase

☐ Laboratory results showing procalcitonin increase

☐ Patients can afford the cost of blood culture

☐ Patients have a health scheme or insurance that covers the cost of blood culture

☐ Patients are likely to have a final diagnosis that includes the cost of blood culture in the package of fee for service

☐ I do not know

☐ I do not want to answer

☐ Other: ……………

(Page break)

Q1-9. Are you aware of any international recommendation(s) or guideline(s) for blood culture sampling? Examples of international recommendations are surviving sepsis campaign (SSC), the diagnostic stewardship of the World Health Organization (WHO), The Infectious Diseases Society of America (IDSA) and The National Institute for Health and Care Excellence (NICE)

○ Yes

○ No (go to Q2-1)

○ I do not want to answer (go to Q2-1)

Q1-10. **Based on your understanding**, can any following statement(s) represent international recommendation(s) for blood culture sampling (you can select more than one answers)

☐ Recommend collecting blood culture in all patients presenting with sepsis

☐ Recommend collecting blood culture in all patients starting parenteral antibiotic treatment

☐ I do not know

☐ I do not want to answer

☐ Other:…………………….

(Page break)

**We would like to understand your current job and how** **doctors in different positions are involved in ordering and collecting blood culture in your current hospital setting.**

Q2-1. First, please state your current job. (please select the most relevant answer)

○ Medical doctor – working in an executive or administrative position (not doing clinical work)

○ Medical doctor – working as a consultant (defined as a doctor with a clinical specialty/subspecialty degree)

○ Medical doctor – working as a physician (defined as a doctor without a clinical specialty/subspecialty degree and not under any postgraduate clinical training)

○ Medical doctor – working as a resident/registra/fellow (defined as a doctor who is currently under any postgraduate clinical training)

○ Intern (defined as a recent medical school graduate who is in the first year of post-graduate on-the-job training)

○ Final-year medical student

○ Other:……………

Final-year medical students (and interns) in some countries or some settings can **initiate an** **order** for a blood culture under authority of residents, consultants or other medical doctors. The order may be supervised, signed or co-signed by residents, consultants or other medical doctors later.

Q2-2. In your current hospital setting, which types of professionals/staff **can order** a blood culture. **“Order**” means initiating an order either verbally or in writing. (you can select more than one answers)

☐ Medical doctors – working in executive or administrative positions (not doing clinical work)

☐ Medical doctors – working as consultants (defined as a doctor with a clinical specialty/subspecialty degree)

☐ Medical doctors – working as physicians (defined as a doctor without a clinical specialty/subspecialty degree and not under any postgraduate clinical training)

☐ Medical doctors – working as residents/registras/fellows (defined as a doctor who is currently under any postgraduate clinical training)

☐ Interns (defined as recent medical school graduates who are in the first year of post-graduate on-the-job training)

☐ Final-year medical students

☐ I do not want to answer

☐ Other:……………

Q2-3. Do you know when and which patients should receive an **order** for a blood culture in your hospital?

○ Definitely (>95-100% of the case)

○ Likely (75-95% of the case)

○ Uncertain (25-74% of the case)

○ Unlikely (5-24% of the case)

○ Rarely (ranging from never to <5% of the case)

○ I do not know

○ I do not want to answer

Q2-4. If you **can** **order** for a blood culture as per your current job description or position, do you think that it is an appropriate part of your current job (as per your job description or position) to **order** a blood culture?

○ Very appropriate

○ Appropriate

○ Uncertain

○ Inappropriate

○ Very inappropriate

○ I cannot order blood culture. It is not part of my job (Go to Q2-5).

○ I do not know

○ I do not want to answer

(Skip to Q2-6 after this question, except answering “I cannot order blood culture. It is not part of my job”)

(Page break)

Q2-5. As you **cannot** **order** for a blood culture as per your current job description or position, do you think that it would be an appropriate part of your current job (as per your job description or position) to **order** a blood culture?

○ Very appropriate

○ Appropriate

○ Uncertain

○ Inappropriate

○ Very inappropriate

○ I do not know

○ I do not want to answer

(Page break)

Q2-6. In your current hospital setting, which **types of professionals** are tasked to **draw blood** from patients for blood culture. (you can select more than one answers)

☐ Medical doctors – working in executive or administrative positions (not doing clinical work)

☐ Medical doctors – working as consultants (defined as a doctor with a clinical specialty/subspecialty degree)

☐ Medical doctors – working as physicians (defined as a doctor without a clinical specialty/subspecialty degree and not under any postgraduate clinical training)

☐ Medical doctors – working as residents/registras/fellows (defined as a doctor who is currently under any postgraduate clinical training)

☐ Interns (defined as recent medical school graduates who are in the first year of post-graduate on-the-job training)☐ Interns

☐ Final-year medical students

☐ Registered nurses

☐ Microbiology laboratory team

☐ Specialized blood draw team

☐ I do not want to answer

☐ Other:……………

Q2-7. Do you think that it is an appropriate part of your job (as per your job description or position) to **draw blood**?

○ Very appropriate

○ Appropriate

○ Uncertain

○ Inappropriate

○ Very inappropriate

○ It is not part of my job to draw blood from patients for blood culture (go to Q2-11)

○ I do not know

○ I do not want to answer

(Page break)

Q2-8. How skilled are you in **drawing blood**?

○ Very good skill

○ Good skill

○ Fair skill

○ Poor skill

○ Very poor skill

○ I do not know

○ I do not want to answer

**Having confidence is different from having skills. Due to many factors, there are times that blood could not be drawn even though we are skilled.**

Q2-9. If you have to draw blood yourself, are you confident that **you can** **draw blood** **successfully**? “Successfully” means obtaining blood.

○ Strongly confident

○ Confident

○ Uncertain

○ Doubtful

○ Strongly doubtful

○ It is not part of my job to draw blood from patients for blood culture

○ I do not know

○ I do not want to answer

Q2-10. Are you confident that **you can** **draw blood** **appropriately**? “Appropriately” means that general recommendations for blood culture specimen collection such as aseptic technique are followed.

○ Strongly confident

○ Confident

○ Uncertain

○ Doubtful

○ Strongly doubtful

○ It is not part of my job to draw blood from patients for blood culture

○ I do not know

○ I do not want to answer

(Page break)

Q2-11. Are you confident that **others (who are tasked to draw blood in your hospital) can** **draw blood** **successfully**?

○ Strongly confident

○ Confident

○ Uncertain

○ Doubtful

○ Strongly doubtful

○ I do not know

○ I do not want to answer

○ I do not want to answer

Q2-12. Are you confident that **others (who are tasked to draw blood in your hospital) can** **draw blood** **appropriately**? “Appropriately” means that general recommendations for blood culture specimen collection such as aseptic technique are followed.

○ Strongly confident

○ Confident

○ Uncertain

○ Doubtful

○ Strongly doubtful

○ I do not know

○ I do not want to answer

Q2-13. In your current hospital setting, how **optimistic** are you that a blood culture will be sampled and processed in the laboratory appropriately if you order a blood culture? “Optimistic” means the confidence that things will happen for the best or that desired goals will be attained.

○ Strongly optimistic

○ Optimistic

○ Neither optimistic nor pessimistic

○ Pessimistic

○ Strongly pessimistic

○ I do not know

○ I do not want to answer

(Page break)

**Many advantages and disadvantages of blood culture have been mentioned in surveys in different countries. This advantages and disadvantages could differ between settings.**

**Please answer of all following question to the best of your ability. Please a check mark “√ “ in the appropriate answer for each question.**

| Q3-1. Do you agree or disagree about the following  potential advantages of blood culture in your current hospital setting? | Strongly agree | Agree | Uncertain | Disagree | Strongly disagree | I do not know | I do not want to answer |
| --- | --- | --- | --- | --- | --- | --- | --- |
| • Blood culture is helpful in clinical decisions. |  |  |  |  |  |  |  |
| • Blood culture is helpful to rule in an infection. |  |  |  |  |  |  |  |
| • Blood culture is helpful to rule out an infection. |  |  |  |  |  |  |  |
| • Blood culture is helpful in detecting antimicrobial-resistant bacterial infections. |  |  |  |  |  |  |  |
| • Blood culture is helpful in adjusting antibiotics. |  |  |  |  |  |  |  |
| • Blood culture can reduce overuse of antibiotics. |  |  |  |  |  |  |  |
| • Blood culture can reduce length of hospital stay. |  |  |  |  |  |  |  |
| • Blood culture can reduce patient mortality. |  |  |  |  |  |  |  |
| • Accumulative results of blood culture (i.e. antimicrobial-resistance surveillance report) are helpful in understanding epidemiology of antimicrobial-resistant bacterial infections. |  |  |  |  |  |  |  |

Q3-2. Additional comments why blood culture is helpful in your current hospital setting (Note: limit to 2,000 characters)

…………………………………………………………………

**Please answer of all following question to the best of your ability. Please a check mark “√ “ in the appropriate answer for each question.**

| Q3-3. Do you agree or disagree about the following disadvantages of blood culture, **making blood culture unnecessary in your current hospital setting?** | Strongly agree | Agree | Uncertain | Disagree | Strongly disagree | I do not know | I do not want to answer |
| --- | --- | --- | --- | --- | --- | --- | --- |
| • Blood culture is unnecessary because antibiotic therapy can be determined based on clinical presentations. |  |  |  |  |  |  |  |
| • The therapeutic consequence of blood culture sampling is questionable. |  |  |  |  |  |  |  |
| • The scientific basis of the guideline on blood culture is questionable |  |  |  |  |  |  |  |
| • Blood culture is unnecessary because results are often delayed. |  |  |  |  |  |  |  |
| • Blood culture is unnecessary because results are often not interpretable. |  |  |  |  |  |  |  |
| • Blood culture is unnecessary because results are often negative or no growth. |  |  |  |  |  |  |  |
| • Blood culture is unnecessary because cultures are often contaminated. |  |  |  |  |  |  |  |
| • Blood culture is unnecessary because results often do not agree with clinical signs. |  |  |  |  |  |  |  |
| • Blood culture is unnecessary because a contaminated result often leads to wrong therapeutic approaches. |  |  |  |  |  |  |  |
| • Blood culture is unnecessary because it is too expensive. |  |  |  |  |  |  |  |
| • Blood culture is not benefiting the patients. |  |  |  |  |  |  |  |
| • It is not too late to collect blood culture later, particularly if patients do not improve after receiving empirical antibiotic treatment. |  |  |  |  |  |  |  |
| • Quality of laboratory is questionable. |  |  |  |  |  |  |  |
| • Levels of local antibiotic resistance are low. |  |  |  |  |  |  |  |

Q3-4. Additional comments why blood culture is not helpful in your current hospital setting (Note: limit to 2,000 characters)

…………………………………………………………………

(Page break)

**In different settings, other tasks may be considered more urgent than collecting blood culture samples.**

Q3-5. In your current hospital setting, how often do you obtain blood culture **prior to administration of empirical antibiotics** in patients presenting with **sepsis**? ‘(‘sepsis’ here is defined as an acute change in total Sequential Organ Failure Assessment [SOFA] score ≥2 points consequent to the infection based on the most recent definition of sepsis [Sepsis-3 criteria])

○ All the time (>95-100% of the time)

○ Often (75-95% of the time)

○ Moderately (25-74% of the time)

○ Occasionally (5-24% of the time)

○ Rarely (ranging from never to <5% of the time)

○ I do not know

○ I do not want to answer

Q3-6. In your current hospital setting, how often do you obtain blood culture **prior to administration of empirical antibiotics** in patients presenting with **septic shock**?

○ All the time (>95-100% of the time)

○ Often (75-95% of the time)

○ Moderately (25-74% of the time)

○ Occasionally (5-24% of the time)

○ Rarely (ranging from never to <5% of the time) ○ Rarely (ranging from never to <5% of the time)

○ I do not know

○ I do not want to answer

**Even if blood culture is recommended, doctors may decide not to order blood culture in some situations.**

**Please answer of all following question to the best of your ability. Please a check mark “√ “ in the appropriate answer for each question.**

| Q3-7. Would you still order blood culture in the following situation? | Definitely not order | Likely not order | Maybe not order | Likely to still order | Very likely to still order | I do not Know | I do not want to answer |
| --- | --- | --- | --- | --- | --- | --- | --- |
| • Patients are already on antibiotics. |  |  |  |  |  |  |  |
| • Patients have anemia. |  |  |  |  |  |  |  |
| • Blood should be used for other laboratory tests. |  |  |  |  |  |  |  |
| • There are no local guidelines/recommendations for blood culture sampling |  |  |  |  |  |  |  |
| • Patients do not meet certain conditions for a blood culture following the local guidelines |  |  |  |  |  |  |  |
| • Patients do not have a health scheme or insurance that covers the cost of blood culture |  |  |  |  |  |  |  |
| • Microbiology laboratory in your hospital is not available |  |  |  |  |  |  |  |

Q3-8. Additional comments why you do not order blood culture regarding situations mentioned above (Note: limit to 2,000 characters)

…………………………………………………………………

(Page break)

**Resources are commonly limited in many settings worldwide.**

Q4-1. In your hospital, how often could you (or doctors in your hospital) **not order blood culture** because consumables (such as blood culture bottles, needles, syringes, blood collection set, etc.) are **not available**?

○ All the time (>95-100% of the time)

○ Often (75-95% of the time)

○ Moderately (25-74% of the time)

○ Occasionally (5-24% of the time)

○ Rarely (ranging from never to <5% of the time)

○ I do not know

○ I do not want to answer

Q4-2. In your hospital, how often could you (or doctors in your hospital) **not order blood culture** because the microbiology laboratory is **not available** or not functioning?

○ All the time (>95-100% of the time)

○ Often (75-95% of the time)

○ Moderately (25-74% of the time)

○ Occasionally (5-24% of the time)

○ Rarely (ranging from never to <5% of the time)

○ I do not know

○ I do not want to answer

Q4-3. In your hospital, how often do patients have to pay for blood culture using their own money (i.e. out of pocket)?

○ All the time (>95-100% of the patients)

○ Often (75-95% of the patients)

○ Moderately (25-74% of the patients)

○ Occasionally (5-24% of the patients)

○ Rarely (ranging from never to <5% of the patients)

○ I do not know I do not know

○ I do not want to answer

Q4-4. Regardless of who pays for the cost of blood culture, would you say that the benefits of blood culture outweigh the cost?

○ Very likely

○ Likely

○ Uncertain

○ Unlikely

○ Very unlikely

○ I do not know

○ I do not want to answer

(Page break)

**Positive and negative consequences could encourage us to follow guidelines.**

Q5-1. Are there **any positive consequences, incentives or rewards** (these can be social [e.g. praise] or material [e.g. a positive score]) if you or doctors in your hospital **order a blood culture when recommended**? (you can select more than one answer)

☐ No

☐ Yes- social

☐ Yes- material

☐ Yes- both social and material

☐ I do not know

☐ I do not want to answer

☐ Other: ……………

Q5-2. Are there **any negative consequences** to you or doctors (these can be social [e.g. verbal reprimand or that you/doctors are at risk of being scrutinized] or material [e.g. a negative score]) if you or doctors in your hospital **do not order a blood culture when recommended**? (you can select more than one answer)

☐ No

☐ Yes- social

☐ Yes- material

☐ Yes- both social and material

☐ I do not know

☐ I do not want to answer

☐ Other: ……………

**Sometimes there are feedbacks that could discourage us to follow guidelines. This could be due to many reasons based on local context.**

Q5-3. Are there **any negative consequences** to you or doctors (these can be social [e.g. verbal reprimand or any pressure from your supervisors/executives of your hospital as the hospital (may) have to pay for the (extra) cost of blood culture] or material [e.g. a negative score, that you/doctors are at risk of having to spend extra time and effort to reimburse the cost of blood culture from any health scheme or insurance, or that you/doctors are at risk of having to pay for the (extra) cost of blood culture yourselves]), if you or doctors in your hospital **order blood culture when recommended**? (you can select more than one answer)

☐ No

☐ Yes- social

☐ Yes- material

☐ Yes- both social and material

☐ I do not know

☐ I do not want to answer

☐ Other: ……………

Q5-4. Additional comments about feedbacks (including encouragement, punishments or any positive and negative consequences) on blood culture sampling in your hospital setting. Also, please provide more comments about whether any consequences you would recommend to implement in your hospital to support blood culture ordering.

…………………………………………………………………

(Page break)

Q5-5. In your hospital, are there **any training, lectures, classes or meetings** that provide you knowledge about local/national/international guidelines for blood culture sampling? (you can select more than one answers)

☐ No

☐ Yes, infrequently (less than once a year)

☐ Yes, occasionally (at least once a year)

☐ Yes, regularly (more than once a year)

☐ I do not know

☐ I do not want to answer

☐ Other: ……………

Q5-6. In your hospital, are there **any procedures** that support you or doctors in your hospital to order or regulate ordering of blood culture per local/national/international guidelines? (you can select more than one answers)

☐ No

☐ Yes, there is a poster (and blood culture is mentioned)

☐ Yes, there is a standard order form for patients presenting with sepsis (and blood culture is already written in the order form)

☐ Yes, there is a computer system to remind ordering blood culture

☐ Yes, there is a case review (e.g. grand round; morning ward round, clinical meetings, etc and blood culture is often mentioned)

☐ Yes, there is a stewardship programme and reviewing blood culture is included in the programme (e.g. post-prescription review and stewardship round, etc.)

☐ Yes, there is a local hospital guideline (e.g. standard operating procedure [SOP])

☐ I do not know

☐ I do not want to answer

☐ Other: ……………

(Page break)

**Due to different personal beliefs, norms and limitations, blood culture sampling is encouraged or discouraged by peers and co-workers in different settings.**

Q6-1. **To what extent do you or doctors in your hospital order blood culture sampling because you are following local norms? “**Norms” mean usual practice that are typical of or accepted within your hospital.

○ All the time (>95-100% of the time)

○ Often (75-95% of the time)

○ Moderately (25-74% of the time)

○ Occasionally (5-24% of the time)

○ Rarely (ranging from never to <5% of the time)

○ I do not know

○ I do not want to answer

**Please answer of all following question to the best of your ability. Please a check mark “√ “ in the appropriate answer for each question.**

| Q6-2. **Do following people have any positive or negative influence on you or doctors in your hospital to order blood culture?** Positive influence could mean facilitate, support or encourage blood culture sampling. Negative influence could mean hinder or discourage blood culture sampling. | Very positive influence | Positive influence | Neither positive nor negative influence | Negative influence | Very negative influence | I do not know | I do not want to answer |
| --- | --- | --- | --- | --- | --- | --- | --- |
| • Nurses |  |  |  |  |  |  |  |
| • Final-year medical students |  |  |  |  |  |  |  |
| • Interns |  |  |  |  |  |  |  |
| • Residents (any postgraduate clinical training) |  |  |  |  |  |  |  |
| • Doctors (defined as a doctor without a specialty/subspecialty degree and not under any postgraduate clinical training) |  |  |  |  |  |  |  |
| • Consultants (defined as a doctor with a clinical specialty/subspecialty degree) |  |  |  |  |  |  |  |
| • Head of the Department |  |  |  |  |  |  |  |
| • Executives of the hospital |  |  |  |  |  |  |  |
| • Patients |  |  |  |  |  |  |  |
| • Family of patients |  |  |  |  |  |  |  |

Q6-3. Additional comments about social influence on blood culture sampling

…………………………………………………………………

Q6-4. Apart from your logical considerations, do you think that **any emotional factors** of anyone involved in ordering and sampling for blood culture (including patients and family of patients) could influence whether blood culture is ordered or sampled? (for example: fear of blood, fear of needle, fear of blood transmitted diseases, etc)

○ No

○ Other: ……………

Q6-5. Additional comments about emotional factors (from anyone involved in ordering and sampling for blood culture; including patients and family of patients) on blood culture sampling

…………………………………………………………………

(Page break)

**Finally, we have some questions about yourself**

Q7-1. Which country do you currently work in?

○ Thailand

○ Vietnam

○ Indonesia

○ I do not want to answer

Province of your current hospital:……………………………………… (Dropdown list for each country)

Q7-2. Are you female or male?

○ Female

○ Male

○ Other

○ I do not want to answer

Q7-3. What is the number of beds in your hospital? (Please use the official number, and please estimate if you are uncertain.)

○ < 200

○ 201 - 400

○ 401 - 600

○ 601 - 1,000

○ 1,001 - 2,000

○ > 2,000

○ I do not know

○ I do not want to answer

☐☐☐☐☐

Q7-4. In which department are you **currently working**? If your role (such as medical students) moves from one department to another department over time, please state the current department you are working in.

(you can select more than one answers; for example both internal medicine and infectious disease devision)

☐ Internal Medicine

☐ Pediatrics

☐ Infection disease division/department

☐ Surgery

☐ Orthopaedics

☐ Obstetrics / Gynaecology

☐ Emergency department

☐ Intensive care unit

☐ I do not want to answer

☐ Other: ……………

(Page break)

Q7-5. Do you want to be contacted for further studies?

○ Yes

○ No

Q7-6. Do you want to be informed the results of this study?

○ Yes

○ No

Q7-7. Your email address (If you want to be contacted via email address. Please leave it blank, if you do not want to be contact via email address)

………………………………………………………………….

Q7-8. Your phone number (if you want to be contacted via phone. Please leave it blank, if you do not want to be contact via phone)

…………………………………………………………………..

Please note that a gift or cash (about $4 in value) for completing the survey is to be provided to you. Participants could receive the gift electronically if email account or telephone number is provided.

Please make sure that you click “submit” on the next page to complete the questionnaire. Otherwise, all answers that you made and your information for compensation will not be submitted to us via the system.

(Page break)

We are grateful for your participation. Thank you very much.

**Appendix S5. PRISMA checklist**

| **Section and Topic** | **Item #** | **Checklist item** | **Location where item is reported** |
| --- | --- | --- | --- |
| **TITLE** | | |  |
| Title | 1 | Identify the report as a systematic review. | Page 1 |
| **ABSTRACT** | | |  |
| Abstract | 2 | See the PRISMA 2020 for Abstracts checklist. | Page 3 |
| **INTRODUCTION** | | |  |
| Rationale | 3 | Describe the rationale for the review in the context of existing knowledge. | Page 5, 7-8 |
| Objectives | 4 | Provide an explicit statement of the objective(s) or question(s) the review addresses. | Page 5, 7-8 |
| **METHODS** | | |  |
| Eligibility criteria | 5 | Specify the inclusion and exclusion criteria for the review and how studies were grouped for the syntheses. | Page 5, 8-9 |
| Information sources | 6 | Specify all databases, registers, websites, organisations, reference lists and other sources searched or consulted to identify studies. Specify the date when each source was last searched or consulted. | Page 5, 8-9 |
| Search strategy | 7 | Present the full search strategies for all databases, registers and websites, including any filters and limits used. | Page 5, 8-9  Appendix S2 |
| Selection process | 8 | Specify the methods used to decide whether a study met the inclusion criteria of the review, including how many reviewers screened each record and each report retrieved, whether they worked independently, and if applicable, details of automation tools used in the process. | Figure 1 |
| Data collection process | 9 | Specify the methods used to collect data from reports, including how many reviewers collected data from each report, whether they worked independently, any processes for obtaining or confirming data from study investigators, and if applicable, details of automation tools used in the process. | Page 8-9  Appendix S1 |
| Data items | 10a | List and define all outcomes for which data were sought. Specify whether all results that were compatible with each outcome domain in each study were sought (e.g. for all measures, time points, analyses), and if not, the methods used to decide which results to collect. | Page 8-9  Appendix S1 |
|  | 10b | List and define all other variables for which data were sought (e.g. participant and intervention characteristics, funding sources). Describe any assumptions made about any missing or unclear information. | Page 8-9  Appendix S1 |
| Study risk of bias assessment | 11 | Specify the methods used to assess risk of bias in the included studies, including details of the tool(s) used, how many reviewers assessed each study and whether they worked independently, and if applicable, details of automation tools used in the process. | Page 8-9  Appendix S1 |
| Effect measures | 12 | Specify for each outcome the effect measure(s) (e.g. risk ratio, mean difference) used in the synthesis or presentation of results. | Page 8-9  Appendix S1 |
| Synthesis methods | 13a | Describe the processes used to decide which studies were eligible for each synthesis (e.g. tabulating the study intervention characteristics and comparing against the planned groups for each synthesis (item #5)). | Page 8-9  Appendix S1 |
|  | 13b | Describe any methods required to prepare the data for presentation or synthesis, such as handling of missing summary statistics, or data conversions. | Page 8-9  Appendix S1 |
|  | 13c | Describe any methods used to tabulate or visually display results of individual studies and syntheses. | Page 8-9  Appendix S1 |
|  | 13d | Describe any methods used to synthesize results and provide a rationale for the choice(s). If meta-analysis was performed, describe the model(s), method(s) to identify the presence and extent of statistical heterogeneity, and software package(s) used. | Page 8-9  Appendix S1 |
|  | 13e | Describe any methods used to explore possible causes of heterogeneity among study results (e.g. subgroup analysis, meta-regression). | Page 8-9  Appendix S1 |
|  | 13f | Describe any sensitivity analyses conducted to assess robustness of the synthesized results. | Not applicable |
| Reporting bias assessment | 14 | Describe any methods used to assess risk of bias due to missing results in a synthesis (arising from reporting biases). | Page 8-9  Appendix S1 |
| Certainty assessment | 15 | Describe any methods used to assess certainty (or confidence) in the body of evidence for an outcome. | Not applicable |
| **RESULTS** | | |  |
| Study selection | 16a | Describe the results of the search and selection process, from the number of records identified in the search to the number of studies included in the review, ideally using a flow diagram. | Page 12-13, Figure 1, Appendix S1 |
|  | 16b | Cite studies that might appear to meet the inclusion criteria, but which were excluded, and explain why they were excluded. | Not applicable |
| Study characteristics | 17 | Cite each included study and present its characteristics. | Appendix S6 and S7 |
| Risk of bias in studies | 18 | Present assessments of risk of bias for each included study. | Appendix S8 |
| Results of individual studies | 19 | For all outcomes, present, for each study: (a) summary statistics for each group (where appropriate) and (b) an effect estimate and its precision (e.g. confidence/credible interval), ideally using structured tables or plots. | Appendix S9 |
| Results of syntheses | 20a | For each synthesis, briefly summarise the characteristics and risk of bias among contributing studies. | Appendix S9 |
|  | 20b | Present results of all statistical syntheses conducted. If meta-analysis was done, present for each the summary estimate and its precision (e.g. confidence/credible interval) and measures of statistical heterogeneity. If comparing groups, describe the direction of the effect. | Not applicable |
|  | 20c | Present results of all investigations of possible causes of heterogeneity among study results. | Page 12-13, Figure 2 |
|  | 20d | Present results of all sensitivity analyses conducted to assess the robustness of the synthesized results. | Not applicable |
| Reporting biases | 21 | Present assessments of risk of bias due to missing results (arising from reporting biases) for each synthesis assessed. | Appendix S6 and S8 |
| Certainty of evidence | 22 | Present assessments of certainty (or confidence) in the body of evidence for each outcome assessed. | Not applicable |
| **DISCUSSION** | | |  |
| Discussion | 23a | Provide a general interpretation of the results in the context of other evidence. | Page 22-24 |
|  | 23b | Discuss any limitations of the evidence included in the review. | Page 22-24 |
|  | 23c | Discuss any limitations of the review processes used. | Page 22-24 |
|  | 23d | Discuss implications of the results for practice, policy, and future research. | Page 22-24 |
| **OTHER INFORMATION** | | |  |
| Registration and protocol | 24a | Provide registration information for the review, including register name and registration number, or state that the review was not registered. | Page 3, 8 |
|  | 24b | Indicate where the review protocol can be accessed, or state that a protocol was not prepared. | Page 3, 8 |
|  | 24c | Describe and explain any amendments to information provided at registration or in the protocol. | Not applicable |
| Support | 25 | Describe sources of financial or non-financial support for the review, and the role of the funders or sponsors in the review. | Page 26 |
| Competing interests | 26 | Declare any competing interests of review authors. | Page 26 |
| Availability of data, code and other materials | 27 | Report which of the following are publicly available and where they can be found: template data collection forms; data extracted from included studies; data used for all analyses; analytic code; any other materials used in the review. | Page 26 |

**Appendix S6: Characteristics of studies included in the systematic review**

| **Studies** | **Countries / settings** | **Research Objectives** | **Topics or factors of investigation (relevant to our review)** | **Methodological/theoretical approach (relevant to our review)** | **Data collection**  **(relevant to our review)** | **Data analysis (relevant to our review)** | **Participants (Patient/HCP/both)** | **Sample Sizes** |
| --- | --- | --- | --- | --- | --- | --- | --- | --- |
| Gross, P. A (1988)^25^ | The United States  Medical Intensive Care Unit (MICU) | To study the BC ordering habits and evaluate implementation of a protocol for appropriate BC ordering | Behavioural regulation | Quantitative non-randomized interventional study  The intervention included an education campaign and reviews of BC order criteria | Record reviews of all BC orders for one-month period in the hospital to generate the protocol  Record reviews of all BC orders at the MICU prior to, during and after the implementation of the intervention. | Percentages  (Unknown statistical tests used) | Patients | N of the review of all BC orders in the hospital in one month = 844 BCs for 315 patients  N of patients at MICU reviewed for all BC orders = 184 patients |
| Salluzzo, R (1991)^26^ | The United States  Emergency department (ED) | To study the manner in which BC were ordered before and after the introduction of general guideline for the use of BC | Knowledge, and behavioural regulation | Quantitative non-randomized interventional study  The intervention included an instruction of new guideline and reviews of BC order criteria | Record review of all BC orders in the ED for one-year period prior to and for one-year period after the implementation of the intervention | Percentages | Patients | N of BC orders in the ED = 3,591 BCs |
| Tabriz, M. S. (2004)^21^ | The United States  Hospital | To determine the extent of and reasons for repeating BC | Memory, attention and decision processes | Quantitative | Record review of all repeated BC orders for one-month period in the hospital | Percentages,  Chi-square test | Patients | N of repeated BC = 199 BCs for 96 patients  (Unknown number of missing data) |
| Parada, J. P. (2005)^2^ | The United States  Hospital | To evaluate BC-related knowledge and its association with specialties and all levels of training | Knowledge | Quantitative | A cross sectional study using a self-administered survey and a convenience sample | ANOVA  Linear regression | HCP including physicians, fellows, residents and medical students | N of respondents =291 respondents  (Unknown number of non-respondents) |
| Falagas, M. E. (2008)^27^ | Unknown countries  Clinicians indexed as corresponding authors who had an e-mail address at the affiliation indexed by PubMed | To investigate the clinical practice of obtaining BC from patients with a central venous catheter (CVC) | Knowledge and social influences | Quantitative | A cross sectional study using a semi-structured web-based questionnaire regarding their routine clinical practice of obtaining BC from patients with a CVC in place. | Percentages | HCP including clinicians indexed as corresponding authors for articles published in international peer-reviewed journals in the fields of intensive care, medicine and haematology. | N of respondent = 386 respondents  (N of clinicians who were invited = 2,851 clinicians) |
| Kerur, B (2012)^19^ | The United States  Members of the American Academy of Pediatrics (AAP) Section on Perinatal Pediatrics | To ascertain BC practices among neonatologists in the United States | Memory, attention and decision processes, and social influences | Quantitative | A cross sectional study using an online survey. Two rounds of invitation. The second round of invitation was conducted eight weeks after the first to catch those who had not responded. | Percentages | HCP including members of the AAP Section on Perinatal Pediatrics | N of respondents = 795 respondents  (N of members of the AAP section on Perinatal Pediatrics = 2,955 members) |
| Chew, K. S. (2013)^28^ | Malaysia  Hospital | To evaluate the level of understanding among healthcare staffs in emergency department (ED), regarding BC sampling practice | Knowledge | Quantitative | A cross sectional study using a self-administered questionnaire | Percentages, (Unknown statistical tests used) | HCP including emergency medicine residents, medical officers, house officers and paramedics in ED | N of participants = 64 participants  (Unknown number of non-respondents) |
| Heine, D (2013)^29^ | The United States  Hospital | To determine the prevalence of bacteremia in pediatric patients with community-acquired pneumonia (CAP) and to test the (potential) effectiveness of newly developed guidelines for obtaining BC | Memory, attention and decision processes, and behavioural regulation | Quantitative | A retrospective chart review in pediatric patients with CAP. Then, applied guideline retrospectively, and estimated percentages of cases which met criteria | Percentages,  Chi-square test | Patients | N of patients = 330 patients  N of BC = 155 BC |
| Ojide, C. K. (2013)^15^ | Nigeria  Hospital | To study the knowledge, attitude and practice of BC sampling among doctors in a Nigerian tertiary hospital | Knowledge, beliefs about consequences, memory, attention and decision processes, and environmental context and resources | Quantitative | A cross sectional study using a self-administered semi-structured questionnaire | Percentages,  Chi-square test or Fisher’s exact test | HCP including house officers, registrars, senior registrars and consultants, from different departments including internal medicine, paediatrics, surgery, obstetrics and gynaecology  and others. | N of respondents = 88 doctors  (Unknown number of non-respondents) |
| Schmitz, R. P. (2013)^17^ | France, Germany, Italy, and UK  Hospitals | To assess the current practice in BC testing in ICUs and microbiological laboratory (LAB) practice across four European countries. | Knowledge, social professional role and identity, memory, attention and decision processes, goals, and environmental context and resources | Quantitative  * The study reported the methodology as a qualitative survey. We considered the study a quantitative study based on the MMAT.^30^ | A cross sectional study using semi-structured individual telephone interviews | Percentages | HCP including ICU directors, ICU physicians, ICU nurses, LAB directors, LAB managers and microbiologists | N of interviewees = 138 interviewees  (Unknown number of non-respondents) |
| Pavese, P. (2014)^31^ | France  Hospital | To evaluate an intervention to  improve BC practices. | Knowledge and behavioural regulation | Quantitative randomized controlled trial  The study compared two types of information dissemination: simple presentation or presentation associated with an infectious diseases (ID) specialist intervention. | A cluster randomized trial in two parallel groups. BC data were extracted each month prior to the intervention and during a follow-up period. The interventions were evaluated using medical chart reviews (practice audit). | Unit of randomization was the department  Percentages,  Chi-square test or Fisher’s exact test  Chi-squared  test for trends  Simple linear and logistic regression  Logistic regression with a random effect variable | HCP. The interventions were given to the whole hospital, including each physician, and each head nurse of each department, in the whole hospital. | N of departments being enrolled = 28 departments  N of patients being audited = 519 patients |
| Kurowski, M. E. (2015)^32^ | The United States  Hospital | To increase ordering of BC for children hospitalized with CAP from 53% to 90% in 6 months, and to evaluate the effect of obtaining BC on length of stay (LOS) | Knowledge, and behavioural regulation | Quantitative non-randomized interventional study  The intervention included three key drivers (a) education, (b) identification and mitigation/education, and (c) electronical medical record changes using plan-do-study-act cycles. | A survey prior to the design of the intervention  The interventions were evaluated using medical chart reviews. | Percentages  Wilcoxon rank-sum test  Linear regression | HCP. The interventions were given to all providers, including residents | N of patients having charts reviewed = 303 patients. |
| Ong, I. (2015) ^33^ | Singapore  Hospital | To reduce the median usage of unindicated anaerobic BC by 50% within 6 months in the pediatric wards. | Knowledge, environmental context and resources, and social influence | Quantitative non-randomized interventional study  The intervention included limiting the number of anaerobic blood culture bottles available in each ward, putting up wall-mounted reminders, bottle tags and indication forms, educational lectures, and brochures using plan-do-study-act cycles. | A survey prior to the intervention  The interventions were evaluated using medical chart reviews. | Frequencies, percentages,  t-test | HCP. The intervention was given to all staff members in the pediatric wards | N of respondents of the survey prior to the intervention = 63 doctors  (Unknown numbers of non-respondents, of patients admitted to the participating pediatric wards and of patients who fulfilled the criteria for anaerobic BC) |
| She, R.C (2015)^14^ | The United States and Germany  Hospitals | To elucidate clinicians’ perspectives on the diagnosis and management of patients with bloodstream infections (BSI) and ascertain how new diagnostic tests for BSI would influence medical decisions and potentially improve patient care. | Knowledge, beliefs about consequences, memory, attention and decision processes, and environmental context and resources | Quantitative | A cross sectional study using a self-administered questionnaire | Percentages,  Chi-square or Fisher’s exact test,  Cronbach’s alpha,  Liner regression | HCP including physicians in infectious diseases/microbiology, critical care, internal medicine, and hematology/oncology services in USA and Germany | N of respondents = 242 respondents  (Unknown number of non-respondents) |
| Casu, S. (2016)^34^ | Germany  The Federation of Medical Director Emergency Medical Services (EMS) | To obtain a general picture of the current state of the EMS with respect to rapid antibiotic treatment for sepsis | Environmental context and resources | Quantitative | A cross sectional study using a web-based survey | Percentages | HCP including medical directors of different EMS districts | N of respondents = 78 medical directors  (N of medical directors invited for the survey = 166 medical directors) |
| Sloane, A. J. (2016)^35^ | The United States  Hospital | To decrease the  collection of blood test specimens in children with uncomplicated skin and soft tissue infections. | Behavioural regulation | Quantitative non-randomized interventional study  The intervention was modified well-known propaganda posters encouraging ED staff to refrain from routine blood testing. | A non-randomized intervention study.  The intervention was evaluated using medical chart reviews prior to, during and post intervention. | Percentages,  Chi-square test,  Wilcoxon rank sum test | HCP including hospital staff, including hospitalists, emergency departments  attending physicians, and residents | N of patients being evaluated = 230 patients |
| Donner, L. M. (2017)^23^ | The United States  Hospital | To assess physicians' interpretation of rapid BC identification system results, and assess their antimicrobial prescribing patterns. | Knowledge and beliefs about capabilities | Quantitative | A cross sectional study using an electronic survey. Participants were invited via email. | Percentages,  Fisher’s exact test,  Mean,  t-test,  ANOVA,  Linear regression | HCP; including physician | N of respondents = 156 physicians  (N of physicians who were emailed the survey = 382 physicians) |
| Raupach-Rosin, H. (2017)^1^ | Germany  Hospitals | To assess knowledge, attitudes, and practice of physicians in Germany regarding BC diagnostics. | Knowledge, skills, social professional role and identity, beliefs about consequences, intensions, goals, memory, attention and decision processes, environmental context and resources, social influences, and behavioural regulation | Mixed methods | A cross-sectional mixed-methods study using qualitative focus groups and a questionnaire-based quantitative study | Thematic Analysis, Percentages,  Chi-square test,  Mean,  t-tests,  Wilcoxon rank-sum test,  Linear regression | HCP including physicians and final-year medical students | N of respondents = 706 medical professionals  (Unknown number of non-respondents) |
| Diallo, K. (2018)^22^ | 56 countries  Professionals in the European Society of Clinical Microbiology and Infectious Diseases. (ESCMID) | To explore variations in the management of patients with bloodstream infections by infection specialists, and to identify demographic and professional individual characteristics associated with Infectious Diseases Society of America (IDSA) guideline-compliant management of MRSA bacteraemia and candidaemia | Memory, attention and decision processes, and behavioural regulation | Quantitative | A cross-sectional study using a self-administered and internet-based survey. Invitations were made advertised survey using the ESCMID Newsletter as well as ESGAP (ESCMID Study Group for Antimicrobial stewardshiP) and  ESGBIS (ESCMID Study Group for Bloodstream Infections and Sepsis) networks. | Percentages  Logistic regression | HCP including hospital-based healthcare professionals (fully trained or in training) who were giving at least weekly advice to colleagues (outside their home department) on their antibiotic prescriptions for positive BC | N of respondents = 616 professionals participated from 56 countries  (Unknown number of non-respondents) |
| Garcia, R. A (2018)^36^ | The United States  Hospitals within systems represented by members of the National Corporate Infection Prevention Director Network | To understand interventions and practices in the  prevention of BC contamination and associated  adverse health care events | Knowledge, goals, environmental context and resources, and behavioural regulation | Quantitative | A cross-sectional study using a survey. Invitations were distributed via e-mail to the hospital infection prevention professionals (IPs) in their system hospitals | Percentages | HCP including hospital IPs | N of respondents = 89 respondents  (N of invitations being sent = 125) |
| WHO CAESAR (2018)^18^ | 10 countries in the WHO European Region  Hospitals | To provide guidance and inspiration to countries that are building or strengthening antimicrobial resistance surveillance and to stimulate the sharing of data internationally | Knowledge, beliefs about consequences, memory, attention and decision processes, environmental context and resources, and behavioural regulation | Qualitative – descriptive and narrative | Focus group discussions and interviews in Armenia, and narrative research in the other countries | Narrative description | HCP including clinicians, nurses,  microbiologists, epidemiologists and hospital managers | N of participating countries = 10 countries  N of participants in four focus group discussions in Armenia = unknown  N of participants to the interviews = unknown |
| Dailey, P. J. (2019)^16^ | Botswana, Cambodia, DR Congo, Ethiopia, Guinea, Lao PDR and Myanmar  Hospitals | To map out a target product profile of a simplified BC system, and to inform product development efforts. | Knowledge, beliefs about consequences, intention, environmental context and resources, and behavioural regulation | Qualitative – descriptive | Interview - structured phone interviews | Thematic Analysis | HCP including infection disease physicians, public health, clinical microbiologist, clinical researcher and technology expert | N of participants = 9 participants from 8 separated locations |
| Idelevich, E. A. (2019)^37^ | 25 European countries.  The ESCMID Study Group for Bloodstream Infections, Endocarditis and Sepsis (ESGBIES) | To assess current practices of microbiological bloodstream infection diagnostics in European microbiological laboratories. | Knowledge, environmental context and resources, social influences and behavioural regulation | Quantitative | A cross-sectional study using an online questionnaire. An invitation was performed using a snowballing technique, with each national coordinator contacting approximately ten laboratories within the country. | Percentages | HCP including microbiology laboratories | N of participating laboratories = 209 laboratories in 25 European countries (N of laboratories being invited = 238 laboratories in 28 European countries) |
| The, T. (2019)^20^ | The United States, Canada, Europe, Australia, and New Zealand  Pediatric emergency research networks | To describe current anaerobic BC practices and laboratory techniques in pediatric patients throughout an international network | Beliefs about consequences, memory, attention and decision processes, environmental context and resources, and social influences | Quantitative | A cross-sectional study using two surveys: a physician survey assessing clinical practice and a microbiology survey assessing anaerobic culture laboratory techniques. Invitation was performed using a convenience sampling | Percentages | HCP including physicians and  microbiologists | N of participating institutions = 65 institutions  (N of institutions being invited = 160 institutions) |
| Tran, P. (2020)^38^ | The United States  Hospital | To determine the impact of an electronic medical record (EMR) decision support and education/compliance feedback intervention on the collection of multiple blood cultures | Knowledge, behavioural regulation | Quantitative non-randomized interventional study  The intervention included (a) the modification of EMR BC order and (b) nursing protocols for BC collection volume | Data on the number and nurse-recorded volume of BC were collected monthly beginning in the intervention period. Data from the EMR were extracted. | Frequencies, percentages,  means,  Chi-square test, Mann-Whitney test,  t-test | HCP including nurses and providers | N of patients being evaluated = 3,948 patients  (N of nurses and providers included in the intervention = unknown) |

**Appendix S7. Study methods and locations of studies included in the systematic review**

| **Study characteristics** | **Frequencies (n=25 studies)** |
| --- | --- |
| Study methods | 15 (60%) Quantitative studies (using surveys [n=13]^2,14,15,17,19,20,22,23,27,28,34,36,37^ and chart reviews [n=2]^21,29^) |
|  | 6 (24%) Quantitative non-randomized interventional studies^25,26,32,33,35,38^ |
|  | 2 (8%) Qualitative study^16,18^ |
|  | 1 (4%) Mixed methods study^1^ |
|  | 1 (4%) Quantitative randomized controlled trial^31^ |
| Study locations | 21 studies (84 %) had participants from 37 high-income countries; including Andorra^22^, Australia^20,22^, Austria^22^, Brunei Darussalam^22^, Belgium^22,37^, Canada^20,22^, Croatia^22,37^, Czech Republic^37^, Denmark^20,22,37^, Estonia^22,37^, Finland^37^, France^17,22,31,37^, Germany^1,14,17,22,34,37^, Greece^22,37^, Ireland^22^, Israel^37^, Italy^17,22,37^, Japan^22^, Kuwait^22^, Latvia^37^, Lithuania^37^, Luxembourg^22^, Malta^37^, Monaco^22^, Netherlands^37^, New Zealand^20^, Norway^22,37^, Poland^37^, Portugal^22^, Saudi Arabia^22^, Singapore^22,33^, Slovenia^37^, Spain^22,37^, Sweden^22,37^, Switzerland^18,22,37^, The United States^2,14,19-23,25,26,29,32,35,36,38,39^ and United Kingdom^17,22,37^ |
|  | 5 studies (20%) had participants from 24 upper-middle-income countries; including Albania^22^, Albania^37^, Argentina^22^,Armenia^22^, Belarus^18^, Bosnia and Herzegovina^18^, Botswana^16^, Brazil^22^, Bulgaria^22,37^, Colombia^22^, Georgia^18,22^, Guatemala^22^, Kosovo^18,22^, Lebanon^22^, Malaysia^22,28^, Mexico^22^, Montenegro^18^, Romania^22^, the Russian Federation (Russia)^18,22^, Serbia^18,22^, South Africa^22^, The former Yugoslav Republic of Macedonia^18^, Turkey^18,22,37^ and Venezuela^22^* |
|  | 4 studies (16%) had participants from 14 lower-middle-income countries; including Bangladesh^22^, Bolivia^22^, Cambodia^16^, Egypt^22^, Ghana^22^, India^22^, Iran^22^, Kenya^22^, Lao PDR^16^, Myanmar^16^, Nigerian^15,22^, Pakistan^22^, Tunisia^22^ and Ukraine^18,22^ |
|  | 1 study (4%) had participants from 3 low-income countries; including DR Congo^16^, Ethiopia^16^ and Guinea^16^ |
|  | 1 study (4%) had no information of countries of participants^27^ |

* Venezuela is classified as an upper-middle-income country based on the historical classification in 2019-2020. Venezuela has been temporarily unclassified since July 2021 pending release of revised national accounts statistics.^6^

**Appendix S8. Quality assessment of studies included in the systematic review**

| Studies (Author/Date)* | Was the research design appropriate to address the aims of the research? | Was the recruitment strategy appropriate to the aims of the research? | Was the data collected in a way that addressed the research issue? | Was the data analysis sufficiently rigorous? | Has the relationship between researcher and participants been adequately considered? | Risk of bias (Low/  medium/  high) |
| --- | --- | --- | --- | --- | --- | --- |
|  | Q1: | Q2: | Q3: | Q4: | Q5: |  |
| Gross, P. A. (1988) | Yes | Yes | Yes | Unclear | Yes | High |
| Salluzzo, R (1991) | Yes | Yes | Yes | Unclear | Yes | High |
| Tabriz, M. S. (2004) | Yes | Yes | Yes | Unclear | Yes | Medium |
| Parada, J. P. (2005) | Yes | Yes | Yes | Unclear | Yes | Medium |
| Falagas, M. E. (2008) | Yes | Yes | Yes | Unclear | Yes | Medium |
| Kerur, B. (2012) | Yes | Yes | Yes | Yes | Yes | Low |
| Chew, K. S. (2013) | Yes | Yes | Yes | Unclear | Yes | Medium |
| Heine, D. (2013) | Yes | Yes | Yes | Unclear | Yes | High |
| Ojide, C. K. (2013) | Yes | Yes | Yes | Unclear | Yes | Medium |
| Schmitz, R.P. (2013) | Yes | Yes | Yes | Unclear | Yes | Medium |
| Pavese, P. (2014) | Yes | Yes | Yes | Yes | Yes | Low |
| Kurowski, M. E. (2015) | Yes | Yes | Yes | Unclear | Yes | Medium |
| Ong, I. (2015) | Yes | Yes | Yes | Unclear | Yes | Medium |
| She, R. C. (2015) | Yes | Yes | Yes | Unclear | Yes | Medium |
| Casu, S. (2016) | Yes | Yes | Yes | Yes | Yes | Low |
| Sloane, A. J. (2016) | Yes | Yes | Yes | Unclear | Yes | Medium |
| Donner, L. M. (2017) | Yes | Yes | Yes | Yes | Yes | Low |
| Raupach-Rosin, H. (2017) | Yes | Yes | Yes | Unclear | Yes | Medium |
| Diallo, K. (2018) | Yes | Yes | Yes | Unclear | Yes | Medium |
| Garcia, R. A. (2018) | Yes | Yes | Yes | Yes | Yes | Low |
| WHO CAESAR (2018) | Yes | Yes | Yes | Unclear | Yes | Medium |
| Dailey, P. J. (2019) | Yes | Yes | Yes | Yes | Yes | Low |
| Idelevich, E. A. (2019) | Yes | Yes | Yes | Yes | Yes | Low |
| ThΘ, T. (2019) | Yes | Yes | Yes | Yes | Yes | Low |
| Tran, P. (2020) | Yes | Yes | Yes | Unclear | Yes | Medium |

* Study type was categorized and quality assessment of the evidence was performed using the Mixed Methods Appraisal Tool (MMAT).^30^

**Appendix S9: Barriers and enablers of the 14 TDF domains identified in HICs and LMICs derived from the systematic review**

| **TDF domains** | **High-income countries (HICs)** | | | | **Low and middle-income countries** | | | |
| --- | --- | --- | --- | --- | --- | --- | --- | --- |
|  | **Frequency (N=study)** | **Barriers** | **Enablers** | **Mixed*** | **Frequency (N=study)** | **Barriers** | **Enablers** | **Mixed*** |
| Knowledge | 14 | 4 | 6 | 4 | 5 | 1 | 1 | 3 |
| Goals | 3 | 2 | 1 | 0 | Not evaluated | Not evaluated | Not evaluated | Not evaluated |
| Intentions | 1 | 0 | 0 | 1 | 1 | 1 | 0 | 0 |
| Social professional role and identity | 2 | 1 | 1 | 0 | Not evaluated | Not evaluated | Not evaluated | Not evaluated |
| Social influences | 6 | 1 | 5 | 0 | 1 | 0 | 1 | 0 |
| Environmental context and resources | 9 | 2 | 2 | 5 | 4 | 1 | 0 | 3 |
| Behavioural regulation | 12 | 1 | 5 | 6 | 4 | 1 | 1 | 2 |
| Reinforcement | Not evaluated | Not evaluated | Not evaluated | Not evaluated | Not evaluated | Not evaluated | Not evaluated | Not evaluated |
| Beliefs about consequences | 4 | 3 | 0 | 1 | 3 | 2 | 0 | 1 |
| Memory, attention and decision processes | 9 | 0 | 7 | 2 | 3 | 1 | 2 | 0 |
| Optimism | Not evaluated | Not evaluated | Not evaluated | Not evaluated | Not evaluated | Not evaluated | Not evaluated | Not evaluated |
| Skills | 1 | 0 | 0 | 1 | Not evaluated | Not evaluated | Not evaluated | Not evaluated |
| Beliefs about capabilities | 1 | 0 | 1 | 0 | Not evaluated | Not evaluated | Not evaluated | Not evaluated |
| Emotion | Not evaluated | Not evaluated | Not evaluated | Not evaluated | Not evaluated | Not evaluated | Not evaluated | Not evaluated |

*Mixed = Mixed barriers and enablers

**Appendix S10. Criteria and rank of TDF domains for Indonesia, Thailand and Vietnam**

| **TDF domains** | (1) ‘frequency’ or number studies that identified each domain obtained from the systematic review * | (2) ‘elaboration’ or number of themes within each domain from the systematic review and the TDF survey | (3) ‘expressed importance’ (either a statement from the authors’ interpretation or direct quotes from study respondents expressing importance from the systematic review and the TDF survey | (4) ‘association between reported barriers or enablers and BC practice’ from the TDF survey ** | Overall rank *** |
| --- | --- | --- | --- | --- | --- |
| Knowledge | 14 | 3 | Highly important | Strongly associated | Very important |
| Goals | 3 | 1 | Highly important | Strongly associated | Very important |
| Intentions | 1 | 1 | Highly Important | Strongly associated | Very important |
| Social professional role and identity | 2 | 4 | Highly important | Strongly associated | Very important |
| Social influences | 6 | 2 | Highly important | Strongly associated | Very important |
| Environmental context and resources | 9 | 3 | Highly important | Strongly associated | Very important |
| Behavioural regulation | 12 | 2 | Highly important | Not observed | Very important |
| Reinforcement | Not evaluated | 2 | Highly important | Strongly associated | Very important |
| Beliefs about consequences | 4 | 2 | Important | Strongly associated | Important |
| Memory, attention and decision processes | 9 | 2 | Important | Strongly associated | Important |
| Optimism | Not evaluated | 1 | Important | Strongly associated | Important |
| Skills | 1 | 1 | Important | Associated | Important |
| Beliefs about capabilities | 1 | 2 | Important | Not observed | Important |
| Emotion | Not evaluated | 2 | Important | Not observed | Important |

* Number of respondents that identified each domain obtained from the TDF survey are presented in Appendix S11. ** Details are presented in Appendix S12. Both size of effect (OR) and p values are considered.^40,41^ P values <0.05 was not used as a simple cutoff whether an association was present or absent. P values less than 0.001 is regarded as providing strong evidence against the null hypothesis. The terms ‘strongly associated’, ‘associated’ and ‘(association was) not observed’ were used to summarized overall presentation of the rating of the criterion. *** Overall rank was decided based on detailed presentation of the ratings of each criterion.

**Appendix S11. Results of the TDF survey in Indonesia, Thailand and Vietnam**

| **Questions** | **Indonesia**  **(n=503)** | **Thailand**  **(n=304)** | **Viet Nam**  **(n=501)** | **P value** |
| --- | --- | --- | --- | --- |
| **Type of hospitals (Q1-1)** |  |  |  |  |
| Government hospital | 340 (67.6%) | 209 (68.8%) | 431 (86.0%) | <0.001 |
| Private hospital | 113 (22.5%) | 15 (4.9%) | 17 (3.4%) |  |
| University hospital | 26 (5.2%) | 76 (25.0%) | 29 (5.8%) |  |
| Other^1^ | 19 (3.8%) | 2 (0.7%) | 22 (4.4%) |  |
| I do not want to answer | 5 (1.0%) | 2 (0.7%) | 2 (0.4%) |  |
| **Case-study: Would you take BC sample from a hypothetical sepsis case? (Q1-3)** |  |  |  |  |
| Definitely (>95-100% of the time) | 157 (31.2%) | 273 (89.8%) | 252 (50.3%) | <0.001 |
| Likely (75-95% of the time) | 138 (27.4%) | 23 (7.6%) | 149 (29.7%) |  |
| Maybe (25-74% of the time) | 116 (23.1%) | 5 (1.6%) | 70 (14.0%) |  |
| Unlikely (5-24% of the time) | 44 (8.7%) | 2 (0.7%) | 19 (3.8%) |  |
| Rarely (ranging from never <5% of the time) | 46 (9.1%) | 1 (0.3%) | 9 (1.8%) |  |
| I do not know | 1 (0.2%) | 0 (0%) | 1 (0.2%) |  |
| I do not want to answer | 1 (0.2%) | 0 (0%) | 1 (0.2%) |  |
| **Knowledge (TDF-1): Do you know of any guideline(s) or guideline(s) used in my hospital (Q1-4)?** |  |  |  |  |
| Yes | 240 (47.7%) | 169 (55.6%) | 347 (69.3%) | <0.001 |
| No, my hospital does not have any | 68 (13.5%) | 33 (10.9%) | 49 (9.8%) |  |
| No, I do not know if my hospital uses any | 183 (36.4%) | 98 (32.2%) | 95 (19.0%) |  |
| I do not want to answer | 12 (2.4%) | 4 (1.3%) | 10 (2.0%) |  |
| **Knowledge (TDF-1): known local guideline among those who answered that they know of local guideline (Q1-5)** |  |  |  |  |
| All patients presenting with SIRS | 155/240 (64.6%) | 147/169 (87.0%) | 218/347 (62.8%) | <0.001 |
| All patients presenting with sepsis | 183/240 (76.2%) | 138/169 (81.7%) | 291/347 (83.9%) | 0.07 |
| All patients presenting with septic shock | 147/240 (61.3%) | 131/169 (77.5%) | 270/347 (77.8%) | <0.001 |
| All patients starting parenteral antibiotic treatment | 92/240 (38.3%) | 92/169 (54.4%) | 73/347 (21.0%) | <0.001 |
| All patients with no clinical improvement after receiving empirical antibiotics | 141/240 (58.7%) | 99/169 (58.6%) | 160/347 (46.1%) | 0.003 |
| All patients presenting with infection and having underlying diseases | 76/240 (31.7%) | 61/169 (36.1%) | 94/347 (27.1%) | 0.10 |
| All patients with chronic fever | 97/240 (40.4%) | 87/169 (51.5%) | 208/347 (59.9%) | <0.001 |
| All patients with fever of unknown origins | 114/240 (47.5%) | 100/169 (59.2%) | 185/347 (53.3%) | 0.06 |
| All patients suspected of infections caused by atypical organisms | 97/240 (40.4%) | 74/169 (43.8%) | 94/347 (27.1%) | <0.001 |
| All patients suspected of infections caused by antimicrobial-resistant organisms | 131/240 (54.6%) | 96/169 (56.8%) | 168/347 (48.4%) | 0.14 |
| All patients suspected of infections caused by multiple-drug-resistant organisms | 136/240 (56.7%) | 103/169 (60.9%) | 194/347 (55.9%) | 0.54 |
| All patients suspected of hospital-acquired infections | 116/240 (48.3%) | 99/169 (58.6%) | 184/347 (53.0%) | 0.12 |
| **Intention (TDF-8): How often do you plan to follow the local guideline among those who answered that they know of local guideline (Q1-6)?** |  |  |  |  |
| All the time (>95-100% of the cases) | 70/240 (29.2%) | 76/169 (45.0%) | 88/347 (25.4%) | <0.001 |
| Often (75-95% of the cases) | 102/240 (42.5%) | 81/169 (47.9%) | 195/347 (56.2%) |  |
| Moderately (25-74% of the cases) | 33/240 (13.8%) | 11/169 (6.5%) | 49/347 (14.1%) |  |
| Occasionally (5-24% of the cases) | 16/240 (6.7%) | 0/169 (0%) | 11/347 (3.2%) |  |
| Rarely (ranging from never <5% of the cases) | 11/240 (4.6%) | 1/169 (0.6%) | 2/347 (0.6%) |  |
| I do not know | 7/240 (2.9%) | 0/169 (0%) | 2/347 (0.6%) |  |
| I do not want to answer | 1/240 (0.4%) | 0/169 (0%) | 0/347 (0%) |  |
| **Memory, attention and decision processes (TDF-10): any additional reasons for deciding to do BC among those who answered that they know of local guideline (Q1-7)?** |  |  |  |  |
| No additional reasons | 77/240 (32.1%) | 35/169 (20.7%) | 110/347 (31.7%) | 0.02 |
| Patients presenting with chills | 15/240 (6.3%) | 39/169 (23.1%) | 23/347 (6.6%) | <0.001 |
| Patients presenting with sepsis | 102/240 (42.5%) | 101/169 (59.8%) | 113/347 (32.6%) | <0.001 |
| Patients presenting with septic shock | 86/240 (35.8%) | 96/169 (56.8%) | 139/347 (40.1%) | <0.001 |
| Patients starting parenteral antibiotic treatment | 48/240 (20.0%) | 59/169 (34.9%) | 35/347 (10.1%) | <0.001 |
| Patient with no clinical improvement after receiving empirical antibiotics | 102/240 (42.5%) | 75/169 (44.4%) | 97/347 (28.0%) | <0.001 |
| Patients with infection and having underlying diseases | 42/240 (17.5%) | 36/169 (21.3%) | 56/347 (16.1%) | 0.35 |
| Patients presenting with chronic fever | 54/240 (22.5%) | 55/169 (32.5%) | 107/347 (30.8%) | 0.04 |
| Patients presenting with fever of unknown origin | 72/240 (30.0%) | 63/169 (37.3%) | 96/347 (27.7%) | 0.08 |
| Patients suspected of infections caused by atypical organisms | 52/240 (21.7%) | 46/169 (27.2%) | 48/347 (13.8%) | 0.001 |
| Patients suspected of infections caused by antimicrobial-resistant organisms | 77/240 (32.1%) | 53/169 (31.4%) | 86/347 (24.8%) | 0.10 |
| Patients suspected of infections caused by multiple-drug-resistant organisms | 82/240 (34.2%) | 63/169 (37.3%) | 92/347 (26.5%) | 0.03 |
| Patients suspected of hospital-acquired infections | 77/240 (32.1%) | 59/169 (34.9%) | 97/347 (28.0%) | 0.24 |
| Laboratory results showing leukocytosis | 29/240 (12.1%) | 42/169 (24.9%) | 25/347 (7.2%) | <0.001 |
| Laboratory results showing neutropenia | 36/240 (15.0%) | 54/169 (32.0%) | 28/347 (8.1%) | <0.001 |
| Laboratory results showing left shift in blood count | 31/240 (12.9%) | 26/169 (15.4%) | 14/347 (4.0%) | <0.001 |
| Laboratory results showing CRP increase | 37/240 (15.4%) | 22/169 (13.0%) | 42/347 (12.1%) | 0.51 |
| Laboratory results showing procalcitonin increase | 55/240 (22.9%) | 22/169 (13.0%) | 94/347 (27.1%) | 0.002 |
| Patients can afford the cost of BC | 25/240 (10.4%) | 9/169 (5.3%) | 32/347 (9.2%) | 0.18 |
| Patients have a health scheme or insurance that covers the cost of BC | 24/240 (10.0%) | 8/169 (4.7%) | 26/347 (7.5%) | 0.14 |
| Patients are likely to have a final diagnosis that includes the cost of BC in the package of fee for service | 18/240 (7.5%) | 0/169 (0%) | 25/347 (7.2%) | 0.001 |
| **Memory, attention and decision processes (TDF-10): any reasons for deciding to do BC among those who did not answer that they know of local guideline (Q1-8)?** |  |  |  |  |
| Patients presenting with chills | 20/263 (7.6%) | 49/135 (36.3%) | 29/154 (18.8%) | <0.001 |
| Patients presenting with sepsis | 188/263 (71.5%) | 132/135 (97.8%) | 109/154 (70.8%) | <0.001 |
| Patients presenting with septic shock | 165/263 (62.7%) | 128/135 (94.8%) | 135/154 (87.7%) | <0.001 |
| Patients starting parenteral antibiotic treatment | 48/263 (18.3%) | 95/135 (70.4%) | 26/154 (16.9%) | <0.001 |
| Patient with no clinical improvement after receiving empirical antibiotics | 188/263 (71.5%) | 119/135 (88.1%) | 84/154 (54.5%) | <0.001 |
| Patients with infection and having underlying diseases | 85/263 (32.3%) | 79/135 (58.5%) | 52/154 (33.8%) | <0.001 |
| Patients presenting with chronic fever | 91/263 (34.6%) | 89/135 (65.9%) | 108/154 (70.1%) | <0.001 |
| Patients presenting with fever of unknown origin | 138/263 (52.5%) | 110/135 (81.5%) | 100/154 (64.9%) | <0.001 |
| Patients suspected of infections caused by atypical organisms | 123/263 (46.8%) | 81/135 (60.0%) | 55/154 (35.7%) | <0.001 |
| Patients suspected of infections caused by antimicrobial-resistant organisms | 177/263 (67.3%) | 108/135 (80.0%) | 85/154 (55.2%) | <0.001 |
| Patients suspected of infections caused by multiple-drug-resistant organisms | 183/263 (69.6%) | 113/135 (83.7%) | 85/354 (24.0%) | <0.001 |
| Patients suspected of hospital-acquired infections | 136/263 (51.7%) | 107/135 (79.3%) | 78/154 (50.6%) | <0.001 |
| Laboratory results showing leukocytosis | 41/263 (15.6%) | 52/135 (38.5%) | 15/154 (9.7%) | <0.001 |
| Laboratory results showing neutropenia | 34/263 (12.9%) | 59/135 (43.7%) | 18/154 (11.7%) | <0.001 |
| Laboratory results showing left shift in blood count | 47/263 (17.9%) | 47/135 (34.8%) | 16/154 (10.4%) | <0.001 |
| Laboratory results showing CRP increase | 59/263 (22.4%) | 23/135 (17.0%) | 26/154 (16.9%) | 0.27 |
| Laboratory results showing procalcitonin increase | 73/263 (27.8%) | 28/135 (20.7%) | 53/154 (34.4%) | 0.04 |
| Patients can afford the cost of BC | 81/263 (30.8%) | 18/135 (13.3%) | 32/154 (20.8%) | <0.001 |
| Patients have a health scheme or insurance that covers the cost of BC | 88/263 (33.5%) | 19/135 (14.1%) | 31/154 (20.1%) | <0.001 |
| Patients are likely to have a final diagnosis that includes the cost of BC in the package of fee for service | 51/263 (19.4%) | 0/135 (0%) | 30/154 (19.5%) | <0.001 |
| **Knowledge (TDF-1):** **Do you know of any international guideline(s) or guideline(s) (Q1-9)?** |  |  |  |  |
| Yes | 229 (45.5%) | 142 (46.7%) | 225 (44.9%) | <0.001 |
| No | 263 (52.3%) | 156 (51.3%) | 233 (46.5%) |  |
| I do not want to answer | 11 (2.2%) | 6 (2.0%) | 43 (8.6%) |  |
| **Knowledge (TDF-1): known international guideline or guideline among those who answered that they know of any international guideline(s) or guideline(s) (Q1-10)** |  |  |  |  |
| BC sampling in all patients presenting with sepsis | 220/229 (96.1%) | 138/142 (97.2%) | 208/225 (92.4%) | 0.08 |
| BC sampling in all patients starting parenteral antibiotic treatment | 125/229 (54.6%) | 87/142 (61.3%) | 147/225 (65.3%) | <0.001 |
| **Professional role (Q2-1): Current job** |  |  |  |  |
| Medical doctor – an executive level | 13 (2.6%) | 5 (1.6%) | 17 (3.4%) | <0.001 |
| Medical doctor – a consultant level | 74 (14.7%) | 75 (24.7%) | 198 (39.5%) |  |
| Medical doctor – a general physician level | 124 (24.7%) | 38 (12.5%) | 112 (22.4%) |  |
| Medical doctor – a resident/registra/fellow level | 168 (33.4%) | 63 (20.7%) | 101 (20.2%) |  |
| Intern – recent medical school graduate | 33 (6.6%) | 35 (11.5%) | 14 (2.8%) |  |
| Final-year medical student | 91 (18.1%) | 88 (28.9%) | 59 (11.8%) |  |
| **Professional role (Q2-2): Which types of professionals/staff can order or initiate an order for a BC?** |  |  |  |  |
| Medical doctor – an executive level | 61 (12.1%) | 163 (53.6%) | 59 (11.8%) | <0.001 |
| Medical doctor – a consultant level | 431 (85.7%) | 250 (82.2%) | 439 (87.6%) | 0.11 |
| Medical doctor – a general physician level | 265 (52.7%) | 240 (78.9%) | 347 (69.3%) | <0.001 |
| Medical doctor – a resident (postgrad training) level | 268 (53.3%) | 242 (79.6%) | 317 (63.3%) | <0.001 |
| Intern – a recent medical school graduate level | 83 (16.5%) | 231 (76.0%) | 118 (23.6%) | <0.001 |
| Final-year medical student | 11 (2.2%) | 87 (28.6%) | 3 (0.6%) | <0.001 |
| I do not want to answer | 3 (0.6%) | 1 (0.3%) | 11 (2.2%) | 0.03 |
| Other | 0 (0%) | 0 (0%) | 0 (0%) | >0.99 |
| **Knowledge (TDF-1): Do you know when and which patients should receive an order for a BC in your hospital (Q2-3)?** |  |  |  |  |
| Definitely (>95-100% of the case) | 65 (12.9%) | 106 (34.9%) | 72 (14.4%) | <0.001 |
| Likely (75-95% of the case) | 200 (39.8%) | 168 (55.3%) | 245 (48.9%) |  |
| Uncertain (25-74% of the case) | 148 (29.4%) | 28 (9.2%) | 128 (25.5%) |  |
| Unlikely (5-24% of the case) | 59 (11.7%) | 0 (0%) | 31 (6.2%) |  |
| Rarely (ranging from never <5% of the case) | 19 (3.8%) | 0 (0%) | 6 (1.2%) |  |
| I do not know | 10 (2.0%) | 1 (0.3%) | 8 (1.6%) |  |
| I do not want to answer | 2 (0.4%) | 1 (0.3%) | 11 (2.2%) |  |
| **Social professional role and identity (TDF-3): Is it an appropriate part of your current job to order a BC (Q2-4)?** |  |  |  |  |
| Very appropriate | 119 (23.7%) | 103 (33.9%) | 110 (22.0%) | <0.001 |
| Appropriate | 232 (46.1%) | 166 (54.6%) | 290 (57.9%) |  |
| Uncertain | 62 (12.3%) | 20 (6.6%) | 48 (9.6%) |  |
| Inappropriate | 21 (4.2%) | 2 (0.7%) | 12 (2.4%) |  |
| Very inappropriate | 2 (0.4%) | 0 (0%) | 0 (0%) |  |
| I do not know | 10 (2.0%) | 0 (0%) | 0 (0%) |  |
| I do not want to answer | 2 (0.4%) | 0 (0%) | 19 (3.8%) |  |
| I cannot order BC. It is not part of my job | 55 (10.9%) | 13 (4.3%) | 22 (4.4%) |  |
| **Social professional role and identity (TDF-3): Would it be an appropriate part of your current job to order a BC among those who answered that they cannot order for a BC (Q2-5)?** |  |  |  |  |
| Very appropriate | 4/55 (7.3%) | 0/13 (0%) | 0/22 (0%) | 0.009 |
| Appropriate | 19/55 (34.5%) | 8/13 (61.5%) | 4/22 (18.2%) |  |
| Uncertain | 10/55 (18.2%) | 4/13 (30.8%) | 2/22 (9.1%) |  |
| Inappropriate | 15/55 (27.3%) | 1/13 (7.7%) | 8/22 (36.4%) |  |
| Very inappropriate | 3/55 (5.5%) | 0/13 (0%) | 2/22 (9.1%) |  |
| I do not know | 4/55 (7.3%) | 0/13 (0%) | 2/22 (9.1%) |  |
| I do not want to answer | 0/55 (0%) | 0/13 (0%) | 4/22 (18.2%) |  |
| **Professional role (Q2-6): Which types of professionals/staff are tasked to draw blood from patients for BC?** |  |  |  |  |
| Medical doctor – executive level | 12 (2.4%) | 44 (14.5%) | 23 (4.6%) | <0.001 |
| Medical doctor – a consultant level | 60 (11.9%) | 90 (29.6%) | 152 (30.3%) | 0.11 |
| Medical doctor – a general physician level | 72 (14.3%) | 105 (34.5%) | 129 (25.7%) | <0.001 |
| Medical doctor – a resident level | 96 (19.1%) | 122 (40.1%) | 113 (22.6%) | <0.001 |
| Intern – recent medical school graduate | 39 (7.8%) | 105 (34.5%) | 85 (17.0%) | <0.001 |
| Final-year medical student | 27 (5.4%) | 99 (32.6%) | 25 (5.0%) | <0.001 |
| Registered nurses | 342 (68.0%) | 215 (70.7%) | 392 (78.2%) | 0.001 |
| Microbiology laboratory team | 227 (45.1%) | 91 (29.9%) | 151 (30.1%) | <0.001 |
| Specialized blood draw team | 197 (39.2%) | 91 (29.9%) | 69 (13.8%) | <0.001 |
| I do not want to answer | 3 (0.6%) | 0 (0%) | 2 (0.4%) | 0.41 |
| **Social professional role and identity (TDF-3): Is it an appropriate part of your current job to draw blood (Q2-7)?** |  |  |  |  |
| Very appropriate | 34 (6.8%) | 36 (11.8%) | 49 (9.8%) | 0.01 |
| Appropriate | 179 (35.6%) | 102 (33.6%) | 179 (35.7%) |  |
| Uncertain | 109 (21.7%) | 52 (17.1%) | 68 (13.6%) |  |
| Inappropriate | 89 (17.7%) | 46 (15.1%) | 85 (17.0%) |  |
| Very inappropriate | 7 (1.4%) | 6 (2.0%) | 3 (0.6%) |  |
| I do not know | 8 (1.6%) | 4 (1.3%) | 4 (0.8%) |  |
| I do not want to answer | 4 (0.8%) | 1 (0.3%) | 4 (0.8%) |  |
| It is not part of my job to draw blood | 73 (14.5%) | 57 (18.8%) | 109 (21.8%) |  |
| **Skill (TDF-2): How skilled are you in drawing blood excluding those whose jobs did not include drawing blood (Q2-8)?** |  |  |  |  |
| Very good skill | 18/430 (4.2%) | 12/247 (4.9%) | 32/392 (8.2%) | <0.001 |
| Good skill | 138/430 (32.1%) | 46/247 (18.6%) | 112/392 (28.6%) |  |
| Fair skill | 202/430 (47.0%) | 118/247 (47.8%) | 196/392 (50.0%) |  |
| Poor skill | 20/430 (4.7%) | 52/247 (21.1%) | 33/392 (8.4%) |  |
| Very poor skill | 4/430 (0.9%) | 16/247 (6.5%) | 1/392 (0.3%) |  |
| I do not know | 39/430 (9.1%) | 3/247 (1.2%) | 11/392 (2.8%) |  |
| I do not want to answer | 9/430 (2.1%) | 0/247 (0%) | 7/392 (1.8%) |  |
| **Beliefs about capabilities (TDF-4): How confident that you can draw blood successfully excluding those whose jobs did not include drawing blood (Q2-9)?** |  |  |  |  |
| Strongly confident | 32/430 (7.4%) | 20/247 (8.1%) | 42/392 (10.7%) | <0.001 |
| Confident | 271/430 (63.0%) | 93/247 (37.7%) | 231/392 (58.9%) |  |
| Uncertain | 74/430 (17.2%) | 81/247 (32.8%) | 90/392 (23.0%) |  |
| Doubtful | 42/430 (9.8%) | 34/247 (13.8%) | 22/392 (5.6%) |  |
| Strongly doubtful | 2/430 (0.5%) | 19/247 (7.7%) | 6/392 (1.5%) |  |
| I do not know | 4/430 (0.9%) | 0/247 (0%) | 0/392 (0%) |  |
| I do not want to answer | 5/430 (1.2%) | 0/247 (0%) | 1/392 (0.3%) |  |
| **Beliefs about capabilities (TDF-4): How confident that you can draw blood appropriately excluding those whose jobs did not include drawing blood (Q2-10)?** |  |  |  |  |
| Strongly confident | 28/430 (6.5%) | 30/247 (12.1%) | 37/392 (9.4%) | <0.001 |
| Confident | 262/430 (60.9%) | 109/247 (44.1%) | 222/392 (56.6%) |  |
| Uncertain | 86/430 (20.0%) | 61/247 (24.7%) | 109/392 (27.8%) |  |
| Doubtful | 44/430 (10.2%) | 33/247 (13.4%) | 17/392 (4.3%) |  |
| Strongly doubtful | 3/430 (0.7%) | 11/247 (4.5%) | 2/392 (0.5%) |  |
| I do not know | 3/430 (0.7%) | 1/247 (0.4%) | 1/392 (0.3%) |  |
| I do not want to answer | 4/430 (0.9%) | 2/247 (0.8%) | 4/392 (1.0%) |  |
| **Beliefs about capabilities (TDF-4): Are you confident that others can draw blood successfully (Q2-11)?** |  |  |  |  |
| Strongly confident | 99 (19.7%) | 106 (34.9%) | 71 (14.2%) | <0.001 |
| Confident | 366 (72.8%) | 176 (57.9%) | 333 (66.5%) |  |
| Uncertain | 17 (3.4%) | 14 (4.6%) | 88 (17.6%) |  |
| Doubtful | 16 (3.2%) | 7 (2.3%) | 6 (1.2%) |  |
| Strongly doubtful | 0 (0%) | 0 (0%) | 1 (0.2%) |  |
| I do not know | 2 (0.4%) | 1 (0.3%) | 1 (0.2%) |  |
| I do not want to answer | 3 (0.6%) | 0 (0%) | 1 (0.2%) |  |
| **Beliefs about capabilities (TDF-4): Are you confident that others can draw blood appropriately (Q2-12)?** |  |  |  |  |
| Strongly confident | 86 (17.1%) | 66 (21.7%) | 45 (9.0%) | <0.001 |
| Confident | 342 (68.0%) | 184 (60.5%) | 273 (54.5%) |  |
| Uncertain | 42 (8.3%) | 45 (14.8%) | 170 (33.9%) |  |
| Doubtful | 26 (5.2%) | 6 (2.0%) | 8 (1.6%) |  |
| Strongly doubtful | 1 (0.2%) | 2 (0.7%) | 2 (0.4%) |  |
| I do not know | 4 (0.8%) | 1 (0.3%) | 1 (0.2%) |  |
| I do not want to answer | 2 (0.4%) | 0 (0%) | 2 (0.4%) |  |
| **Optimism (TDF-5): how optimistic are you that a BC will be sampled and processed in the laboratory appropriately (Q2-13)?** |  |  |  |  |
| Strongly optimistic | 70 (13.9%) | 38 (12.5%) | 31 (6.2%) | <0.001 |
| Optimistic | 332 (66.0%) | 225 (74.0%) | 338 (67.5%) |  |
| Neither optimistic nor pessimistic | 74 (14.7%) | 31 (10.2%) | 124 (24.8%) |  |
| Pessimistic | 8 (1.6%) | 4 (1.3%) | 4 (0.8%) |  |
| Strongly pessimistic | 5 (1.0%) | 0 (0%) | 1 (0.2%) |  |
| I do not know | 10 (2.0%) | 5 (1.6%) | 2 (0.4%) |  |
| I do not want to answer | 4 (0.8%) | 1 (0.3%) | 1 (0.2%) |  |
| **Beliefs about consequence (TDF-6): BC is helpful in clinical decisions (Q3-1-1).** |  |  |  |  |
| Strongly agree | 204 (40.6%) | 153 (50.3%) | 194 (38.7%) | <0.001 |
| Agree | 279 (55.5%) | 144 (47.4%) | 246 (49.1%) |  |
| Uncertain | 13 (2.6%) | 6 (2.0%) | 47 (9.4%) |  |
| Disagree | 4 (0.8%) | 1 (0.3%) | 11 (2.2%) |  |
| Strongly disagree | 0 (0%) | 0 (0%) | 1 (0.2%) |  |
| I do not know | 2 (0.4%) | 0 (0%) | 0 (0%) |  |
| I do not want to answer | 1 (0.2%) | 0 (0%) | 2 (0.4%) |  |
| **Beliefs about consequence (TDF-6): BC is helpful to rule in an infection (Q3-1-2).** |  |  |  |  |
| Strongly agree | 192 (38.2%) | 123 (40.5%) | 162 (32.3%) | <0.001 |
| Agree | 276 (54.9%) | 159 (52.3%) | 260 (51.9%) |  |
| Uncertain | 14 (2.8%) | 10 (3.3%) | 51 (10.2%) |  |
| Disagree | 18 (3.6%) | 7 (2.3%) | 24 (4.8%) |  |
| Strongly disagree | 0 (0%) | 1 (0.3%) | 2 (0.4%) |  |
| I do not know | 2 (0.4%) | 4 (1.3%) | 0 (0%) |  |
| I do not want to answer | 1 (0.2%) | 0 (0%) | 2 (0.4%) |  |
| **Beliefs about consequence (TDF-6): BC is helpful to rule out an infection (Q3-1-3).** |  |  |  |  |
| Strongly agree | 137 (27.2%) | 72 (23.7%) | 59 (11.8%) | <0.001 |
| Agree | 258 (51.3%) | 97 (31.9%) | 163 (32.5%) |  |
| Uncertain | 44 (8.7%) | 32 (10.5%) | 126 (25.1%) |  |
| Disagree | 56 (11.1%) | 79 (26.0%) | 127 (25.3%) |  |
| Strongly disagree | 5 (1.0%) | 22 (7.2%) | 23 (4.6%) |  |
| I do not know | 2 (0.4%) | 2 (0.7%) | 0 (0%) |  |
| I do not want to answer | 1 (0.2%) | 0 (0%) | 3 (0.6%) |  |
| **Beliefs about consequence (TDF-6): BC is helpful in detecting AMR infections (Q3-1-4).** |  |  |  |  |
| Strongly agree | 267 (53.1%) | 147 (48.4%) | 154 (30.7%) | <0.001 |
| Agree | 219 (43.5%) | 140 (46.1%) | 272 (54.3%) |  |
| Uncertain | 10 (2.0%) | 11 (3.6%) | 51 (10.2%) |  |
| Disagree | 4 (0.8%) | 4 (1.3%) | 18 (3.6%) |  |
| Strongly disagree | 0 (0%) | 1 (0.3%) | 4 (0.8%) |  |
| I do not know | 2 (0.4%) | 1 (0.3%) | 1 (0.2%) |  |
| I do not want to answer | 1 (0.2%) | 0 (0%) | 1 (0.2%) |  |
| **Beliefs about consequence (TDF-6): BC is helpful in adjusting antibiotics (Q3-1-5).** |  |  |  |  |
| Strongly agree | 285 (56.7%) | 172 (56.6%) | 177 (35.3%) | <0.001 |
| Agree | 206 (41.0%) | 128 (42.1%) | 256 (51.1%) |  |
| Uncertain | 9 (1.8%) | 2 (0.7%) | 40 (8.0%) |  |
| Disagree | 0 (0%) | 1 (0.3%) | 21 (4.2%) |  |
| Strongly disagree | 1 (0.2%) | 1 (0.3%) | 3 (0.6%) |  |
| I do not know | 1 (0.2%) | 0 (0%) | 0 (0%) |  |
| I do not want to answer | 1 (0.2%) | 0 (0%) | 4 (0.8%) |  |
| **Beliefs about consequence (TDF-6): BC can reduce overuse of antibiotics (Q3-1-6).** |  |  |  |  |
| Strongly agree | 241 (47.9%) | 142 (46.7%) | 157 (31.3%) | <0.001 |
| Agree | 220 (43.7%) | 131 (43.1%) | 249 (49.7%) |  |
| Uncertain | 30 (6.0%) | 19 (6.3%) | 59 (11.8%) |  |
| Disagree | 9 (1.8%) | 11 (3.6%) | 30 (6.0%) |  |
| Strongly disagree | 1 (0.2%) | 1 (0.3%) | 4 (0.8%) |  |
| I do not know | 1 (0.2%) | 0 (0%) | 0 (0%) |  |
| I do not want to answer | 1 (0.2%) | 0 (0%) | 2 (0.4%) |  |
| **Beliefs about consequence (TDF-6): BC can reduce length of hospital stay (Q3-1-7).** |  |  |  |  |
| Strongly agree | 167 (33.2%) | 101 (33.2%) | 106 (21.2%) | <0.001 |
| Agree | 215 (42.7%) | 122 (40.1%) | 227 (45.3%) |  |
| Uncertain | 97 (19.3%) | 54 (17.8%) | 124 (24.8%) |  |
| Disagree | 18 (3.6%) | 23 (7.6%) | 39 (7.8%) |  |
| Strongly disagree | 0 (0%) | 2 (0.7%) | 3 (0.6%) |  |
| I do not know | 4 (0.8%) | 1 (0.3%) | 0 (0%) |  |
| I do not want to answer | 2 (0.4%) | 1 (0.3%) | 2 (0.4%) |  |
| **Beliefs about consequence (TDF-6): BC can reduce patient mortality (Q3-1-8).** |  |  |  |  |
| Strongly agree | 178 (35.4%) | 120 (39.5%) | 124 (24.8%) | <0.001 |
| Agree | 228 (45.3%) | 135 (44.4%) | 242 (48.3%) |  |
| Uncertain | 79 (15.7%) | 38 (12.5%) | 98 (19.6%) |  |
| Disagree | 12 (2.4%) | 8 (2.6%) | 31 (6.2%) |  |
| Strongly disagree | 1 (0.2%) | 0 (0%) | 3 (0.6%) |  |
| I do not know | 4 (0.8%) | 3 (1.0%) | 1 (0.2%) |  |
| I do not want to answer | 1 (0.2%) | 0 (0%) | 2 (0.4%) |  |
| **Beliefs about consequence (TDF-6): Accumulative results of BC are helpful in understanding epidemiology of AMR bacterial infections (Q3-1-9).** |  |  |  |  |
| Strongly agree | 237 (47.1%) | 144 (47.4%) | 193 (38.5%) | 0.003 |
| Agree | 247 (49.1%) | 141 (46.4%) | 266 (53.1%) |  |
| Uncertain | 13 (2.6%) | 16 (5.3%) | 32 (6.4%) |  |
| Disagree | 0 (0%) | 1 (0.3%) | 7 (1.4%) |  |
| Strongly disagree | 1 (0.2%) | 0 (0%) | 1 (0.2%) |  |
| I do not know | 4 (0.8%) | 2 (0.7%) | 0 (0%) |  |
| I do not want to answer | 1 (0.2%) | 0 (0%) | 2 (0.4%) |  |
| **Beliefs about consequence (TDF-6): BC is unnecessary because antibiotic therapy can be determined based on clinical presentations (Q3-3-1).** |  |  |  |  |
| Strongly agree | 13 (2.6%) | 7 (2.3%) | 18 (3.6%) | <0.001 |
| Agree | 89 (17.7%) | 48 (15.8%) | 53 (10.6%) |  |
| Uncertain | 154 (30.6%) | 48 (15.8%) | 113 (22.6%) |  |
| Disagree | 199 (39.6%) | 146 (48.0%) | 264 (52.7%) |  |
| Strongly disagree | 42 (8.3%) | 54 (17.8%) | 53 (10.6%) |  |
| I do not know | 6 (1.2%) | 1 (0.3%) | 0 (0%) |  |
| I do not want to answer | 0 (0%) | 0 (0%) | 0 (0%) |  |
| **Beliefs about consequence (TDF-6): The therapeutic consequence of BC sampling is questionable (Q3-3-2).** |  |  |  |  |
| Strongly agree | 12 (2.4%) | 25 (8.2%) | 16 (3.2%) | <0.001 |
| Agree | 82 (16.3%) | 58 (19.1%) | 45 (9.0%) |  |
| Uncertain | 167 (33.2%) | 60 (19.7%) | 123 (24.6%) |  |
| Disagree | 191 (38.0%) | 116 (38.2%) | 275 (54.9%) |  |
| Strongly disagree | 34 (6.8%) | 39 (12.8%) | 34 (6.8%) |  |
| I do not know | 17 (3.4%) | 5 (1.6%) | 2 (0.4%) |  |
| I do not want to answer | 0 (0%) | 1 (0.3%) | 6 (1.2%) |  |
| **Beliefs about consequence (TDF-6): The scientific basis of the guideline on BC is questionable (Q3-3-3).** |  |  |  |  |
| Strongly agree | 9 (1.8%) | 16 (5.3%) | 15 (3.0%) | <0.001 |
| Agree | 45 (8.9%) | 63 (20.7%) | 43 (8.6%) |  |
| Uncertain | 106 (21.1%) | 58 (19.1%) | 141 (28.1%) |  |
| Disagree | 248 (49.3%) | 120 (39.5%) | 254 (50.7%) |  |
| Strongly disagree | 79 (15.7%) | 39 (12.8%) | 41 (8.2%) |  |
| I do not know | 15 (3.0%) | 7 (2.3%) | 4 (0.8%) |  |
| I do not want to answer | 1 (0.2%) | 1 (0.3%) | 3 (0.6%) |  |
| **Beliefs about consequence (TDF-6): BC is unnecessary because results are often delayed (Q3-3-4).** |  |  |  |  |
| Strongly agree | 15 (3.0%) | 8 (2.6%) | 15 (3.0%) | <0.001 |
| Agree | 113 (22.5%) | 31 (10.2%) | 38 (7.6%) |  |
| Uncertain | 119 (23.7%) | 23 (7.6%) | 82 (16.4%) |  |
| Disagree | 212 (42.1%) | 161 (53.0%) | 303 (60.5%) |  |
| Strongly disagree | 36 (7.2%) | 80 (26.3%) | 62 (12.4%) |  |
| I do not know | 8 (1.6%) | 0 (0%) | 0 (0%) |  |
| I do not want to answer | 0 (0%) | 1 (0.3%) | 1 (0.2%) |  |
| **Beliefs about consequence (TDF-6): BC is unnecessary because results are often not interpretable (Q3-3-5).** |  |  |  |  |
| Strongly agree | 7 (1.4%) | 4 (1.3%) | 11 (2.2%) | <0.001 |
| Agree | 46 (9.1%) | 18 (5.9%) | 26 (5.2%) |  |
| Uncertain | 120 (23.9%) | 18 (5.9%) | 70 (14.0%) |  |
| Disagree | 275 (54.7%) | 166 (54.6%) | 326 (65.1%) |  |
| Strongly disagree | 47 (9.3%) | 97 (31.9%) | 67 (13.4%) |  |
| I do not know | 7 (1.4%) | 1 (0.3%) | 0 (0%) |  |
| I do not want to answer | 1 (0.2%) | 0 (0%) | 1 (0.2%) |  |
| **Beliefs about consequence (TDF-6): BC is unnecessary because results are often negative or no growth (Q3-3-6).** |  |  |  |  |
| Strongly agree | 9 (1.8%) | 6 (2.0%) | 11 (2.2%) | <0.001 |
| Agree | 57 (11.3%) | 26 (8.6%) | 39 (7.8%) |  |
| Uncertain | 114 (22.7%) | 37 (12.2%) | 83 (16.6%) |  |
| Disagree | 261 (51.9%) | 149 (49.0%) | 312 (62.3%) |  |
| Strongly disagree | 51 (10.1%) | 85 (28.0%) | 55 (11.0%) |  |
| I do not know | 10 (2.0%) | 1 (0.3%) | 0 (0%) |  |
| I do not want to answer | 1 (0.2%) | 0 (0%) | 1 (0.2%) |  |
| **Beliefs about consequence (TDF-6): BC is unnecessary because cultures are often contaminated (Q3-3-7).** |  |  |  |  |
| Strongly agree | 8 (1.6%) | 6 (2.0%) | 10 (2.0%) | <0.001 |
| Agree | 65 (12.9%) | 23 (7.6%) | 31 (6.2%) |  |
| Uncertain | 166 (33.0%) | 44 (14.5%) | 105 (21.0%) |  |
| Disagree | 212 (42.1%) | 153 (50.3%) | 290 (57.9%) |  |
| Strongly disagree | 39 (7.8%) | 77 (25.3%) | 59 (11.8%) |  |
| I do not know | 12 (2.4%) | 0 (0%) | 1 (0.2%) |  |
| I do not want to answer | 1 (0.2%) | 1 (0.3%) | 5 (1.0%) |  |
| **Beliefs about consequence (TDF-6): BC is unnecessary because results often do not agree with clinical signs (Q3-3-8).** |  |  |  |  |
| Strongly agree | 8 (1.6%) | 5 (1.6%) | 13 (2.6%) | <0.001 |
| Agree | 46 (9.1%) | 22 (7.2%) | 21 (4.2%) |  |
| Uncertain | 147 (29.2%) | 36 (11.8%) | 84 (16.8%) |  |
| Disagree | 249 (49.5%) | 158 (52.0%) | 325 (64.9%) |  |
| Strongly disagree | 43 (8.5%) | 83 (27.3%) | 49 (9.8%) |  |
| I do not know | 10 (2.0%) | 0 (0%) | 0 (0%) |  |
| I do not want to answer | 0 (0%) | 0 (0%) | 9 (1.8%) |  |
| **Beliefs about consequence (TDF-6): BC is unnecessary because a contaminated result often leads to wrong therapeutic approaches (Q3-3-9).** |  |  |  |  |
| Strongly agree | 10 (2.0%) | 7 (2.3%) | 14 (2.8%) | <0.001 |
| Agree | 85 (16.9%) | 23 (7.6%) | 38 (7.6%) |  |
| Uncertain | 128 (25.4%) | 42 (13.8%) | 116 (23.2%) |  |
| Disagree | 229 (45.5%) | 148 (48.7%) | 277 (55.3%) |  |
| Strongly disagree | 41 (8.2%) | 83 (27.3%) | 42 (8.4%) |  |
| I do not know | 9 (1.8%) | 1 (0.3%) | 3 (0.6%) |  |
| I do not want to answer | 1 (0.2%) | 0 (0%) | 11 (2.2%) |  |
| **Environmental context and resources (TDF-11): BC is unnecessary because it is too expensive (Q3-3-10).** |  |  |  |  |
| Strongly agree | 25 (5.0%) | 6 (2.0%) | 12 (2.4%) | <0.001 |
| Agree | 83 (16.5%) | 19 (6.3%) | 24 (4.8%) |  |
| Uncertain | 114 (22.7%) | 37 (12.2%) | 79 (15.8%) |  |
| Disagree | 227 (45.1%) | 133 (43.8%) | 310 (61.9%) |  |
| Strongly disagree | 39 (7.8%) | 103 (33.9%) | 64 (12.8%) |  |
| I do not know | 12 (2.4%) | 5 (1.6%) | 2 (0.4%) |  |
| I do not want to answer | 3 (0.6%) | 1 (0.3%) | 10 (2.0%) |  |
| **Beliefs about consequence (TDF-6): BC is not benefiting the patients (Q3-3-11).** |  |  |  |  |
| Strongly agree | 5 (1.0%) | 5 (1.6%) | 10 (2.0%) | <0.001 |
| Agree | 19 (3.8%) | 17 (5.6%) | 20 (4.0%) |  |
| Uncertain | 88 (17.5%) | 13 (4.3%) | 46 (9.2%) |  |
| Disagree | 290 (57.7%) | 139 (45.7%) | 302 (60.3%) |  |
| Strongly disagree | 92 (18.3%) | 130 (42.8%) | 121 (24.2%) |  |
| I do not know | 8 (1.6%) | 0 (0%) | 0 (0%) |  |
| I do not want to answer | 1 (0.2%) | 0 (0%) | 2 (0.4%) |  |
| **Beliefs about consequence (TDF-6): It is not too late to collect BC later, particularly if patients do not improve after receiving empirical antibiotic treatment (Q3-3-12).** |  |  |  |  |
| Strongly agree | 23 (4.6%) | 48 (15.8%) | 15 (3.0%) | <0.001 |
| Agree | 116 (23.1%) | 114 (37.5%) | 107 (21.4%) |  |
| Uncertain | 95 (18.9%) | 32 (10.5%) | 89 (17.8%) |  |
| Disagree | 208 (41.4%) | 65 (21.4%) | 226 (45.1%) |  |
| Strongly disagree | 49 (9.7%) | 45 (14.8%) | 61 (12.2%) |  |
| I do not know | 11 (2.2%) | 0 (0%) | 3 (0.6%) |  |
| I do not want to answer | 1 (0.2%) | 0 (0%) | 0 (0%) |  |
| **Beliefs about consequence (TDF-6): Quality of laboratory is questionable (Q3-3-13).** |  |  |  |  |
| Strongly agree | 15 (3.0%) | 11 (3.6%) | 9 (1.8%) | <0.001 |
| Agree | 77 (15.3%) | 27 (8.9%) | 55 (11.0%) |  |
| Uncertain | 147 (29.2%) | 81 (26.6%) | 148 (29.5%) |  |
| Disagree | 196 (39.0%) | 114 (37.5%) | 239 (47.7%) |  |
| Strongly disagree | 48 (9.5%) | 62 (20.4%) | 40 (8.0%) |  |
| I do not know | 18 (3.6%) | 8 (2.6%) | 5 (1.0%) |  |
| I do not want to answer | 2 (0.4%) | 1 (0.3%) | 5 (1.0%) |  |
| **Beliefs about consequence (TDF-6):** **Levels of local antibiotic resistance are low (Q3-3-14).** |  |  |  |  |
| Strongly agree | 5 (1.0%) | 4 (1.3%) | 8 (1.6%) | <0.001 |
| Agree | 45 (8.9%) | 22 (7.2%) | 42 (8.4%) |  |
| Uncertain | 120 (23.9%) | 63 (20.7%) | 111 (22.2%) |  |
| Disagree | 225 (44.7%) | 130 (42.8%) | 268 (53.5%) |  |
| Strongly disagree | 87 (17.3%) | 77 (25.3%) | 68 (13.6%) |  |
| I do not know | 21 (4.2%) | 7 (2.3%) | 3 (0.6%) |  |
| I do not want to answer | 0 (0%) | 1 (0.3%) | 1 (0.2%) |  |
| **Goals (TDF-9):** **How often do you obtain BC prior to administration of empirical antibiotics in patients presenting with sepsis (Q3-5)?** |  |  |  |  |
| All the time (>95-100% of the time) | 95 (18.9%) | 158 (52.0%) | 150 (29.9%) | <0.001 |
| Often (75-95% of the time) | 156 (31.0%) | 116 (38.2%) | 230 (45.9%) |  |
| Moderately (25-74% of the time) | 85 (16.9%) | 21 (6.9%) | 64 (12.8%) |  |
| Occasionally (5-24% of the time) | 45 (8.9%) | 5 (1.6%) | 12 (2.4%) |  |
| Rarely (ranging from never <5% of the time) | 82 (16.3%) | 0 (0%) | 19 (3.8%) |  |
| I do not know | 34 (6.8%) | 4 (1.3%) | 11 (2.2%) |  |
| I do not want to answer | 6 (1.2%) | 0 (0%) | 15 (3.0%) |  |
| **Goals (TDF-9):** **How often do you obtain BC prior to administration of empirical antibiotics in patients presenting with septic shock (Q3-6)?** |  |  |  |  |
| All the time (>95-100% of the time) | 90 (17.9%) | 234 (77.0%) | 218 (43.5%) | <0.001 |
| Often (75-95% of the time) | 160 (31.8%) | 59 (19.4%) | 175 (34.9%) |  |
| Moderately (25-74% of the time) | 76 (15.1%) | 6 (2.0%) | 48 (9.6%) |  |
| Occasionally (5-24% of the time) | 48 (9.5%) | 0 (0%) | 18 (3.6%) |  |
| Rarely (ranging from never <5% of the time) | 84 (16.7%) | 0 (0%) | 20 (4.0%) |  |
| I do not know | 40 (8.0%) | 3 (1.0%) | 9 (1.8%) |  |
| I do not want to answer | 5 (1.0%) | 2 (0.7%) | 13 (2.6%) |  |
| **Memory, attention and decision processes (TDF-10): Would you still order BC if patients are already on antibiotics (Q3-7-1)?** |  |  |  |  |
| Definitely not order | 11 (2.2%) | 14 (4.6%) | 6 (1.2%) | <0.001 |
| Likely not order | 19 (3.8%) | 53 (17.4%) | 28 (5.6%) |  |
| Maybe not order | 295 (58.6%) | 38 (12.5%) | 85 (17.0%) |  |
| Likely to still order | 143 (28.4%) | 116 (38.2%) | 308 (61.5%) |  |
| Very likely to still order | 18 (3.6%) | 81 (26.6%) | 72 (14.4%) |  |
| I do not know | 16 (3.2%) | 2 (0.7%) | 1 (0.2%) |  |
| I do not want to answer | 1 (0.2%) | 0 (0%) | 1 (0.2%) |  |
| **Memory, attention and decision processes (TDF-10): Would you still order BC if patients have anemia (Q3-7-2)?** |  |  |  |  |
| Definitely not order | 16 (3.2%) | 84 (27.6%) | 24 (4.8%) | <0.001 |
| Likely not order | 59 (11.7%) | 64 (21.1%) | 33 (6.6%) |  |
| Maybe not order | 255 (50.7%) | 52 (17.1%) | 58 (11.6%) |  |
| Likely to still order | 124 (24.7%) | 52 (17.1%) | 257 (51.3%) |  |
| Very likely to still order | 20 (4.0%) | 45 (14.8%) | 115 (23.0%) |  |
| I do not know | 28 (5.6%) | 5 (1.6%) | 2 (0.4%) |  |
| I do not want to answer | 1 (0.2%) | 2 (0.7%) | 12 (2.4%) |  |
| **Memory, attention and decision processes (TDF-10): Would you still order BC if blood should be used for other laboratory tests (Q3-7-3)?** |  |  |  |  |
| Definitely not order | 7 (1.4%) | 57 (18.8%) | 59 (11.8%) | <0.001 |
| Likely not order | 43 (8.5%) | 57 (18.8%) | 64 (12.8%) |  |
| Maybe not order | 228 (45.3%) | 75 (24.7%) | 117 (23.4%) |  |
| Likely to still order | 158 (31.4%) | 63 (20.7%) | 172 (34.3%) |  |
| Very likely to still order | 20 (4.0%) | 40 (13.2%) | 60 (12.0%) |  |
| I do not know | 41 (8.2%) | 12 (3.9%) | 21 (4.2%) |  |
| I do not want to answer | 6 (1.2%) | 0 (0%) | 8 (1.6%) |  |
| **Memory, attention and decision processes (TDF-10): Would you still order BC if there are no local guidelines/guidelines for BC sampling (Q3-7-4)?** |  |  |  |  |
| Definitely not order | 11 (2.2%) | 42 (13.8%) | 42 (8.4%) | <0.001 |
| Likely not order | 41 (8.2%) | 43 (14.1%) | 66 (13.2%) |  |
| Maybe not order | 241 (47.9%) | 95 (31.3%) | 136 (27.1%) |  |
| Likely to still order | 152 (30.2%) | 66 (21.7%) | 174 (34.7%) |  |
| Very likely to still order | 19 (3.8%) | 33 (10.9%) | 41 (8.2%) |  |
| I do not know | 32 (6.4%) | 24 (7.9%) | 35 (7.0%) |  |
| I do not want to answer | 7 (1.4%) | 1 (0.3%) | 7 (1.4%) |  |
| **Memory, attention and decision processes (TDF-10): Would you still order BC if patients do not meet certain conditions for a BC following the local guidelines (Q3-7-5)?** |  |  |  |  |
| Definitely not order | 28 (5.6%) | 39 (12.8%) | 54 (10.8%) | <0.001 |
| Likely not order | 131 (26.0%) | 80 (26.3%) | 93 (18.6%) |  |
| Maybe not order | 250 (49.7%) | 93 (30.6%) | 177 (35.3%) |  |
| Likely to still order | 58 (11.5%) | 54 (17.8%) | 121 (24.2%) |  |
| Very likely to still order | 11 (2.2%) | 22 (7.2%) | 44 (8.8%) |  |
| I do not know | 23 (4.6%) | 15 (4.9%) | 8 (1.6%) |  |
| I do not want to answer | 2 (0.4%) | 1 (0.3%) | 4 (0.8%) |  |
| **Memory, attention and decision processes (TDF-10): Would you still order BC if patients do not have a health scheme or insurance that covers the cost of BC (Q3-7-6)?** |  |  |  |  |
| Definitely not order | 39 (7.8%) | 7 (2.3%) | 21 (4.2%) | <0.001 |
| Likely not order | 56 (11.1%) | 33 (10.9%) | 43 (8.6%) |  |
| Maybe not order | 306 (60.8%) | 95 (31.3%) | 101 (20.2%) |  |
| Likely to still order | 68 (13.5%) | 87 (28.6%) | 265 (52.9%) |  |
| Very likely to still order | 6 (1.2%) | 63 (20.7%) | 61 (12.2%) |  |
| I do not know | 23 (4.6%) | 14 (4.6%) | 5 (1.0%) |  |
| I do not want to answer | 5 (1.0%) | 5 (1.6%) | 5 (1.0%) |  |
| **Memory, attention and decision processes (TDF-10): Would you still order BC if microbiology laboratory in your hospital is not available (Q3-7-7)?** |  |  |  |  |
| Definitely not order | 53 (10.5%) | 21 (6.9%) | 97 (19.4%) | <0.001 |
| Likely not order | 114 (22.7%) | 53 (17.4%) | 101 (20.2%) |  |
| Maybe not order | 229 (45.5%) | 77 (25.3%) | 120 (24.0%) |  |
| Likely to still order | 74 (14.7%) | 79 (26.0%) | 109 (21.8%) |  |
| Very likely to still order | 10 (2.0%) | 54 (17.8%) | 36 (7.2%) |  |
| I do not know | 19 (3.8%) | 12 (3.9%) | 30 (6.0%) |  |
| I do not want to answer | 4 (0.8%) | 8 (2.6%) | 8 (1.6%) |  |
| **Environmental context and resources (TDF-11): How often could you not order BC because consumables are not available (Q4-1)?** |  |  |  |  |
| All the time (>95-100% of the time) | 24 (4.8%) | 12 (3.9%) | 19 (3.8%) | <0.001 |
| Often (75-95% of the time) | 61 (12.1%) | 15 (4.9%) | 19 (3.8%) |  |
| Moderately (25-74% of the time) | 52 (10.3%) | 11 (3.6%) | 56 (11.2%) |  |
| Occasionally (5-24% of the time) | 86 (17.1%) | 15 (4.9%) | 51 (10.2%) |  |
| Rarely (ranging from never <5% of the time) | 219 (43.5%) | 232 (76.3%) | 309 (61.7%) |  |
| I do not know | 53 (10.5%) | 18 (5.9%) | 25 (5.0%) |  |
| I do not want to answer | 8 (1.6%) | 1 (0.3%) | 22 (4.4%) |  |
| **Environmental context and resources (TDF-11): How often could you not order BC because the microbiology laboratory is not available or not functioning (Q4-2)?** |  |  |  |  |
| All the time (>95-100% of the time) | 34 (6.8%) | 9 (3.0%) | 15 (3.0%) | <0.001 |
| Often (75-95% of the time) | 58 (11.5%) | 13 (4.3%) | 28 (5.6%) |  |
| Moderately (25-74% of the time) | 48 (9.5%) | 9 (3.0%) | 37 (7.4%) |  |
| Occasionally (5-24% of the time) | 78 (15.5%) | 14 (4.6%) | 27 (5.4%) |  |
| Rarely (ranging from never <5% of the time) | 224 (44.5%) | 238 (78.3%) | 342 (68.3%) |  |
| I do not know | 56 (11.1%) | 21 (6.9%) | 28 (5.6%) |  |
| I do not want to answer | 5 (1.0%) | 0 (0%) | 24 (4.8%) |  |
| **Environmental context and resources (TDF-11): How often do patients have to pay for BC using their own money (i.e. out of pocket) (Q4-3)?** |  |  |  |  |
| All the time (>95-100% of the time) | 26 (5.2%) | 11 (3.6%) | 6 (1.2%) | <0.001 |
| Often (75-95% of the time) | 52 (10.3%) | 17 (5.6%) | 28 (5.6%) |  |
| Moderately (25-74% of the time) | 50 (9.9%) | 19 (6.3%) | 67 (13.4%) |  |
| Occasionally (5-24% of the time) | 69 (13.7%) | 48 (15.8%) | 134 (26.7%) |  |
| Rarely (ranging from never <5% of the time) | 138 (27.4%) | 135 (44.4%) | 173 (34.5%) |  |
| I do not know | 163 (32.4%) | 73 (24.0%) | 72 (14.4%) |  |
| I do not want to answer | 5 (1.0%) | 1 (0.3%) | 21 (4.2%) |  |
| **Environmental context and resources (TDF-11): Would you say that the benefits of BC outweigh the cost (Q4-4)?** |  |  |  |  |
| Very likely | 101 (20.1%) | 135 (44.4%) | 184 (36.7%) | <0.001 |
| Likely | 210 (41.7%) | 97 (31.9%) | 223 (44.5%) |  |
| Uncertain | 93 (18.5%) | 37 (12.2%) | 34 (6.8%) |  |
| Unlikely | 45 (8.9%) | 10 (3.3%) | 16 (3.2%) |  |
| Very unlikely | 3 (0.6%) | 13 (4.3%) | 17 (3.4%) |  |
| I do not know | 49 (9.7%) | 12 (3.9%) | 17 (3.4%) |  |
| I do not want to answer | 2 (0.4%) | 0 (0%) | 10 (2.0%) |  |
| **Reinforcement (TDF-7): Are there any positive consequences if you order a BC when recommended (Q5-1)?** |  |  |  |  |
| No | 283 (56.3%) | 187 (61.5%) | 206 (41.1%) | <0.001 |
| Yes, social | 31 (6.2%) | 37 (12.2%) | 59 (11.8%) |  |
| Yes, material | 4 (0.8%) | 2 (0.7%) | 8 (1.6%) |  |
| Yes, both social and material | 33 (6.6%) | 18 (5.9%) | 103 (20.6%) |  |
| I do not know | 143 (28.4%) | 58 (19.1%) | 75 (15.0%) |  |
| I do not want to answer | 8 (1.6%) | 1 (0.3%) | 45 (9.0%) |  |
| Other | 1 (0.2%) | 1 (0.3%) | 5 (1.0%) |  |
| **Reinforcement (TDF-7): Are there any negative consequences if you do not order a BC when recommended (Q5-2)?** |  |  |  |  |
| No | 248 (49.3%) | 101 (33.2%) | 134 (26.7%) | <0.001 |
| Yes, social | 65 (12.9%) | 115 (37.8%) | 100 (20.0%) |  |
| Yes, material | 8 (1.6%) | 4 (1.3%) | 13 (2.6%) |  |
| Yes, both social and material | 27 (5.4%) | 22 (7.2%) | 111 (22.2%) |  |
| I do not know | 142 (28.2%) | 60 (19.7%) | 83 (16.6%) |  |
| I do not want to answer | 12 (2.4%) | 2 (0.7%) | 55 (11.0%) |  |
| Other | 1 (0.2%) | 0 (0%) | 5 (1.0%) |  |
| **Reinforcement (TDF-7): Are there any negative consequences if you order a BC when recommended (Q5-3)?** |  |  |  |  |
| No | 251 (49.9%) | 162 (53.3%) | 210 (41.9%) | <0.001 |
| Yes, social | 47 (9.3%) | 43 (14.1%) | 31 (6.2%) |  |
| Yes, material | 10 (2.0%) | 3 (1.0%) | 31 (6.2%) |  |
| Yes, both social and material | 30 (6.0%) | 14 (4.6%) | 91 (18.2%) |  |
| I do not know | 150 (29.8%) | 78 (25.7%) | 83 (16.6%) |  |
| I do not want to answer | 14 (2.8%) | 4 (1.3%) | 53 (10.6%) |  |
| Other | 1 (0.2%) | 0 (0%) | 2 (0.4%) |  |
| **Behaviour regulation (TDF-14): Any training, lectures, classes or meetings that provide you knowledge about local/national/international guidelines for BC sampling (Q5-5)?** |  |  |  |  |
| No | 153 (30.4%) | 64 (21.1%) | 52 (10.4%) | <0.001 |
| Yes, infrequent (less than once a year) | 90 (17.9%) | 87 (28.6%) | 111 (22.2%) |  |
| Yes, occasionally (at least once a year) | 109 (21.7%) | 84 (27.6%) | 196 (39.1%) |  |
| Yes, regularly | 53 (10.5%) | 22 (7.2%) | 61 (12.2%) |  |
| I do not know | 91 (18.1%) | 46 (15.1%) | 74 (14.8%) |  |
| I do not want to answer | 5 (1.0%) | 1 (0.3%) | 6 (1.2%) |  |
| Other | 2 (0.4%) | 0 (0%) | 1 (0.2%) |  |
| **Behaviour regulation (TDF-14): any procedures that support you or doctors to order or regulate ordering of BC per local/national/international guidelines (Q5-6)?** |  |  |  |  |
| No | 129 (25.7%) | 71 (23.4%) | 76 (15.2%) | <0.001 |
| Poster | 57 (11.3%) | 40 (13.2%) | 66 (13.2%) | 0.62 |
| Standard order form | 120 (23.9%) | 90 (29.6%) | 107 (21.4%) | 0.03 |
| Computer system to remind ordering BC | 25 (5.0%) | 14 (4.6%) | 74 (14.8%) | <0.001 |
| case review (e.g. grand round; morning ward round, clinical meetings, and BC is often mentioned) | 76 (15.1%) | 86 (28.3%) | 164 (32.7%) | <0.001 |
| Stewardship programme and reviewing BC is included in the programme | 61 (12.1%) | 25 (8.2%) | 121 (24.2%) | <0.001 |
| Local hospital guideline (e.g. standard operating procedure [SOP]) | 113 (22.5%) | 77 (25.3%) | 162 (32.3%) | 0.002 |
| I do not know | 107 (21.3%) | 49 (16.1%) | 66 (13.2%) | 0.003 |
| I do not want to answer | 9 (1.8%) | 2 (0.7%) | 15 (3.0%) | 0.07 |
| **Social influence (TDF-12): To what extent do you order BC because you are following local norms (Q6-1)?** |  |  |  |  |
| All the time (>95-100% of the time) | 50 (9.9%) | 67 (22.0%) | 64 (12.8%) | <0.001 |
| Often (75-95% of the time) | 130 (25.8%) | 166 (54.6%) | 174 (34.7%) |  |
| Moderately (25-74% of the time) | 84 (16.7%) | 41 (13.5%) | 144 (28.7%) |  |
| Occasionally (5-24% of the time) | 67 (13.3%) | 15 (4.9%) | 40 (8.0%) |  |
| Rarely (ranging from never <5% of the time) | 80 (15.9%) | 8 (2.6%) | 40 (8.0%) |  |
| I do not know | 87 (17.3%) | 7 (2.3%) | 25 (5.0%) |  |
| I do not want to answer | 5 (1.0%) | 0 (0%) | 14 (2.8%) |  |
| **Social influence (TDF-12): Influence from nurses (Q6-2-1)? Positive influence could mean facilitate, support or encourage BC sampling. Negative influence could mean hinder or discourage BC sampling.** |  |  |  |  |
| Very positive influence | 46 (9.1%) | 29 (9.5%) | 60 (12.0%) | <0.001 |
| Positive influence | 230 (45.7%) | 103 (33.9%) | 154 (30.7%) |  |
| Neither positive nor negative influence | 162 (32.2%) | 122 (40.1%) | 228 (45.5%) |  |
| Negative influence | 15 (3.0%) | 26 (8.6%) | 25 (5.0%) |  |
| Very negative influence | 1 (0.2%) | 1 (0.3%) | 0 (0%) |  |
| I do not know | 45 (8.9%) | 19 (6.3%) | 30 (6.0%) |  |
| I do not want to answer | 4 (0.8%) | 4 (1.3%) | 4 (0.8%) |  |
| **Social influence (TDF-12): Influence from final-year medical students (Q6-2-2)?** |  |  |  |  |
| Very positive influence | 29 (5.8%) | 22 (7.2%) | 30 (6.0%) | 0.004 |
| Positive influence | 155 (30.8%) | 87 (28.6%) | 104 (20.8%) |  |
| Neither positive nor negative influence | 249 (49.5%) | 157 (51.6%) | 315 (62.9%) |  |
| Negative influence | 4 (0.8%) | 3 (1.0%) | 6 (1.2%) |  |
| Very negative influence | 1 (0.2%) | 1 (0.3%) | 0 (0%) |  |
| I do not know | 60 (11.9%) | 27 (8.9%) | 42 (8.4%) |  |
| I do not want to answer | 5 (1.0%) | 7 (2.3%) | 4 (0.8%) |  |
| **Social influence (TDF-12): Influence from Interns (Q6-2-3)?** |  |  |  |  |
| Very positive influence | 31 (6.2%) | 41 (13.5%) | 33 (6.6%) | <0.001 |
| Positive influence | 182 (36.2%) | 134 (44.1%) | 170 (33.9%) |  |
| Neither positive nor negative influence | 205 (40.8%) | 96 (31.6%) | 251 (50.1%) |  |
| Negative influence | 5 (1.0%) | 4 (1.3%) | 3 (0.6%) |  |
| Very negative influence | 1 (0.2%) | 0 (0%) | 1 (0.2%) |  |
| I do not know | 70 (13.9%) | 24 (7.9%) | 38 (7.6%) |  |
| I do not want to answer | 9 (1.8%) | 5 (1.6%) | 5 (1.0%) |  |
| **Social influence (TDF-12): Influence from residents (Q6-2-4)?** |  |  |  |  |
| Very positive influence | 64 (12.7%) | 73 (24.0%) | 79 (15.8%) | <0.001 |
| Positive influence | 270 (53.7%) | 138 (45.4%) | 219 (43.7%) |  |
| Neither positive nor negative influence | 109 (21.7%) | 63 (20.7%) | 161 (32.1%) |  |
| Negative influence | 2 (0.4%) | 3 (1.0%) | 1 (0.2%) |  |
| Very negative influence | 0 (0%) | 0 (0%) | 1 (0.2%) |  |
| I do not know | 51 (10.1%) | 23 (7.6%) | 37 (7.4%) |  |
| I do not want to answer | 7 (1.4%) | 4 (1.3%) | 3 (0.6%) |  |
| **Social influence (TDF-12): Influence from doctors (Q6-2-5)?** |  |  |  |  |
| Very positive influence | 82 (16.3%) | 62 (20.4%) | 67 (13.4%) | <0.001 |
| Positive influence | 293 (58.3%) | 125 (41.1%) | 216 (43.1%) |  |
| Neither positive nor negative influence | 90 (17.9%) | 85 (28.0%) | 188 (37.5%) |  |
| Negative influence | 6 (1.2%) | 3 (1.0%) | 3 (0.6%) |  |
| Very negative influence | 0 (0%) | 3 (1.0%) | 1 (0.2%) |  |
| I do not know | 29 (5.8%) | 23 (7.6%) | 15 (3.0%) |  |
| I do not want to answer | 3 (0.6%) | 3 (1.0%) | 11 (2.2%) |  |
| **Social influence (TDF-12): Influence from consultants (Q6-2-6)?** |  |  |  |  |
| Very positive influence | 172 (34.2%) | 117 (38.5%) | 109 (21.8%) | <0.001 |
| Positive influence | 255 (50.7%) | 125 (41.1%) | 261 (52.1%) |  |
| Neither positive nor negative influence | 38 (7.6%) | 41 (13.5%) | 113 (22.6%) |  |
| Negative influence | 5 (1.0%) | 4 (1.3%) | 4 (0.8%) |  |
| Very negative influence | 1 (0.2%) | 2 (0.7%) | 0 (0%) |  |
| I do not know | 26 (5.2%) | 11 (3.6%) | 13 (2.6%) |  |
| I do not want to answer | 6 (1.2%) | 4 (1.3%) | 1 (0.2%) |  |
| **Social influence (TDF-12): Influence from head of department (Q6-2-7)?** |  |  |  |  |
| Very positive influence | 81 (16.1%) | 51 (16.8%) | 135 (26.9%) | <0.001 |
| Positive influence | 254 (50.5%) | 89 (29.3%) | 252 (50.3%) |  |
| Neither positive nor negative influence | 104 (20.7%) | 119 (39.1%) | 95 (19.0%) |  |
| Negative influence | 10 (2.0%) | 6 (2.0%) | 6 (1.2%) |  |
| Very negative influence | 0 (0%) | 1 (0.3%) | 0 (0%) |  |
| I do not know | 48 (9.5%) | 34 (11.2%) | 11 (2.2%) |  |
| I do not want to answer | 6 (1.2%) | 4 (1.3%) | 2 (0.4%) |  |
| **Social influence (TDF-12): Influence from executive or administrative level of the hospital (Q6-2-8)?** |  |  |  |  |
| Very positive influence | 55 (10.9%) | 35 (11.5%) | 101 (20.2%) | <0.001 |
| Positive influence | 188 (37.4%) | 67 (22.0%) | 216 (43.1%) |  |
| Neither positive nor negative influence | 169 (33.6%) | 145 (47.7%) | 154 (30.7%) |  |
| Negative influence | 21 (4.2%) | 8 (2.6%) | 7 (1.4%) |  |
| Very negative influence | 8 (1.6%) | 2 (0.7%) | 1 (0.2%) |  |
| I do not know | 57 (11.3%) | 42 (13.8%) | 19 (3.8%) |  |
| I do not want to answer | 5 (1.0%) | 5 (1.6%) | 3 (0.6%) |  |
| **Social influence (TDF-12): Influence from patients (Q6-2-9)?** |  |  |  |  |
| Very positive influence | 43 (8.5%) | 44 (14.5%) | 57 (11.4%) | <0.001 |
| Positive influence | 197 (39.2%) | 74 (24.3%) | 148 (29.5%) |  |
| Neither positive nor negative influence | 197 (39.2%) | 141 (46.4%) | 250 (49.9%) |  |
| Negative influence | 18 (3.6%) | 14 (4.6%) | 21 (4.2%) |  |
| Very negative influence | 1 (0.2%) | 1 (0.3%) | 1 (0.2%) |  |
| I do not know | 44 (8.7%) | 26 (8.6%) | 20 (4.0%) |  |
| I do not want to answer | 3 (0.6%) | 4 (1.3%) | 4 (0.8%) |  |
| **Social influence (TDF-12): Influence from family of patients (Q6-2-10)?** |  |  |  |  |
| Very positive influence | 32 (6.4%) | 21 (6.9%) | 34 (6.8%) | <0.001 |
| Positive influence | 171 (34.0%) | 40 (13.2%) | 119 (23.8%) |  |
| Neither positive nor negative influence | 221 (43.9%) | 186 (61.2%) | 282 (56.3%) |  |
| Negative influence | 23 (4.6%) | 20 (6.6%) | 39 (7.8%) |  |
| Very negative influence | 3 (0.6%) | 2 (0.7%) | 2 (0.4%) |  |
| I do not know | 50 (9.9%) | 30 (9.9%) | 19 (3.8%) |  |
| I do not want to answer | 3 (0.6%) | 5 (1.6%) | 6 (1.2%) |  |
| **Emotions (TDF-13): Any emotional factors (Q6-4)?** |  |  |  |  |
| Yes | 51 (10.1%) | 10 (3.3%) | 32 (6.4%) | 0.001 |
| **Gender (Q7-2)** |  |  |  |  |
| Female | 263 (52.3%) | 195 (64.1%) | 222 (44.3%) | <0.001 |
| Male | 236 (46.9%) | 106 (34.9%) | 263 (52.5%) |  |
| Other | 1 (0.2%) | 0 (0%) | 0 (0%) |  |
| I do not want to answer | 3 (0.6%) | 3 (1.0%) | 16 (3.2%) |  |
| **Hospital bed size (Q7-3)** |  |  |  |  |
| <200 | 99 (19.7%) | 35 (11.5%) | 24 (4.8%) | <0.001 |
| 201-400 | 107 (21.3%) | 46 (15.1%) | 29 (5.8%) |  |
| 401-600 | 72 (14.3%) | 39 (12.8%) | 62 (12.4%) |  |
| 601-1,000 | 66 (13.1%) | 45 (14.8%) | 144 (28.7%) |  |
| 1,001-2,000 | 39 (7.8%) | 82 (27.0%) | 125 (25.0%) |  |
| > 2,000 | 27 (5.4%) | 30 (9.9%) | 74 (14.8%) |  |
| I do not know | 89 (17.7%) | 27 (8.9%) | 35 (7.0%) |  |
| I do not want to answer | 4 (0.8%) | 0 (0%) | 8 (1.6%) |  |
| **Department (Q7-4)** |  |  |  |  |
| Internal medicine | 149 (29.6%) | 155 (51.0%) | 146 (29.1%) | <0.001 |
| Pediatrics | 65 (12.9%) | 43 (14.1%) | 45 (9.0%) | 0.05 |
| Infection disease division/department | 12 (2.4%) | 5 (1.6%) | 56 (11.2%) | <0.001 |
| Surgery | 21 (4.2%) | 45 (14.8%) | 81 (16.2%) | <0.001 |
| Orthopaedics | 6 (1.2%) | 18 (5.9%) | 14 (2.8%) | 0.001 |
| Obstetrics / Gynaecology | 20 (4.0%) | 29 (9.5%) | 7 (1.4%) | <0.001 |
| Emergency department | 112 (22.3%) | 34 (11.2%) | 29 (5.8%) | <0.001 |
| Intensive care unit | 45 (8.9%) | 13 (4.3%) | 51 (10.2%) | 0.01 |
| I do not want to answer | 24 (4.8%) | 25 (8.2%) | 52 (10.4%) | 0.004 |
| Other | 137 (27.2%) | 29 (9.5%) | 58 (11.6%) | <0.001 |

Gray color represents questions that were asked to subsets of participants. ^1^ Included primary health care, clinic, retired and answers as role of doctors (including residents, interns and medical students).

**Appendix S12. Associations between barriers or enablers and the responses that they would definitely take BC in the case scenario in the Theory Domain Framework (TDF) survey**

| **Barriers or enablers** | **Indonesia**^1^  **(n=503)** | **Thailand**^1^  **(n=304)** | **Viet Nam**^1^  **(n=501)** | **Odds ratio**^2^ | **P value** |
| --- | --- | --- | --- | --- | --- |
| **TDF Domain: Knowledge** |  |  |  |  |  |
| **Awareness of local guidelines** |  |  |  |  |  |
| Yes | 42.7% (102/239) | 91.1% (154/169) | 59.5% (206/346) | 2.55 (1.93-3.38) | <0.001 |
| No^1^ | 21.1% (53/251) | 89.3% (117/131) | 29.4% (42/143) | 1.0 |  |
| **Awareness of international guidelines** |  |  |  |  |  |
| Yes | 38.9% (138/226) | 90.8% (128/141) | 65.9% (147/223) | 1.97 (1.50-2.57) | <0.001 |
| No | 25.4% (67/264) | 89.9% (143/159) | 38.0% (101/266) | 1.0 |  |
| **Any training, lectures, classes or meetings that provide knowledge about guidelines for BC sampling** |  |  |  |  |  |
| Available | 36.2% (92/254) | 92.2% (178/193) | 53.5% (197/368) | 1.68 (1.18-2.38) | 0.004 |
| Not available | 21.7% (33/152) | 82.8% (53/64) | 46.2% (24/52) | 1.0 |  |
| **TDF Domain: Goals** |  |  |  |  |  |
| How often do you obtain BC prior to receiving empirical antibiotic in patients presenting with sepsis? |  |  |  |  |  |
| All the time / Often (>75-100% of the time) | 45.4% (113/249) | 91.6% (251/274) | 58.6% (222/379) | 4.25 (3.04-5.94) | <0.001 |
| Moderately / Occasionally / Rarely (0-74% of the time) | 15.6% (33/212) | 73.1% (19/26) | 22.1% (21/95) | 1.0 |  |
| How often do you obtain BC prior to receiving empirical antibiotic in patients presenting with septic shock? |  |  |  |  |  |
| All the time / Often (>75-100% of the time) | 44.8% (111/248) | 90.1% (264/293) | 56.4% (221/392) | 3.71 (2.61-5.27) | <0.001 |
| Moderately / Occasionally / Rarely (0-74% of the time) | 15.4% (32/208) | 83.3% (5/6) | 25.6% (22/86) | 1.0 |  |
| **TDF Domain: Intention** |  |  |  |  |  |
| **Intention to follow local guidelines**^3^ |  |  |  |  |  |
| All the time / Often (>75-100% of the cases) | 51.7% (89/172) | 90.5% (142/157) | 64.7% (183/283) | 2.92 (1.88-4.53) | <0.001 |
| Moderately / Occasionally / Rarely (0-74% of the cases) | 18.6% (11/59) | 100% (12/12) | 37.7% (23/61) | 1.0 |  |
| **TDF Domain: Social professional role and identity** |  |  |  |  |  |
| **Current job** |  |  |  |  |  |
| Medical doctor – an executive level | 15.4% (2/13) | 60.0% (2/3) | 35.3% (6/17) | 0.20 (0.09-0.47) | <0.001 |
| Medical doctor – a consultant level | 34.4% (25/73) | 90.7% (68/75) | 49.2% (97/197) | 0.48 (0.33-0.69) |  |
| Medical doctor – a general physician level | 10.5% (13/124) | 81.6% (31/38) | 46.0% (51/111) | 0.27 (0.18-0.40) |  |
| Medical doctor – a resident/registra/fellow level | 48.8% (82/168) | 93.7% (59/63) | 68.3% (69/101) | 1.0 |  |
| Intern – recent medical school graduate | 12.1% (4/33) | 88.6% (31/35) | 35.7% (5/14) | 0.26 (0.14-0.49) |  |
| Final-year medical student | 34.4% (31/90) | 92.1% (81/88) | 40.7% (24/59) | 0.50 (0.33-0.76) |  |
| **Perception about their role to order or initiate an order for BC** |  |  |  |  |  |
| Very appropriate / Appropriate | 45.5% (120/264) | 91.2% (250/274) | 61.2% (195/317) | 3.36 (2.50-4.51) | <0.001 |
| Uncertain / Inappropriate / Very inappropriate | 16% (36/225) | 78.6% (22/28) | 33.3% (55/165) | 1.0 |  |
| **Perception about their role to draw blood for BC**^3^ |  |  |  |  |  |
| Very appropriate / Appropriate | 38.0% (27/71) | 87.8% (65/74) | 52.4% (54/103) | 1.94 (1.04-3.64) | 0.04 |
| Uncertain / Inappropriate / Very inappropriate | 28.6% (4/14) | 94.8% (55/58) | 25.6% (10/39) | 1.0 |  |
| **TDF Domain: Social influences** |  |  |  |  |  |
| To what extent do you order BC in your hospital because you are following local norms? ^5^ |  |  |  |  |  |
| All the time / Often (>75-100% of the time) | 45.3% (81/179) | 90.1% (210/233) | 61.3% (146/238) | 2.20 (1.67-2.90) | <0.001 |
| Moderately / Occasionally / Rarely (0-74% of the time) | 22.2% (51/230) | 90.6% (58/64) | 41.3% (92/223) | 1.0 |  |
| **TDF Domain: Environmental context and resources** |  |  |  |  |  |
| **Do the benefits of BC outweigh the cost?** |  |  |  |  |  |
| Very likely / likely | 35.3% (109/309) | 91.0% (211/232) | 53.1% (216/407) | 1.63 (1.17-2.26) | 0.004 |
| Uncertain / Unlikely / Very unlikely | 22.0% (31/141) | 86.7% (52/60) | 42.3% (29/67) | 1.0 |  |
| **How often are consumables for BC not available?** |  |  |  |  |  |
| All the time / Often (>75-100% of the time) | 31.3% (26/83) | 88.9% (24/27) | 34.2% (13/38) | 0.81 (0.53-1.22) | 0.32 |
| Moderately / Occasionally / Rarely (0-74% of the time) | 31.9% (114/357) | 89.5% (231/258) | 53.5% (222/415) | 1.0 |  |
| **How often are laboratories not available or not functioning?** |  |  |  |  |  |
| All the time / Often (>75-100% of the time) | 28.9% (26/90) | 90.9% (2/22) | 48.8% (21/43) | 0.94 (0.63-1.41) | 0.78 |
| Moderately / Occasionally / Rarely (0-74% of the time) | 32.6% (114/350) | 89.3% (233/261) | 53.3% (216/405) | 1.0 |  |
| **How often do patients have to pay for BC using their own money?** |  |  |  |  |  |
| All the time / Often (>75-100% of the time) | 22.4% (17/76) | 92.7% (26/28) | 47.1% (16/34) | 0.79 (0.51-1.22) | 0.29 |
| Moderately / Occasionally / Rarely (0-74% of the time) | 36.2% (93/257) | 88.1% (178/202) | 55.8% (208/373) | 1.0 |  |
| **Considering whether “patients can afford the cost of BC” as another reason for deciding to do BC sampling** |  |  |  |  |  |
| Yes | 31.1% (33/106) | 92.6% (25/27) | 46.9% (30/64) | 1.12 (0.79-1.61) | 0.53 |
| No | 31.4% (124/395) | 89.5% (248/277) | 51.0% (222/435) | 1.0 |  |
| **TDF Domain: Behavioural regulation** |  |  |  |  |  |
| **Considering whether “patients have a health scheme or insurance that covers the cost of BC” as another reason for deciding to do BC sampling** ^6^ |  |  |  |  |  |
| Yes | 27.7% (31/112) | 92.6% (25/27) | 38.6% (22/57) | 0.82 (0.57-1.18) | 0.29 |
| No | 32.4% (126/389) | 89.5% (248/277) | 52.0% (230/442) | 1.0 |  |
| **Considering whether “Patients are likely to have a final diagnosis that includes the cost of BC in the package of fee for service” as another reason for deciding to do BC sampling** ^6^ |  |  |  |  |  |
| Yes | 33.8% (24/69) | 96.4% (27/28) | 41.8% (23/55) | 1.04 (0.70-1.54) | 0.85 |
| No | 30.8% (133/432) | 89.1% (246/276) | 51.6% (229/444) | 1.0 |  |
| **Procedures that support doctors to order or regulate ordering of BC** |  |  |  |  |  |
| No | 44.7% (34/76) | 88.7% (63/71) | 24.2% (31/128) | 1.0 | 0.006 |
| Poster (and BC is mentioned) | 36.8% 921/57) | 92.5% (37/40) | 51.5% (34/66) | 1.13 (0.76-1.69) |  |
| Standard order form for patients with sepsis (with BC written) | 32.5% (39/120) | 92.2% (83/90) | 46.7% (50/107) | 0.82 (0.59-1.14) |  |
| Computer system to remind ordering BC | 36.0% (9/25) | 92.9% (13/14) | 45% (33/73) | 0.72 (0.48-1.15) |  |
| case reviews (e.g. grand round; with BC often mentioned) | 44.7% (34/76) | 90.7% (78/86) | 57.3% (94/164) | 1.38 (0.94-2.00) |  |
| Stewardship programmes (including BC) | 49.2% (30/61) | 92.0% (23/25) | 58.7% (71/121) | 1.33 (0.87-2.03) |  |
| Local hospital guideline (e.g. standard operating procedure) | 37.2% (42/113) | 94.8% (73/77) | 58.6% (95/162) | 1.45 (1.06-1.99) |  |
| **TDF Domain: Reinforcement** |  |  |  |  |  |
| Positive consequences if doctors order a BC when recommended |  |  |  |  |  |
| Yes | 29.9% (20/67) | 86.0% (49/57) | 42.4% (72/170) | 0.53 (0.37-0.74) | <0.001 |
| No | 32.0% (136/425) | 90.6% (222/245) | 57.4% (160/279) | 1.0 |  |
| Negative consequences if doctors do not order a BC when recommended |  |  |  |  |  |
| Yes | 39.4% (39/99) | 90.1% (127/141) | 50.0% (112/224) | 0.87 (0.63-1.21) | 0.42 |
| No | 30.1% (117/389) | 89.4% (144/161) | 55.6% (120/216) | 1.0 |  |
| Negative consequences if doctors order a BC when recommended |  |  |  |  |  |
| Yes | 29.2% (19/65) | 86.0% (49/57) | 41.4% (67/162) | 0.48 (0.34-0.67) | <0.001 |
| No | 32.3% (136/421) | 90.5% (220/243) | 60.1% (170/283) | 1.0 |  |
| **TDF Domain: Belief about consequences** |  |  |  |  |  |
| BC is helpful in clinical decision |  |  |  |  |  |
| Strongly agree / Agree | 31.5% (152/482) | 89.9% (267/297) | 54.1% (237/438) | 2.96 (1.71-5.12) | <0.001 |
| Uncertain / Disagree / Strongly disagree | 23.5% (4/17) | 85.7% (6/7) | 23.7% (14/59) | 1.0 |  |
| BC is helpful to rule in an infection |  |  |  |  |  |
| Strongly agree / Agree | 31.9% (149/467) | 90.1% (254/282) | 52.4% (220/420) | 1.58 (1.04-2.39) | 0.03 |
| Uncertain / Disagree / Strongly disagree | 21.9% (7/32) | 100% (18/18) | 40.3% (31/77) | 1.0 |  |
| BC is helpful to rule out an infection |  |  |  |  |  |
| Strongly agree / Agree | 31.2% (123/394) | 88.2% (149/169) | 47.7% (105/220) | 0.91 (0.69-1.19) | 0.49 |
| Uncertain / Disagree / Strongly disagree | 31.4% (33/105) | 91.7% (122/133) | 52.9% (146/276) | 1.0 |  |
| BC is helpful to detecting AMR bacterial infections |  |  |  |  |  |
| Strongly agree / Agree | 31.3% (152/485) | 89.2% (256/287) | 51.2% (217/424) | 1.26 (0.80-1.98) | 0.32 |
| Uncertain / Disagree / Strongly disagree | 28.6% (4/14) | 100% (16/16) | 45.2% (33/73) | 1.0 |  |
| BC is helpful in adjusting antibiotics |  |  |  |  |  |
| Strongly agree / Agree | 31.0% (152/490) | 89.7% (269/300) | 52.2% (225/431) | 1.50 (0.90-2.50) | 0.12 |
| Uncertain / Disagree / Strongly disagree | 44.4% (4/9) | 100% (4/4) | 39.1% (25/64) | 1.0 |  |
| BC can reduce overuse of antibiotics |  |  |  |  |  |
| Strongly agree / Agree | 30.7% (141/460) | 89.0% (243/273) | 52.2% (211/404) | 1.08 (0.74-1.58) | 0.68 |
| Uncertain / Disagree / Strongly disagree | 38.5% (15/39) | 97% (30/31) | 42.0% (40/93) | 1.0 |  |
| BC can reduce length of hospital stay |  |  |  |  |  |
| Strongly agree / Agree | 31.5% (120/381) | 91.5% (204/223) | 55.3% (183/331) | 1.53 (1.14-2.04) | 0.004 |
| Uncertain / Disagree / Strongly disagree | 29.6% (34/115) | 86.1% (68/79) | 41.0% (68/166) | 1.0 |  |
| BC can reduce patient mortality |  |  |  |  |  |
| Strongly agree / Agree | 32.8% (133/405) | 89.0% (227/255) | 55.0% (200/364) | 1.61 (1.18-2.20) | 0.003 |
| Uncertain / Disagree / Strongly disagree | 23.9% (22/92) | 95.7% (44/46) | 38.6% (51/132) | 1.0 |  |
| Accumulative results of BC are helpful in understanding epidemiology of AMR bacterial infections |  |  |  |  |  |
| Strongly agree / Agree | 31.5% (152/483) | 90.5% (258/285) | 52.5% (240/457) | 2.89 (1.60-5.19) | <0.001 |
| Uncertain / Disagree / Strongly disagree | 21.4% (3/14) | 76.5% (13/17) | 25% (10/40) | 1.0 |  |
| BC is unnecessary because antibiotic therapy can be determined based on clinical presentation |  |  |  |  |  |
| Strongly agree / Agree | 20.8% (21/101) | 83.6% (46/44) | 33.8% (24/71) | 0.51 (0.36-0.73) | <0.001 |
| Uncertain / Disagree / Strongly disagree | 33.9% (134/395) | 91.1% (226/248) | 53.3% (228/428) | 1.0 |  |
| The therapeutic consequence of BC is questionable |  |  |  |  |  |
| Strongly agree / Agree | 32.3% (30/93) | 88.0% (73/83) | 41.0% (25/61) | 0.84 (0.59-1.19) | 0.32 |
| Uncertain / Disagree / Strongly disagree | 30.6% (120/392) | 91.2% (196/215) | 51.9% (223/430) | 1.0 |  |
| The scientific basis of the guideline on BC is questionable |  |  |  |  |  |
| Strongly agree / Agree | 32.0% (17/53) | 87.3% (69/79) | 32.8% (19/58) | 0.66 (0.45-0.98) | 0.04 |
| Uncertain / Disagree / Strongly disagree | 30.4% (132/433) | 91.2% (198/217) | 53.2% (231/434) | 1.0 |  |
| BC is unnecessary because results are often delayed |  |  |  |  |  |
| Strongly agree / Agree | 18.9% (24/127) | 82.1% (32/39) | 30.2% (16/53) | 0.48 (0.33-0.69) | <0.001 |
| Uncertain / Disagree / Strongly disagree | 35.2% (129/367) | 90.9% (240/264) | 53.0% (236/445) | 1.0 |  |
| BC is unnecessary because results are often not interpretable |  |  |  |  |  |
| Strongly agree / Agree | 25.0% (13/52) | 77.3% (17/22) | 29.7% (11/37) | 0.54 (0.34-0.87) | 0.01 |
| Uncertain / Disagree / Strongly disagree | 31.7% (140/442) | 90.8% (255/281) | 52.3% (241/461) | 1.0 |  |
| BC is unnecessary because results are often negative or no growth |  |  |  |  |  |
| Strongly agree / Agree | 30.8% (20/65) | 81.3% (26/32) | 28.0% (14/50) | 0.58 (0.39-0.88) | 0.01 |
| Uncertain / Disagree / Strongly disagree | 30.8% (131/426) | 91.1% (247/271) | 53.1% (238/448) | 1.0 |  |
| BC is unnecessary because cultures are often contaminated |  |  |  |  |  |
| Strongly agree / Agree | 26.3% (19/72) | 79.3% (23/29) | 34.2% (14/41) | 0.64 (0.42-0.98) | 0.04 |
| Uncertain / Disagree / Strongly disagree | 31.9% (133/417) | 90.9% (249/274) | 52.2% (236/452) | 1.0 |  |
| BC is unnecessary because results often do not agree with clinical signs |  |  |  |  |  |
| Strongly agree / Agree | 34.0% (18/53) | 88.9% (24/27) | 23.5% (8/34) | 0.77 (0.48-1.22) | 0.27 |
| Uncertain / Disagree / Strongly disagree | 30.8% (135/439) | 89.9% (249/277) | 52.9% (241/456) | 1.0 |  |
| BC is unnecessary because it is too expensive |  |  |  |  |  |
| Strongly agree / Agree | 25.5% (24/94) | 80.0% (24/30) | 32.7% (17/52) | 0.62 (0.42-0.92) | 0.02 |
| Uncertain / Disagree / Strongly disagree | 32.4% (129/398) | 91.2% (249/273) | 52.9% (229/443) | 1.0 |  |
| BC is not benefiting the patients |  |  |  |  |  |
| Strongly agree / Agree | 14.0% (15/107) | 84.0% (21/25) | 19.4% (7/36) | 0.37 (0.24-0.57) | <0.001 |
| Uncertain / Disagree / Strongly disagree | 35.8% (136/380) | 90.1% (246/273) | 53.0% (239/451) | 1.0 |  |
| BC is unnecessary because a contaminated result often leads to wrong therapeutic approaches |  |  |  |  |  |
| Strongly agree / Agree | 30.4% (7/23) | 86.4% (19/22) | 20.0% (6/30) | 0.53 (0.30-0.95) | 0.03 |
| Uncertain / Disagree / Strongly disagree | 31.5% (148/470) | 90.1% (254/282) | 52.5% (245/467) | 1.0 |  |
| It is not too late to collect BC later, particularly if patients do not improve after receiving empirical antibiotic treatment |  |  |  |  |  |
| Strongly agree / Agree | 13.8% (19/138) | 88.3% (143/162) | 31.2% (38/122) | 0.37 (0.27-0.51) | <0.001 |
| Uncertain / Disagree / Strongly disagree | 38.1% (134/352) | 91.6% (130/142) | 57.2% (214/373) | 1.0 |  |
| Quality of laboratory is questionable |  |  |  |  |  |
| Strongly agree / Agree | 24.2% (22/91) | 84.2% (32/38) | 26.6% (17/64) | 0.48 (0.33-0.70) | <0.001 |
| Uncertain / Disagree / Strongly disagree | 32.7% (128/391) | 90.3% (232/257) | 54.1% (230/435) | 1.0 |  |
| Levels of local antibiotic resistance are low |  |  |  |  |  |
| Strongly agree / Agree | 34.7% (17/49) | 76.9% (20/26) | 32.0% (16/50) | 0.64 (0.41-0.98) | 0.04 |
| Uncertain / Disagree / Strongly disagree | 31.3% (135/432) | 91.1% (246/270) | 52.8% (235/445) | 1.0 |  |
| **TDF Domain: Memory, attention and decision processes** |  |  |  |  |  |
| Deciding to do BC in patients presenting with sepsis |  |  |  |  |  |
| Yes | 34.1% (130/381) | 90.2% (259/287) | 54.2% (219/404) | 1.79 (1.27-2.52) | 0.001 |
| No | 22.5 (27/120) | 82.4% (14/17) | 34.7% (33/95) | 1.0 |  |
| Deciding to do BC in patients presenting with septic shock |  |  |  |  |  |
| Yes | 35.2% (114/324) | 89.8% (246/274) | 50.7% (216/426) | 1.27 (0.93-1.75) | 0.14 |
| No | 24.3% (43/177) | 90.0% (27/30) | 49.3% (36/73) | 1.0 |  |
| Deciding to do BC in patients starting parenteral antibiotic treatment |  |  |  |  |  |
| Yes | 47.0% (71/151) | 93.1% (190/204) | 65.0% (76/117) | 2.32 (1.75-3.09) | <0.001 |
| No | 24.6% (86/350) | 83.0% (83/100) | 46.1% (176/382) | 1.0 |  |
| Even when BC is recommended, would you still order BC if patients are already on antibiotics |  |  |  |  |  |
| Definitely not order / likely not order | 20.0% (6/30) | 86.6% (58/67) | 41.2% (14/34) | 0.69 (0.42-1.11) | 0.13 |
| Maybe not order/ likely to still order / very likely to still order | 31.2% (142/455) | 90.6% (213/235) | 51.3% (238/464) | 1.0 |  |
| Even when BC is recommended, would you still order BC if patients have anemia |  |  |  |  |  |
| Definitely not order / likely not order | 21.3% (16/75) | 91.9% (136/148) | 47.4% (27/57) | 0.89 (0.62-1.28) | 0.55 |
| Maybe not order/ likely to still order / very likely to still order | 32.2% (128/398) | 87.3% (130/149) | 51.3% (220/429) | 1.0 |  |
| **TDF: Optimism** |  |  |  |  |  |
| Strongly optimistic / Optimistic | 33.3% (133/400) | 90.5% (238/263) | 54.4% (200/368) | 1.78 (1.29-2.46) | <0.001 |
| Neither / Pessimistic / Strongly pessimistic | 20.7% (18/87) | 88.6% (31/35) | 39.8% (51/128) | 1.0 |  |
| **TDF: Skills** |  |  |  |  |  |
| How skilled are you in drawing blood? ^4^ |  |  |  |  |  |
| Very good / Good | 38.5% (15/39) | 88.2% (30/34) | 57.1% (40/70) | 1.74 (1.02-2.97) | 0.04 |
| Fair / Poor / Very poor | 31.8% (14/44) | 93.1% (81/87) | 35.1% (20/57) | 1.0 |  |
| **TDF: Beliefs about capabilities** |  |  |  |  |  |
| Are you confident that you can draw blood successfully? ^4,7^ |  |  |  |  |  |
| Strongly confident / Confident | 34.7% (25/72) | 89.1% (57/64) | 51.9% (56/108) | 1.39 (0.69-2.79) | 0.36 |
| Uncertain / Doubtful / Strongly doubtful | 36.4% (4/11) | 94.7% (54/57) | 22.2% (4/18) | 1.0 |  |
| Are you confident that you can draw blood appropriately? ^4,7^ |  |  |  |  |  |
| Strongly confident / Confident | 34.8% (24/69) | 89.7% (70/78) | 54.6% (54/99) | 1.67 (0.88-3.17) | 0.11 |
| Uncertain / Doubtful / Strongly doubtful | 35.7% (5/14) | 95.2% (40/42) | 22.2% (6/27) | 1.0 |  |
| Are you confident that others (who are tasked to draw blood in your hospital) can draw blood successfully? ^7^ |  |  |  |  |  |
| Strongly confident / Confident | 30.7% (142/463) | 90.1% (254/282) | 52.5% (212/404) | 1.35 (0.91-2.00) | 0.13 |
| Uncertain / Doubtful / Strongly doubtful | 33.3% (11/33) | 85.7% (18/21) | 43.0% (40/93) | 1.0 |  |
| Are you confident that others (who are tasked to draw blood in your hospital) can draw blood appropriately? ^7^ |  |  |  |  |  |
| Strongly confident / Confident | 31.0% (132/426) | 89.6% (224/250) | 52.8% (168/318) | 1.20 (0.89-1.62) | 0.23 |
| Uncertain / Doubtful / Strongly doubtful | 31.9% (22/69) | 90.6% (48/53) | 46.6% (83/178) | 1.0 |  |
| **TDF: Emotion** |  |  |  |  |  |
| Any emotional factors |  |  |  |  |  |
| Yes | 25.5% (13/51) | 80% (8/10) | 65.6% (21/32) | 1.06 (0.65-1.71) | 0.82 |
| No | 32.0% (144/450) | 90.1% (265/294) | 49.5% (231/467) | 1.0 |  |

^1^ Percentage of participants who answered with “definitely take BC” in the case scenario are presented. For each question, participants who answered ‘I do not know’ or ‘I do not want to answer’ were excluded. ^2^ Estimated by using logistic regression models with random effects for countries, for types of hospital nested in the same country, and for professional roles nested in the same types of hospital. ^3^ Among those who answered that they know of local guidelines. ^4^ Among those who answered that their professional roles are tasked of drawing blood for BC. ^5^ “Norms” means usual practice that are typical of or accepted within your hospital. ^6^ Included answers in Q1-7 (which were asked to those who answered that they knew of local guideline) and Q1-8 (which were asked to those who answered that they did not know of local guideline) (Appendix S4). ^7^ “Successfully” means obtaining blood; “Appropriately” means that general recommendations for BC specimen collection such as aseptic technique are followed.

**Appendix S13. Links between TDF, COM-B components (Capability, Opportunity, motivation and behaviour components), and suggested intervention types and policies.**

Links between TDF and COM-B components*

| **COM-B components** |  | **TDF Domains** |
| --- | --- | --- |
| **Capability** | **Psychological** | **Knowledge** |
|  |  | **Skills** |
|  |  | **Memory, attention and decision processes** |
|  |  | **Behavioural regulation** |
|  | **Physical** | **Skills** |
| **Opportunity** | **Social** | **Social Influences** |
|  | **Physical** | **Environmental Context and Resources** |
| **Motivation** | **Reflective** | **Social/professional role and Identity** |
|  |  | **Beliefs about capabalities** |
|  |  | **Optimism** |
|  |  | **Beliefs about Consequences** |
|  |  | **Intentions** |
|  |  | **Goals** |
|  | **Automatic** | **Social/professional role and Identity** |
|  |  | **Optimism** |
|  |  | **Reinforcement** |
|  |  | **Emotion** |

*as previously published.^42^

Links between COM-B components and intervention types*

| **Intervention functions** | **COM-B components** | | | | | |
| --- | --- | --- | --- | --- | --- | --- |
|  | **Capability** | | **Opportunity** | | **Motivation** | |
|  | **Psychological** | **Physical** | **Social** | **Physical** | **Reflective** | **Automatic** |
| **Education** |  | X |  |  | X |  |
| **Persuasion** |  |  |  |  | X | X |
| **Incentivisation** |  |  |  |  | X | X |
| **Coerction** |  |  |  |  | X | X |
| **Training** | X | X |  |  |  |  |
| **Restriction** |  |  | X | X |  |  |
| **Environmental restructuring** |  |  | X | X |  | X |
| **Modelling** |  |  |  |  |  | X |
| **Enablement** | X | X | X | X |  | X |

* as previously published.^43^

Links between intervention functions and policy categories*

| **Intervention functions** | **Policy categories** | | | | | | |
| --- | --- | --- | --- | --- | --- | --- | --- |
|  | **Communication/Marketing** | **Guidelines** | **Fiscal** | **Regulation** | **Legislation** | **Environmental/social planning** | **Service Provision** |
| **Education** | X | X |  | X | X |  | X |
| **Persuasion** | X | X |  | X | X |  | X |
| **Incentivisation** | X | X | X | X | X |  | X |
| **Coerction** | X | X | X | X | X |  | X |
| **Training** |  | X | X | X | X |  | X |
| **Restriction** |  | X |  | X | X |  |  |
| **Environmental restructuring** |  | X | X | X | X | X |  |
| **Modelling** | X |  |  |  |  |  | X |
| **Enablement** |  | X | X | X | X | X | X |

* as previously published.^43^

**Appendix S14. References**
